# Deep Learning-Based Multimodal Clustering Model for Endotyping and Post-Arthroplasty Response Classification using Knee Osteoarthritis Subject-Matched Multi-Omic Data

**DOI:** 10.1101/2024.06.13.24308857

**Authors:** Jason S. Rockel, Divya Sharma, Osvaldo Espin-Garcia, Katrina Hueniken, Amit Sandhu, Chiara Pastrello, Kala Sundararajan, Pratibha Potla, Noah Fine, Starlee S. Lively, Kimberly Perry, Nizar N. Mohamed, Khalid Syed, Igor Jurisica, Anthony V. Perruccio, Y. Raja Rampersaud, Rajiv Gandhi, Mohit Kapoor

## Abstract

**Background:** Primary knee osteoarthritis (KOA) is a heterogeneous disease with clinical and molecular contributors. Biofluids contain microRNAs and metabolites that can be measured by omic technologies. Deep learning captures complex non-linear associations within multimodal data but, to date, has not been used for multi-omic-based endotyping of KOA patients. We developed a novel multimodal deep learning framework for clustering of multi-omic data from three subject-matched biofluids to identify distinct KOA endotypes and classify one-year post-total knee arthroplasty (TKA) pain/function responses.

**Materials and Methods:** In 414 KOA patients, subject-matched plasma, synovial fluid and urine were analyzed by microRNA sequencing or metabolomics. Integrating 4 high-dimensional datasets comprising metabolites from plasma (n=151 features), along with microRNAs from plasma (n=421), synovial fluid (n=930), or urine (n=1225), a multimodal deep learning variational autoencoder architecture with K-means clustering was employed. Features influencing cluster assignment were identified and pathway analyses conducted. An integrative machine learning framework combining 4 molecular domains and a clinical domain was then used to classify WOMAC pain/function responses post-TKA within each cluster.

**Findings:** Multimodal deep learning-based clustering of subjects across 4 domains yielded 3 distinct patient clusters. Feature signatures comprising microRNAs and metabolites across biofluids included 30, 16, and 24 features associated with Clusters 1-3, respectively. Pathway analyses revealed distinct pathways associated with each cluster. Integration of 4 multi-omic domains along with clinical data improved response classification performance, with Cluster 3 achieving AUC=0·879 for subject pain response classification and Cluster 2 reaching AUC=0·808 for subject function response, surpassing individual domain classifications by 12% and 15% respectively.

**Interpretation:** We have developed a deep learning-based multimodal clustering model capable of integrating complex multi-fluid, multi-omic data to assist in KOA patient endotyping and test outcome response to TKA surgery.

**Funding:** Canada Research Chairs Program, Tony and Shari Fell Chair, Campaign to Cure Arthritis, University Health Network Foundation.

## Introduction

Osteoarthritis (OA) is a degenerative, painful and disabling joint disease affecting over 500 million people worldwide,^1^ with the knee most commonly afflicted^2^. Primary knee (K)OA patients are heterogeneous^3^. Risk factors of KOA include age, sex, and obesity status.^2^ Mental health and persistent pain status have also associated with KOA clinical phenotypes.^4,5^ Total joint arthroplasty (TKA) is the only available therapy for KOA patients who no longer respond to conservative management; however, up to 34% of KOA patients with TKA fail to achieve clinically-relevant pain reduction.^6^ Identifying those at high risk of non-response is of significant interest. It is possible that KOA heterogeneity captured by biological features may improve our ability to classify patient responses to TKA.

Biofluid microRNAs (miRNAs) and metabolites can provide highly descriptive, individualized categorizations of patients beyond clinical measures. MiRNAs epigenetically modify target RNA expression. Biofluid metabolomes represents snapshots of the metabolic activity contributed by associated cells and tissues. Advanced omic technologies can measure miRNAs (miRNomics) and metabolites (metabolomics), primarily by next generation sequencing (NGS) and liquid chromatography-mass spectrometry/mass spectrometry (LC-MS/MS), respectively.^7,8^

In a case-control study using the UK Biobank cohort, 14 distinct OA risk phenotypes were identified by multi-modal machine learning (ML) using clinical factors alone.^9^ The inclusion of individual proteomics, genomics or metabolomics data showed no prediction improvement of case-control status over clinical factor modeling alone.^9^ In contrast, studies using individual biofluids have identified endotypes of OA patients. Three endotypes of OA patients (low tissue turnover, structural damage and systemic inflammation) were identified from a panel of 16 serum and urine proteins/peptides using unsupervised ML.^10^ Plasma metabolomics alone uncovered multiple endotypes of KOA patients,^11,12^ with some endotype-related metabolite ratios able to differentiate specific endotypes of KOA subjects from control participants.^11^ KOA patient biofluid miRNA signatures were also able to differentiate between slow and fast progressors,^13^ early and late KOA,^14,15^ and patients requiring TKA or not.^16^ Thus, endotype data from biofluids is important for understanding KOA heterogeneity, and, consequently, may be associated with patient outcomes. To our knowledge, KOA endotypes have yet to be evaluated across multiple biofluids using multi-omic technologies in an integrated approach.^8^

Our experience to date suggests that multi-biofluid, mult-omic endotyping requires more complex modeling systems. Deep learning aids in extracting complex patterns from data. However, integrating multimodal data from multiple sources presents challenges due to the diversity within and across data types. Variational Autoencoders (VAEs) address this challenge by embedding diverse data domains into reduced latent dimensions, facilitating improved data clustering.^17–19^ Despite the potential of VAEs, there is a lack of unified frameworks for leveraging these methods to identify clusters from multimodal data and to classify clinical responses by integrating diverse data domains. Additionally, VAEs have not been applied to investigate OA endotypes.

In this study, we developed a novel multimodal deep learning framework employing VAEs for integrative clustering using 4 high-dimensional domains of subject-matched multi-omic data from synovial fluid, urine and plasma and tested its ability to determine distinct clusters (endotypes) of a sample of KOA patients. Leveraging these endotypes, we further developed an integrative ML framework and tested the potential of this methodology to assess pain and function responses to TKA surgery.

## Methods

### Study Sample

A sample of 414 patients with primary KOA who underwent TKA within the Longitudinal Evaluation in the Arthritis Program-OA cohort (University Health Network, Toronto, ON, Canada), as previously described,^20^ and who had synovial fluid (collected intra-operatively), plasma and urine (collected up to 3 months prior to surgery) available, were selected for analysis. Subjects completed self-reported, multidimensional questionnaires from which baseline Western Ontario and McMaster Universities Arthritis Index (WOMAC) pain and function,^21^ Hospital Anxiety and Depression Scale (HADS)^22^ and painDETECT^23^ at baseline (completed within the 3-months preceding surgery), and WOMAC pain and function 1-year post-TKA, were calculated. Improvement in WOMAC pain and function from baseline to 1 year post-TKA was calculated and individuals were categorized as responders (>33% improvement) or non-responders (≤33% improvement). HADS depression and anxiety scores were each categorized as normal (score 0-7), borderline case (score 8-11), and definite case (score 11-21)^22^. PainDETECT neuropathic-like pain scores were used to classify patients pain as likely nociceptive (score 0-12), unclear (score 13-18), or likely neuropathic (score 19-38).^23^ Baseline age, sex, and weight and height [from which body mass index (BMI; kg/m^2^) was calculated] were also collected. Biofluids and patient data were collected with written informed consent from each patient and Research Ethics Board approval of the University Health Network, Toronto, ON (REB# 07-0383-BE; 14-7592-AE).

### MiRNA sequencing (miRNomics) and metabolomics

MiRNA extraction was performed using 200 µL plasma (N=414), 100 µL synovial fluid (N=414), and 1 ml urine (N=414). cDNA libraries were prepared using a protocol we previously reported.^15^ NGS was conducted at the Schroeder Arthritis Institute (Toronto) sequencing facility using the Illumina NextSeq550 platform. Alignment, processing and quality assessment was performed using a previously reported pipeline.^24^ Targeted metabolomics was used to profile 188 metabolites (Biocrates AbsoluteIDQ p180 kit, Biocrates Life Sciences AG, Austria) in N=414 plasma samples at The Metabolomics Innovation Centre (Calgary, Alberta) by LC-MS/MS, as previously described.^12^ Metabolite quantification and batch correction was conducted using the Absolute IDQ-coupled MetIDQ software (Biocrates). MiRNA count data and metabolite concentrations were normalized using sum normalization, log-transformation, and Pareto scaling.^25^ To stabilize variance estimates within differential expression analysis, empirical Bayes moderation techniques were applied.

### OmicVAE: integrative variational autoencoder architecture for multimodal clustering

We generated a novel variational autoencoder (VAE) architecture named ‘*omicVAE*’ designed to cluster multimodal multi-omic data (Figure 1). *OmicVAE* consists of a single encoder network followed by 4 individual decoder networks, to perform integrative clustering combining 4 modalities: metabolomics, miRNA plasma, miRNA synovial fluid, and miRNA urine.

**Figure 1:**
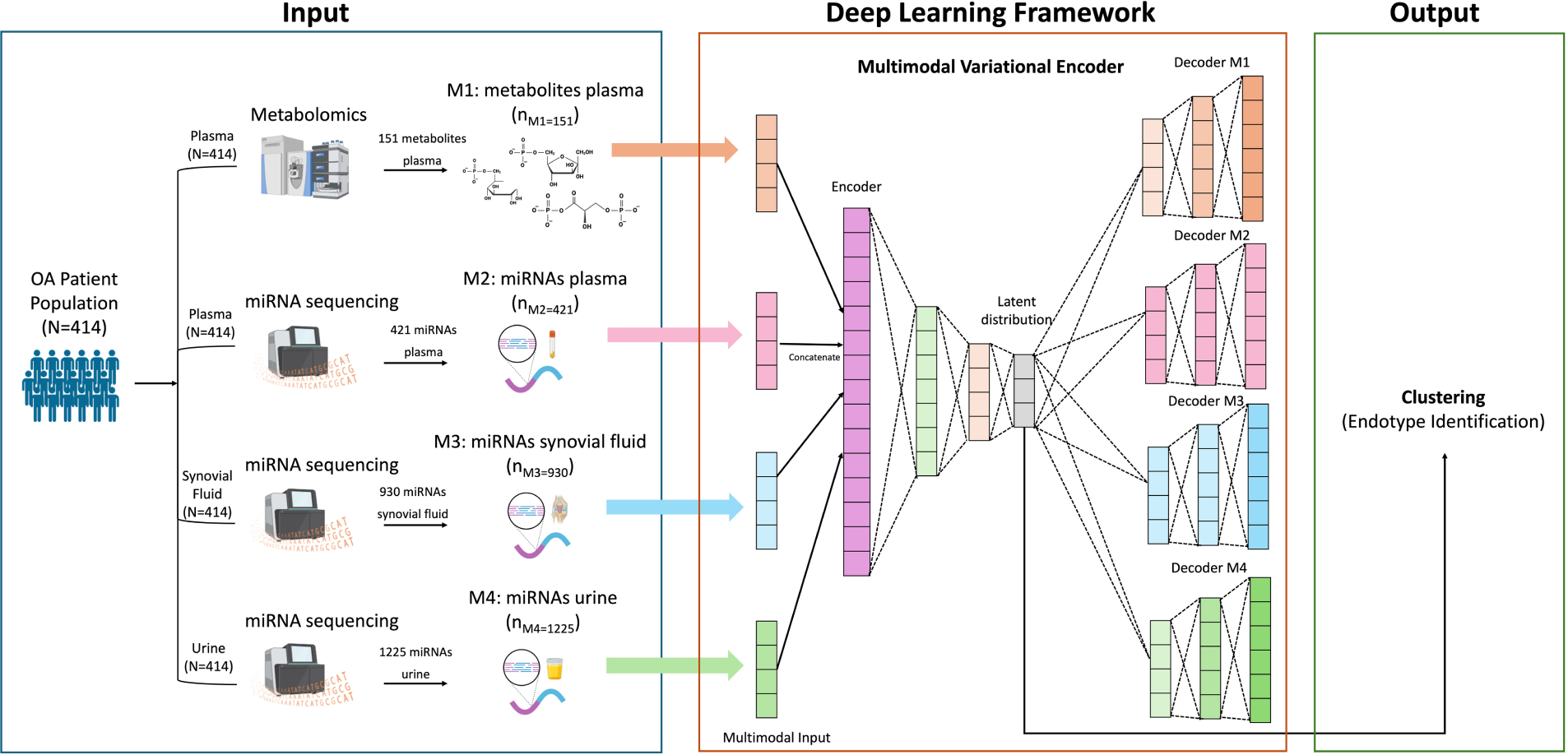
Overall framework for Deep Learning-based Multimodal Clustering. Deep-learning framework for multimodal integration and clustering using Variational Autoencoder (VAE) modeling. Four individual decoders in the proposed VAE accurately identify the latent distribution within each domain (M1-M4) capturing the complex non-linear associations within the multimodal data. K-means approach is used for identifying three clusters from the obtained latent distribution. ‘N’ represents the no. of patients and ‘nMx’ represents the number of features in each data domain.

The encoder network inputs concatenated multimodal multi-omic data and maps it to a shared latent space representation using multiple fully connected neural network (FNN) layers with sigmoid activation. The encoder network’s output layers parameterize the mean and variance of a Gaussian distribution representing the shared latent space. Each decoder network reconstructs its respective domain’s input data from samples drawn from this latent space, using multiple fully connected neural network (FNN) layers. During training, variational inference optimizes the VAE’s parameters. The objective function *L_total_* includes the reconstruction loss (*L_rec_*) and the Kullback-Leibler (KL) divergence (*L_KL_*) between the learned latent distribution and a predefined prior (eqs. 1-3). Minimizing KL divergence regularizes the latent space, preventing overfitting and ensuring it remains structured and interpretable. The reconstruction loss measures the discrepancy between the input data and its reconstruction by the VAE decoder for each modality.

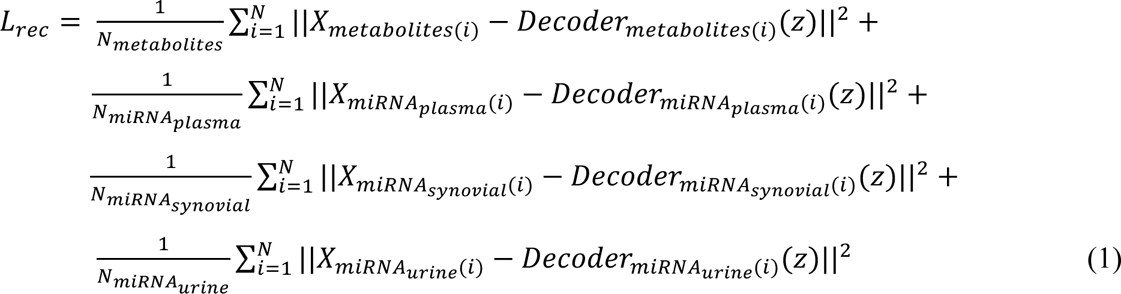

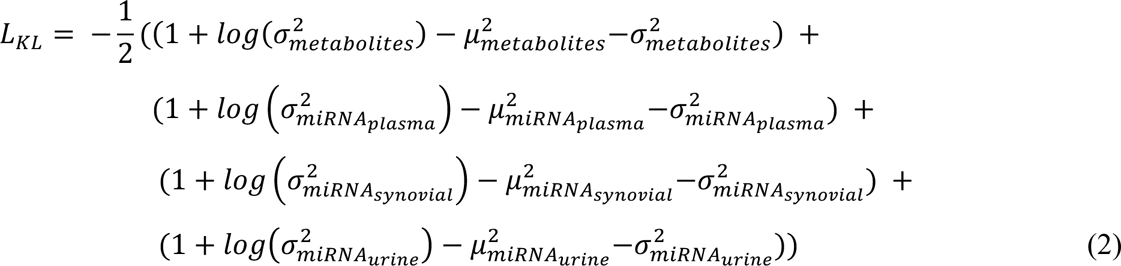

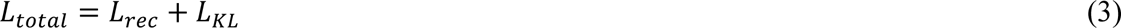

where i represents the samples in each modality, X is the input data, Decoder(z) is the reconstructed data, and *μ* and *σ* denote the mean and variance of the Gaussian distribution in the latent space. Once omicVAE is trained, K-means clustering on the learned latent space is used to identify distinct subpopulations within the multimodal multi-omic data.

### Multi-omic signature identification within each cluster

We employed a comprehensive approach to identify signature features (miRNAs and/or metabolites within three biofluids) influencing cluster assignment within each domain. We concurrently conducted standardized mean differences (SMD) analysis and differential expression (DE) analysis for pairwise cluster comparisons (one cluster vs others), using Benjamini-Hochberg (BH) adjusted p-values (q < 0.05) to identify significant features. By integrating these analyses, we identified features with both large SMDs and significant DE, capturing robust signature features distinguishing clusters.

### Endotype pathway analysis

MiRNAs gene targets per cluster were identified using the top 1% of targets per miRNA using mirDIP v. 5.2 (https://ophid.utoronto.ca/mirDIP)^26^ We performed pathway enrichment analysis for sets of gene targets in each cluster using pathDIP 5 (https://ophid.utoronto.ca/pathDIP).^27^ Diseases, drugs and vitamins, and genetic information processing pathway types were excluded from enrichment analysis. Only pathways with q-value (BH adjusted) <0.01 were considered. Metabolite pathway enrichment analysis was not possible, so we identified pathways that included metabolites specific for each cluster for further analyses. Selected pathways specific for each cluster were visualized using NAViGaTOR 3.0.19 (https://navigator.ophid.utoronto.ca/navigatorwp).^28^ Mapping of pathways to consolidated categories in pathDIP was used to calculate the number of pathways per category. ggradar2_1.1.0 in R 4.3.0 was subsequently used to plot their distribution per cluster, scaling category pathway counts from 0% to 100%.

### Integrative machine learning framework for classifying response

We developed a comprehensive two-step ML framework (Figure 4a) to integrate plasma metabolites, miRNA plasma, miRNA synovial fluid, and miRNA urine domain, along with clinical domain (consisting of age, sex, BMI, depression and anxiety categories, and neuropathic pain category), to classify 1-year pain and function responses (i.e. responders vs non-responders). In the first step, we trained separate unimodal ML models for each domain to extract features classifying 1-year response. We utilized the MICE library in R for imputing missing clinical data (missingness <8%). We explored various ML algorithms, including logistic regression, lasso regression, ridge regression, support vector machines and random forests, selecting models based on 10-fold cross-validation performance.

In the second step, we integrated features from all domains using a naïve-Bayes meta-classifier, trained with classifiers from the unimodal models. Cross-validation was used for performance estimation and hyperparameter tuning. The final classification was generated by the meta-classifier based on integrated features. Model evaluation involved assessing the overall framework performance using area under the receiver operating characteristic curve (AUC) and analyzing feature importance with mean Gini impurity metrics.

## Findings

### Endotype and signature identification using omicVAE and K-means clustering

We first sought to identify endotypes from our sample of 414 KOA patients. The patient sample was 57% female, with a mean age (±sd) of 65·7±8·7 years, and BMI (±sd) of 31±7·1 kg/m^2^. The majority of subjects had anxiety or depression symptom scores in the normal range and the majority had painDETECT scores indicating likely nociceptive pain. Mean baseline WOMAC pain score for the sample was 10·1±3·5 points on a 20 total point scale, and baseline WOMAC function score was 34·9 ± 11·9 points on a 68 total point scale (Supplementary Table 1).

After metabolomics and miRNomics analyses of plasma, synovial fluid and urine, our analytical dataset consisted of 2727 molecular features from 4 domains: 151 plasma metabolites, 421 plasma miRNAs, 930 synovial fluid miRNAs, and 1225 urine miRNAs. We then developed *omicVAE* with K-means clustering (Figure 1), which uncovered three clusters of patients using the 4 domains (Figure 2a). Distribution of most baseline clinical, demographic and anthropometric measures were similar across clusters, except cluster 3 that had a higher proportion of subjects with depression scores in the normal range, and cluster 1 that had a higher proportion of subjects with likely neuropathic pain (Table 1).

**Figure 2.**
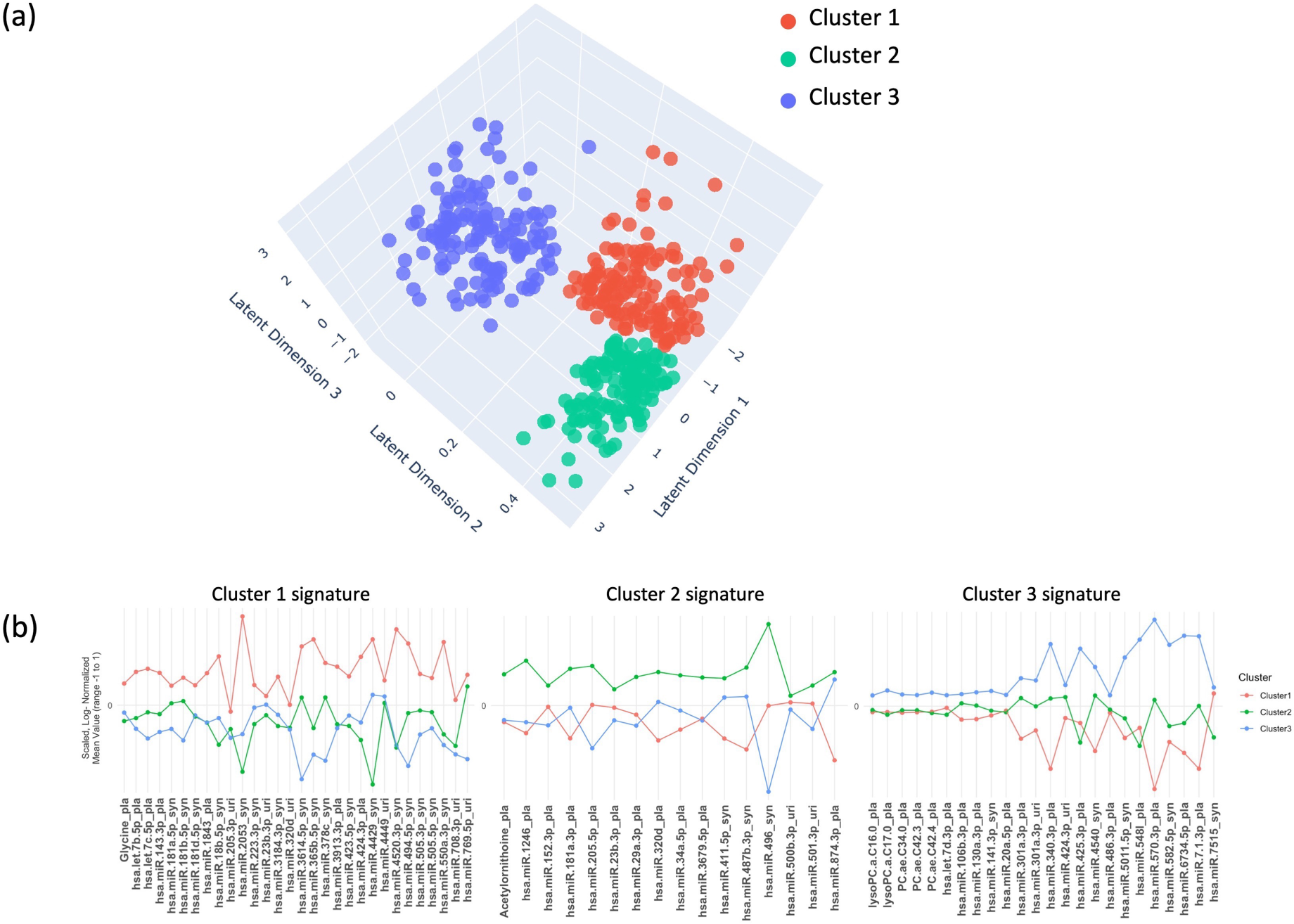
Integrated analysis of patient clustering and miRNA and metabolite feature signatures. (a) Three-dimensional illustration (latent dimension 1-3) of the three clusters obtained using a Variational Autoencoder-based deep learning framework. Cluster 1 (red) comprises 146 patients, Cluster 2 (green) consists of 138 patients, and Cluster 3 (blue) includes 130 patients. (b) Molecular signature profiles for clusters 1, 2, and 3 respectively (left to right) derived through the intersection of the most significant variables (p < 0.05) identified from both standardized mean differences analysis and differential expression analysis within the plasma metabolites and miRNA domains, differentiating each cluster.

**Table 1.** Summary statistics of clinical variables within each cluster identified from variational autoencoder machine-learning modeling and K-means clustering. Frequency (percentage) are provided for categorical variables while median (Quartile 1, Quartile 3) values are presented for continuous variables according to patients within each cluster. p value is computed using wilcoxon rank sum test for continuous variables and chi-square or fisher’s test as appropriate for categorical variables.

|  | Cluster 1 (n=146) | Cluster 2 (n=138) | Cluster 3 (n=130) | p-value between clusters |
| --- | --- | --- | --- | --- |
| **Sex** |  |  |  | 0.9 |
| Female [N, (%)] | 84 (58) | 76 (55) | 72 (55) |  |
| Male [N, (%)] | 62 (42) | 62 (45) | 58 (45) |  |
| **Age** |  |  |  | 0.37 |
| Mean (sd) | 66.2 (8.2) | 65.4 (8.0) | 65.5 (8.9) |  |
| Median (Q1,Q3) | 67 (62, 71) | 65.0 (60.0, 70.8) | 66 (60, 72) |  |
| **BMI** |  |  |  | 0.55 |
| Mean (sd) | 30.7 (6.1) | 31.9 (7.7) | 31.4 (7.3) |  |
| Median (Q1,Q3) | 29.2 (26.4, 33.8) | 30.0 (26.5, 35.7) | 31.1 (25.3, 34.7) |  |
| **HADS anxiety** |  |  |  | 0.72 |
| normal [N, (%)] | 103 (72) | 98 (73) | 100 (79) |  |
| borderline [N, (%)] | 21 (15) | 20 (15) | 13 (10) |  |
| case [N, (%)] | 19 (13) | 17 (13) | 14 (11) |  |
| Missing (N) | 3 | 3 | 3 |  |
| **HADS depression** |  |  |  | 0.037 |
| normal [N, (%)] | 107 (74) | 105 (78) | 112 (87) |  |
| borderline [N, (%)] | 23 (16) | 19 (14) | 6 (5) |  |
| case [N, (%)] | 15 (10) | 11 (8) | 11 (9) |  |
| Missing (N) | 1 | 3 | 1 |  |
| **painDETECT** |  |  |  | 0.011 |
| neuropathic [N, (%)] | 30 (21) | 14 (10) | 11 (8) |  |
| nociceptive [N, (%)] | 86 (59) | 82 (59) | 87 (67) |  |
| unclear [N, (%)] | 27 (18) | 31 (22) | 23 (18) |  |
| Missing [N, (%)] | 3 (2) | 11 (8) | 9 (7) |  |
| **WOMAC pain baseline** |  |  |  | 0.98 |
| Mean (sd) | 10.2 (3.5) | 10.0 (3.2) | 10.0 (3.7) |  |
| Median (Q1,Q3) | 10 (8, 12) | 10 (8, 12) | 10.0 (8.0, 12.4) |  |
| **WOMAC pain 1yr** |  |  |  | 0.34 |
| Mean (sd) | 3.7 (4.0) | 3.7 (3.6) | 3.2 (3.6) |  |
| Median (Q1,Q3) | 3.0 (1.0, 5.8) | 3 (1, 6) | 2.0 (0.2, 5.0) |  |
| **WOMAC pain change (1 yr - baseline)** |  |  |  | 0.55 |
| Mean (sd) | -6.5 (4.1) | -6.3 (3.9) | -6.8 (4.1) |  |
| Median (Q1,Q3) | -7 (-9, -4) | -6.0 (-9.0, -3.2) | -7.0 (-9.8, -4.0) |  |
| **WOMAC pain change categorical** |  |  |  | 0.48 |
| ≤33.3% [N, (%)] | 27 (18) | 33 (24) | 22 (17) |  |
| >33.3% [N, (%)] | 119 (82) | 105 (76) | 108 (83) |  |
| **WOMAC function baseline** |  |  |  | 0.74 |
| Mean (sd) | 34.7 (11.9) | 34.9 (11.4) | 33.8 (12.3) |  |
| Median (Q1,Q3) | 36 (26, 41) | 35.0 (26.2, 43.0) | 34 (26, 41) |  |
| Missing (N) | 1 | 0 | 1 |  |
| **WOMAC function 1 yr** |  |  |  | 0.24 |
| Mean (sd) | 14.8 (13.7) | 14.7 (12.9) | 12.8 (13.0) |  |
| Median (Q1,Q3) | 12.0 (4.2, 21.0) | 11.5 (4.0, 20.8) | 9.0 (2.2, 19.0) |  |
| **WOMAC function change (1 year- baseline)** |  |  |  | 0.76 |
| Mean (sd) | -20.3 (13.4) | -20.0 (13.8) | -20.2 (13.3) |  |
| Median (Q1,Q3) | -20 (-30, -11) | -18.4 (-31.0, -10.0) | -19.0 (-28.8, -11.0) |  |
| **WOMAC function change categorical** |  |  |  | 0.24 |
| ≤33.3% [N, (%)] | 33 (23) | 32 (23) | 24 (19) |  |
| >33.3% [N, (%)] | 112 (77) | 106 (77) | 105 (81) |  |
| Missing (N) | 1 | 0 | 1 |  |

BMI, body mass index; HADS, hospital anxiety and depression scale; Q, quartile; sd, standard deviation.

Significant features associated with each cluster were identified by the intersection of differential expression and standardized mean difference analyses. Distinct signatures consisting of 30, 16 and 24 features for clusters 1-3, respectively, were identified (Figure 2b and Supplementary Table 2). Notably, each signature contained features from all 4 domains. In Cluster 1, the highest mean value difference was observed for synovial fluid hsa-miR-2053. In Cluster 2, synovial fluid hsa-miR-496 exhibited the highest mean value. In Cluster 3, plasma hsa-miR-570-3p had the highest mean value. Thus, each cluster represents a group of subjects with a distinct endotype.

### Endotype feature signatures are enriched for unique pathways

We next sought to determine if cluster endotype signatures were associated with unique physiological pathways. We first identified putative miRNA-gene targets using mirDIP,^26^ identifying 3257, 2211, and 2319 individual genes targeted by the miRNAs in each of the endotype signatures associated with clusters 1-3, respectively. Using these gene set lists, we performed enrichment analysis using pathDIP (Supplementary Table 3-5).^27^ For metabolites in each endotype signature, pathway annotations were also identified using pathDIP (Supplementary Table 6-8). All pathways were also annotated with categories in pathDIP. For each endotype, individual miRNA-gene targets and metabolites were linked to some common as well as unique pathways. The top unique enriched and annotated pathways linked to miRNA-targeted genes or metabolites, respectively, for each endotype are displayed in a network showing individual pathways and categories (Figure 3a, Supplementary Figures 1-3).

**Figure 3.**
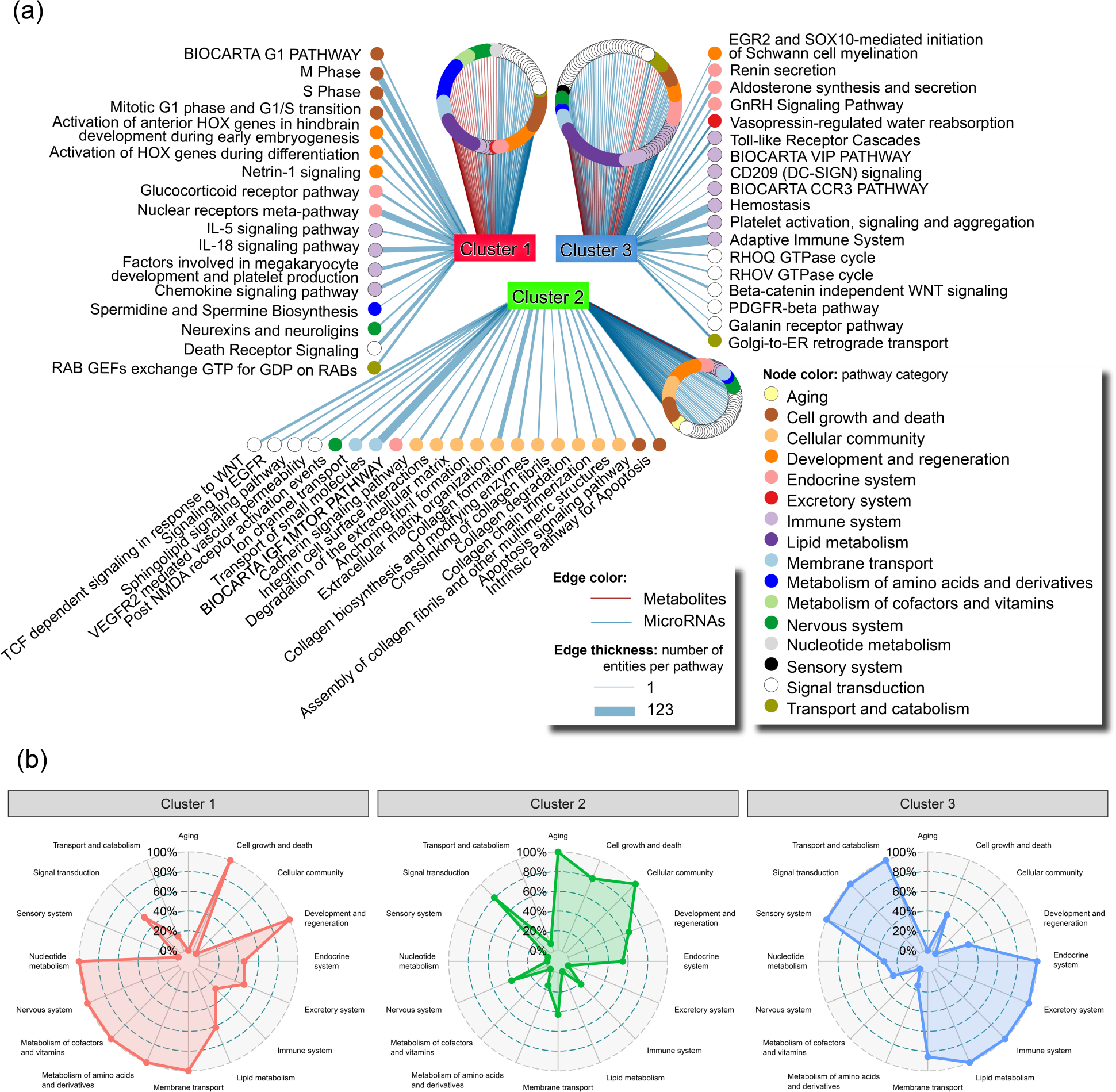
Cluster endotype signatures are associated with different physiological pathways. (a) Network depicting unique enriched pathways from miRNA-gene targets and annotated pathways from metabolites associated with individual cluster endotype signatures. Labels show pathways with lowest q-value or highest number of annotated genes. (b) Radar plots of pathway categorizations from enriched pathways from miRNA-gene targets and annotated pathways from metabolites indicating categories most associated to individual clusters endotypes.

We next used pathway categories to evaluate physiologically-relevant mechanisms linked to each endotype. Each enriched miRNA-derived pathway or annotated metabolite pathway was counted based on its category annotation in pathDIP, scaled and visualized using radar plots (Figure 3b). The cluster 1 endotype signature was most linked to pathway categories associated with development and regeneration, membrane transport, metabolism of various molecules, and the nervous system. The cluster 2 endotype signature was most linked to aging, and cellular community categories. Finally, the cluster 3 endotype signature was most linked to transport and catabolism, signal transduction, sensory system, endocrine system, excretory system, immune system, catabolism, and lipid metabolism categories. Overall, these analyses suggested that features associated with each endotype were uniquely associated with distinct physiological pathways.

### Evaluation of classification performance for WOMAC pain and function responses

To determine the classification performance of our clusters for identifying post-TKA WOMAC pain and function response status, we used an integrative ML framework using five domains— plasma metabolites, plasma miRNAs, synovial fluid miRNAs, urine miRNAs, and clinical data (age, sex, BMI, anxiety and depression categories, and neuropathic pain category; Figure 4a). Subject clusters had similar mean pain and function scores 1 year post-TKA, change in scores from baseline to 1 year, as well as pain and function response rates (Table 1). We first conducted differential expression analysis in each cluster, identifying metabolites with a fold-change of 1·1 and miRNAs within each biofluid with a fold change of 1·5 between responders and non-responders. Subsequently, we employed 10-fold cross-validation to estimate the AUC. Of the ML approaches compared, random forests consistently outperformed others in each individual domain (Supplementary Table 9) and was used for the unimodal ML models.

**Figure 4:**
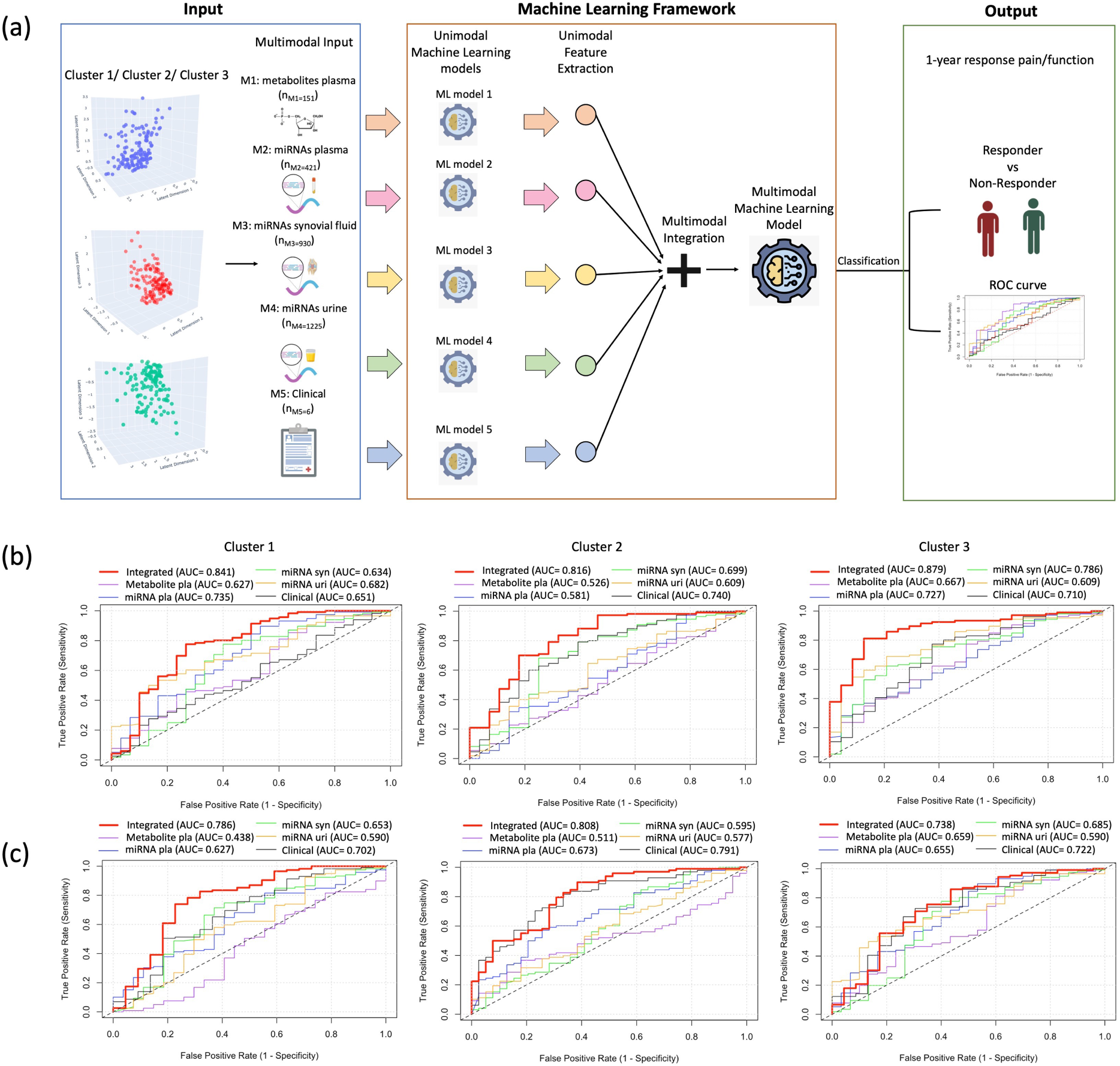
Machine Learning modeling for classifying response to 1 year pain and function within each cluster. (a) Comprehensive two-step machine learning (ML) framework wherein, initially, unimodal ML models extract features from each domain, including metabolites, miRNA from plasma, synovial fluid, urine, and clinical data. A second-level multimodal machine learning classifier integrates these features to efficiently classify response vs. non-response at 1 year. (b) Receiver Operating Characteristic (ROC) plots illustrating the Area Under the Curve (AUC) for individual domains, alongside the integrated AUC (in red) combining all five domains to classify WOMAC pain response. (c) Receiver Operating Characteristic (ROC) plots illustrating the Area Under the Curve (AUC) for individual domains, alongside the integrated AUC (in red) combining all five domains to classify WOMAC function response. Notably, the integrated AUC outperforms individual AUCs in Cluster 1, Cluster 2, and Cluster 3.

After differential analysis-based feature selection, cluster 1-3 retained 250, 87, and 49 features, respectively for the ML analysis to classify pain response (Supplementary Table 10). Initially, unimodal models were applied to each domain. Within cluster 1, miRNA plasma demonstrated the highest unimodal performance with an AUC of 0·735. However, the integrative performance, combining all 4 domains in the meta-classifier, notably improved AUC to 0·841 (highlighted in red in the ROC plot). For cluster 2, the clinical domain had the highest unimodal AUC of 0·740, while the integrative AUC was 0·816. Cluster 3 achieved the highest integrative AUC of 0·879, with miRNA urine showing the highest unimodal AUC of 0·786 (Figure 4b).

Based on differential analysis-based feature selection to classify function response, clusters 1-3 retained 63, 46, and 46 features, respectively, for the ML analysis (Supplementary Table 11). Across clusters 1-3, the clinical data domain consistently exhibited the best performance among unimodal domains, with AUCs of 0·702, 0·791, and 0·722, respectively. In terms of integrative performance, clusters 1-3 achieved AUCs of 0·786, 0·808, 0·738 (Figure 4c), respectively.

### Identifying key response classification features in our multimodal machine learning framework

To enhance the interpretability of our model we identified the most important features (molecular and clinical) contributing to response classification in each cluster. The top 20 features contributing to WOMAC pain or function response classification are shown in Supplementary Figures 4 & 5, respectively. Each top 20 list consisted of features from all 4 molecular domains, with a notable absence of clinical features; however, all molecular and clinical features inherently played a role in response classification (Supplementary Tables 12&13). Interestingly, only 3 miRNAs overlapped in the top 20 important features for WOMAC pain response between clusters 2 & 3, namely synovial fluid hsa-miR-1265 and hsa-mir-642a-3p, plasma hsa-3942-5p. In addition, only the metabolite glutamine overlapped in the top 20 important feature lists of clusters 2 & 3 for WOMAC function response. Thus, using our integrative approach, the vast majority of the most important features for response classification were unique for each cluster and were primarily driven by molecular entities. Overall, these findings highlight the diverse molecular features associated with outcome classification in each cluster, emphasizing the importance of integrating multiple domains for classification modeling of WOMAC pain and function responses post-TKA.

## Interpretation

Demographic, anthropometric and clinical characteristics of OA patients are heterogeneous, influencing outcomes to therapy, including TKA.^3,29^ Heterogeneity has also been identified through biofluid data, however, most studies to date have used single biofluids with single molecular type measures to identify endotypes within OA cohorts.^10,11^ We developed a novel multimodal deep learning algorithm, *omicVAE*, to cluster a sample of 414 KOA subjects who underwent TKA using preoperative miRNA and metabolite feature sets, identified by miRNomics and targeted metabolomics, from plasma, synovial fluid and urine, and uncovered three unique cluster endotypes. To our knowledge, our study is the first to use patient-matched multi-fluid, multi-omic approach to KOA patient endotyping. Not only did we uncover three unique multi-omic-based cluster endotypes, each was linked to unique biologically-relevant pathways. Despite a similar clinical phenotype, the cluster 1 endotype was primarily linked to metabolic processes and nervous system pathways, the cluster 2 endotype was primarily associated with aging pathways, and the cluster 3 endotype was primarily linked to immune, endocrine, and lipid metabolism pathways. Overall, the cluster endotypes uncovered are likely to contribute to, or be the result of, distinct mechanisms associated with KOA patients. Finally, using this novel approach to cluster endotyping, combined with integrative multimodal ML, we enhanced classification of patient-reported pain and function responses beyond that achieved using clinical measures alone. Surprisingly, it was the molecular entities that primarily drove classification of pain and function responses using our integrative modeling. Overall, our unique methodological approach reduced OA patient heterogeneity by defining patient clusters that had intra-cluster molecular differences that enhanced classifing pain and function responses to TKA. As endotypes are further refined and molecular entities best associated with classification are further characterized, it will be important to understand how these response-based signatures may relate to physiological responses post-surgery.

The strength of our methodology lies in developing integrative deep learning and ML techniques for efficient multi-omic endotyping and response classification in KOA patients. VAEs offer advantages to integrating multiple domains for clustering compared to traditional approaches by effectively capturing the underlying structure of heterogeneous data through a joint latent representation. While traditional approaches such as dimensionality reduction or sequential clustering may provide insights, they often suffer from limitations such as difficulty in capturing non-linear relationships, and inadequate integration of domain-specific characteristics. Unlike standard VAEs,^19^ we employed 4 separate decoders, enabling domain-specific reconstruction facilitating robust subject clustering, accounting for the inherent uncertainty in the latent space via the VAE’s probabilistic nature. Overall, VAEs effectively leverage complementary information from multiple modalities for a more comprehensive characterization of KOA patient endotypes. The integration of multiple data domains through our comprehensive two-step ML framework represents a significant advancement in response modeling for KOA outcomes by combining complementary information inherent in metabolomics, miRNA, and clinical data domains. Utilization of unimodal ML models in the first step allowed for extraction of domain-specific features that could classify 1-year TKA pain and function responses, while subsequent integration of these features using a naïve-Bayes meta-classifier enhanced classification accuracy. Naïve-Bayes classifiers emulate aspects of clinical decision-making by probabilistically combining evidence from multiple sources to make classifications.^30^ Importantly, our novel framework demonstrated improvements in classification performance compared to unimodal domain-specific models, underscoring the utility of an integrative approach.

Although we included three biofluids to integrate miRNA or metabolite features to identify endotypes among KOA patients, additional endotypes may exist. Incorporating additional omic technologies (e.g. proteomics, genomics, methylomics) in the presented framework, as well as comprehensively evaluating omic measures across all biofluids may further refine endotypes, or uncover additional endotypes to further improve our understanding of KOA and ability to more accurately classify responses to interventions. Future studies should also focus on easily obtained patient biofluids, such as urine and blood, to determine if the presented approach can show similar endotyping capability and response classification accuracy. In response classification modelling, we only evaluated a subset of clinical and demographic variables associated with KOA and patient outcomes to TKA, but incorporation of additional patient-related clinical and sociodemographic variables (e.g. comorbidities, medication use, race etc.), alongside endotype data, may also help improve modeling accuracy. Although we extensively validated our integrative ML unimodal models using a 10-time, 10-fold cross-validation, a lack of external validation remains. For external validation to be accomplished, better patient clinical and sociodemographic annotations, omic data and biosample sharing practices, and harmonization are needed.^7,8^ Lastly, similar evaluations in additional patient cohorts, such as those with early-KOA and other afflicted joints, or evaluating other response measures, would also be of interest moving forward.

Overall, using our novel modelling framework, we were able to unravel some heterogeneity of a sample of late-stage surgical KOA patients and evaluate post-TKA response classification. We anticipate this methodological approach will aid in understanding underlying molecular contributors and pathways to clusters of OA patients, and define molecular signatures contributing to intervention response. With additional studies, our methodological approach could ultimately help in shared patient-clinician decision making with regard to proceeding with selected therapies, including TKA for primary KOA.

## Data Availability

De-identified subject primary microRNA sequencing datasets are available on the Gene Expression Omnibus under accession number GSE222979. Software code and the dataset of processed miRNA counts, metabolite concentrations and demographic, anthropometric and clinical questionnaire responses used in this study is available at GitHub.

https://github.com/divya031090/DeepLearning_KOA

https://www.ncbi.nlm.nih.gov/geo/query/acc.cgi?acc=GSE222979

## Contributors

JSR, DS, OEG, KH, AS, YRR, AVP, RG and MK conceptualized the study. NNM, K. Syed, AVP, YRR, RG and MK supervised patient data and biofluid collection. KP managed biofluid and patient data storage. JSR, DS, OEG, KH, AS, CP, IJ, K. Sundararajan, YRR, AVP, RG and MK processed and curated the data. JSR, DS, OEG, KH, AS, CP, PP, NF, IJ and MK created the methodology and validated the data. DS developed the deep learning and machine learning algorithms. DS, OEG, and KH performed statistical analyses. JSR, DS, CP, IJ and MK created figures and tables. JSR, DS, KH, AS, CP and MK wrote the manuscript. MK supervised and acquired funding for multi-omic analysis. All authors had full access to the study data, were involved in manuscript editing, and were responsible for the decision to submit for publication.

## Data Sharing

De-identified subject primary microRNA sequencing datasets are available on the Gene Expression Omnibus under accession number GSE222979. Software code and the dataset of processed miRNA counts, metabolite concentrations and demographic, anthropometric and clinical questionnaire responses used in this study is available at https://github.com/divya031090/DeepLearning_KOA.

## Acknowledgments

Funding for this project was provided by the Canada Research Chairs Program (MK), Tony and Shari Fell Platinum Chair in Arthritis Research (MK), Campaign to Cure Arthritis, University Health Network Foundation. AVP is supported by the Arthritis Society Canada STAR Award-20-0000000012 and YRR is supported by J. Bernard Gosevitz Chair in Arthritis Research at University Health Network. Computational analysis was supported in part by funding from Natural Sciences and Engineering Research Council of Canada (NSERC RGPIN-2024-04314), Canada Foundation for Innovation (CFI #225404, #30865), and Ontario Research Funds (RDI #34876, RE010-020). The funders had no role in study design, data collection and analysis, decision to publish, or preparation of the manuscript. Authors would like to thank the clinical research team within the Division of Orthopedics and members of the Buchan Arthritis Center at the Schroeder Arthritis Institute for their assistance in study recruitment. We also thank Dr. Max Kotlyar for his assistance during initial discussions related to the study.

