## Supplemental Figures and Tables for "Deep Learning-Based Multimodal Clustering Model for Endotyping and Post-Arthroplasty Response Classification using Knee Osteoarthritis Subject-Matched Multi-Omic Data"

**Supplementary Figures and Tables**

**Contents**


Supplementary Figure 1……………………………………………………………………………Pg. 2
Supplementary Figure 2……………………………………………………………………………Pg. 3
Supplementary Figure 3……………………………………………………………………………Pg. 4
Supplementary Figure 4……………………………………………………………………………Pg. 5
Supplementary Figure 5……………………………………………………………………………Pg. 6
Supplementary Table 1...…………………………………………………………………………...Pg. 7
Supplementary Table 2...…………………………………………………………………………...Pg. 8
Supplementary Table 3...……………………………………………………………………………Pg. 10
Supplementary Table 4...……………………………………………………………………………Pg. 16
Supplementary Table 5...……………………………………………………………………………Pg. 22
Supplementary Table 6...……………………………………………………………………………Pg. 29
Supplementary Table 7...…………………………………………………………………...…...Pg. 31
Supplementary Table 8...…………………………………………………………………...…...Pg. 32
Supplementary Table 9...…………………………………………………………………...…...Pg. 35
Supplementary Table 10……………………………………………………………………………Pg. 36
Supplementary Table 11……………………………………………………………………………Pg. 42
Supplementary Table 12……………………………………………………………………………Pg. 45
Supplementary Table 13……………………………………………………………………………Pg. 51

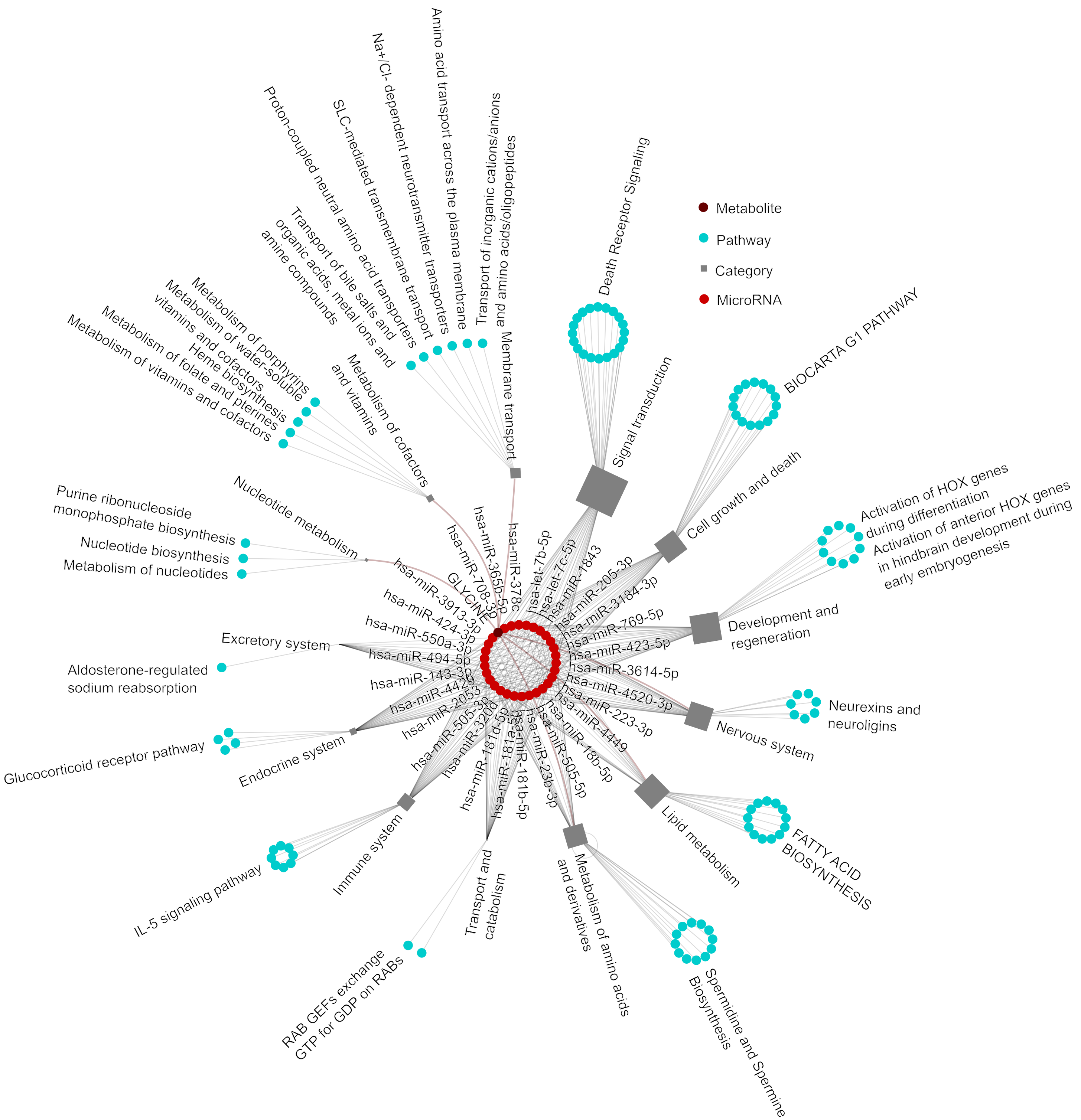


**Supplementary Figure 1. Network of unique pathways enriched and/or annotated for endotype signature of cluster 1.** Top pathways enriched for miRNA target genes identified by mirDIP and pathways annotated with metabolites are connected to corresponding pathway categories annotated in pathDIP, and further linked to cluster endotype signature features.

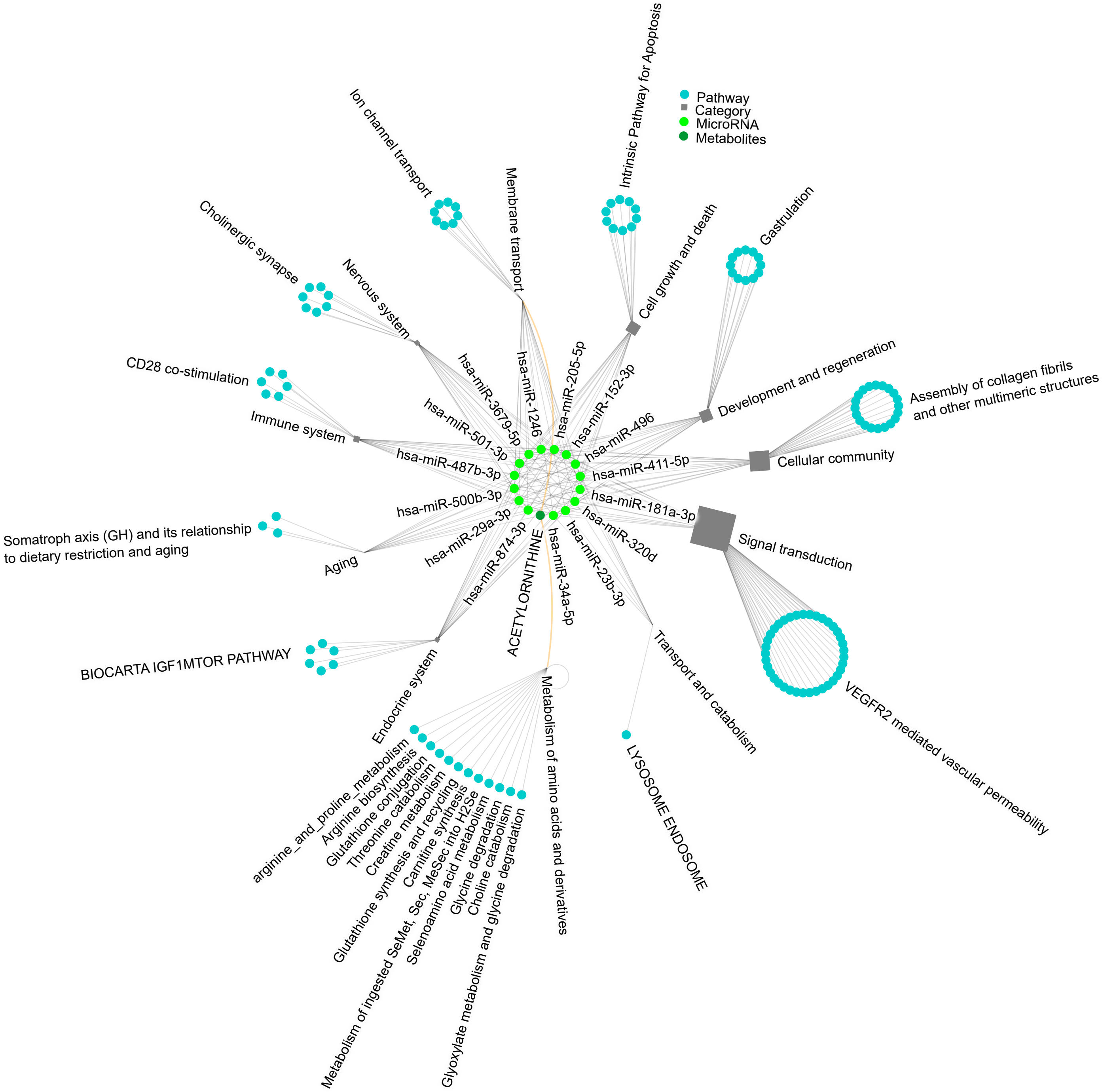


**Supplementary Figure 2. Network of unique pathways enriched and/or annotated for endotype signature of cluster 2.** Top pathways enriched for miRNA target genes identified by mirDIP and pathways annotated with metabolites are connected to corresponding pathway categories annotated in pathDIP, and further linked to cluster endotype signature features.

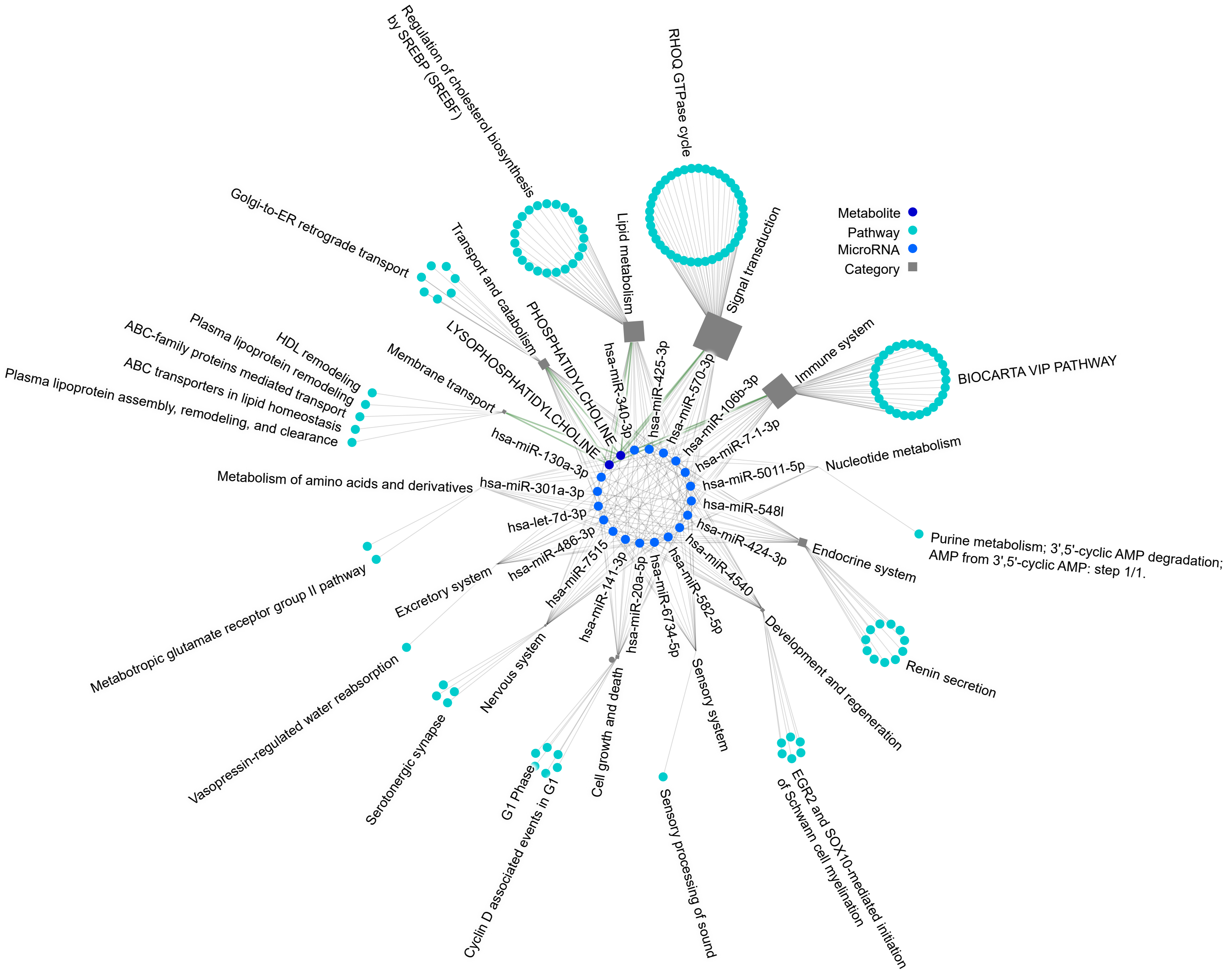


**Supplementary Figure 3. Network of unique pathways enriched and/or annotated for endotype signature of cluster 3.** Top pathways enriched for miRNA target genes identified by mirDIP and pathways annotated with metabolites are connected to corresponding pathway categories annotated in pathDIP, and further linked to cluster endotype signature features.

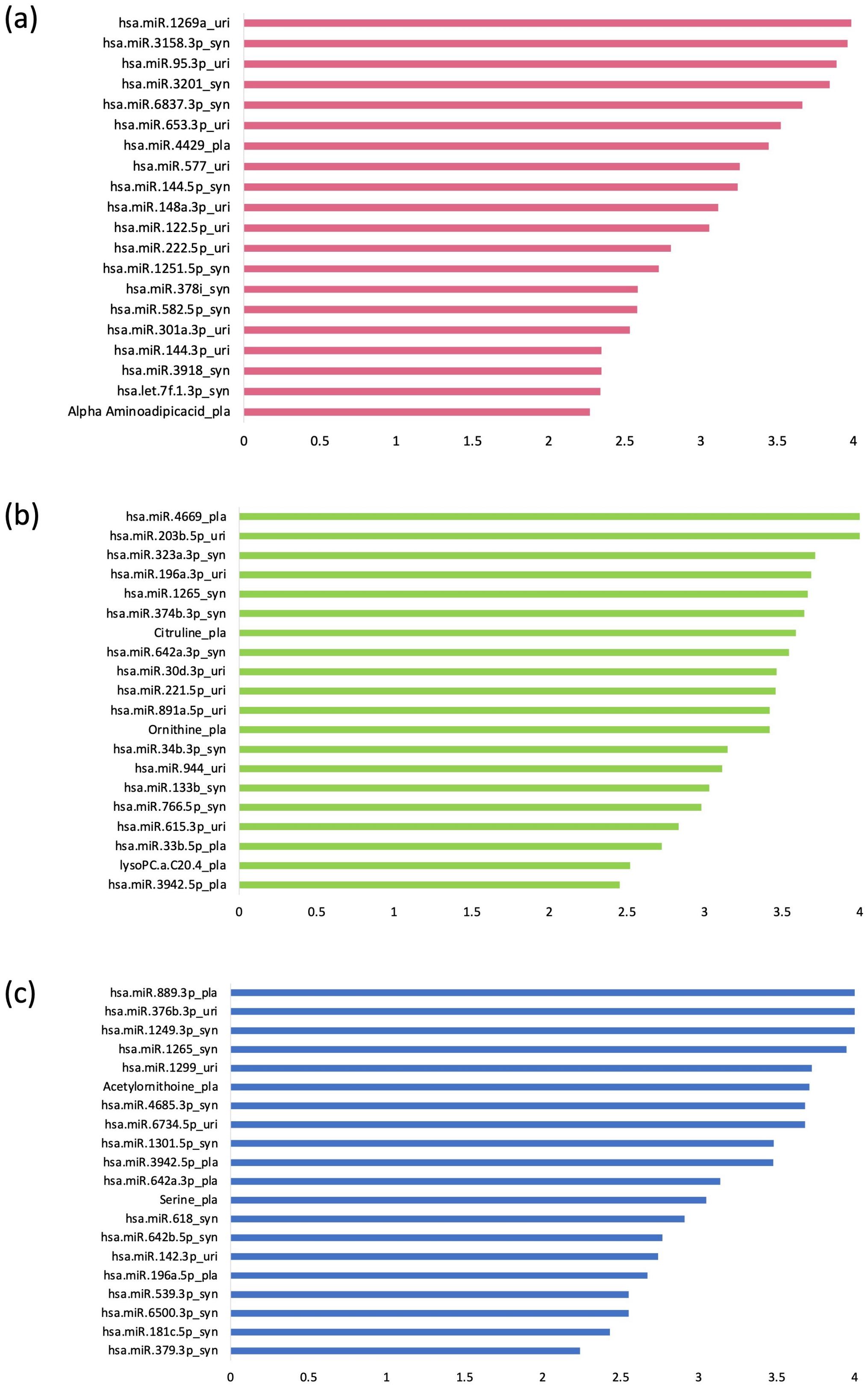


**Supplementary Figure 4. Cluster-wise feature importance plot obtained using integrative multimodal machine learning framework.** (a-c) mean Gini impurity-based feature importance for top-20 features across various domains, including clinical data, miRNA plasma, miRNA synovial, miRNA urine, and metabolite plasma, within the integrated model of WOMAC pain response classification for Cluster 1, 2 and 3, respectively.

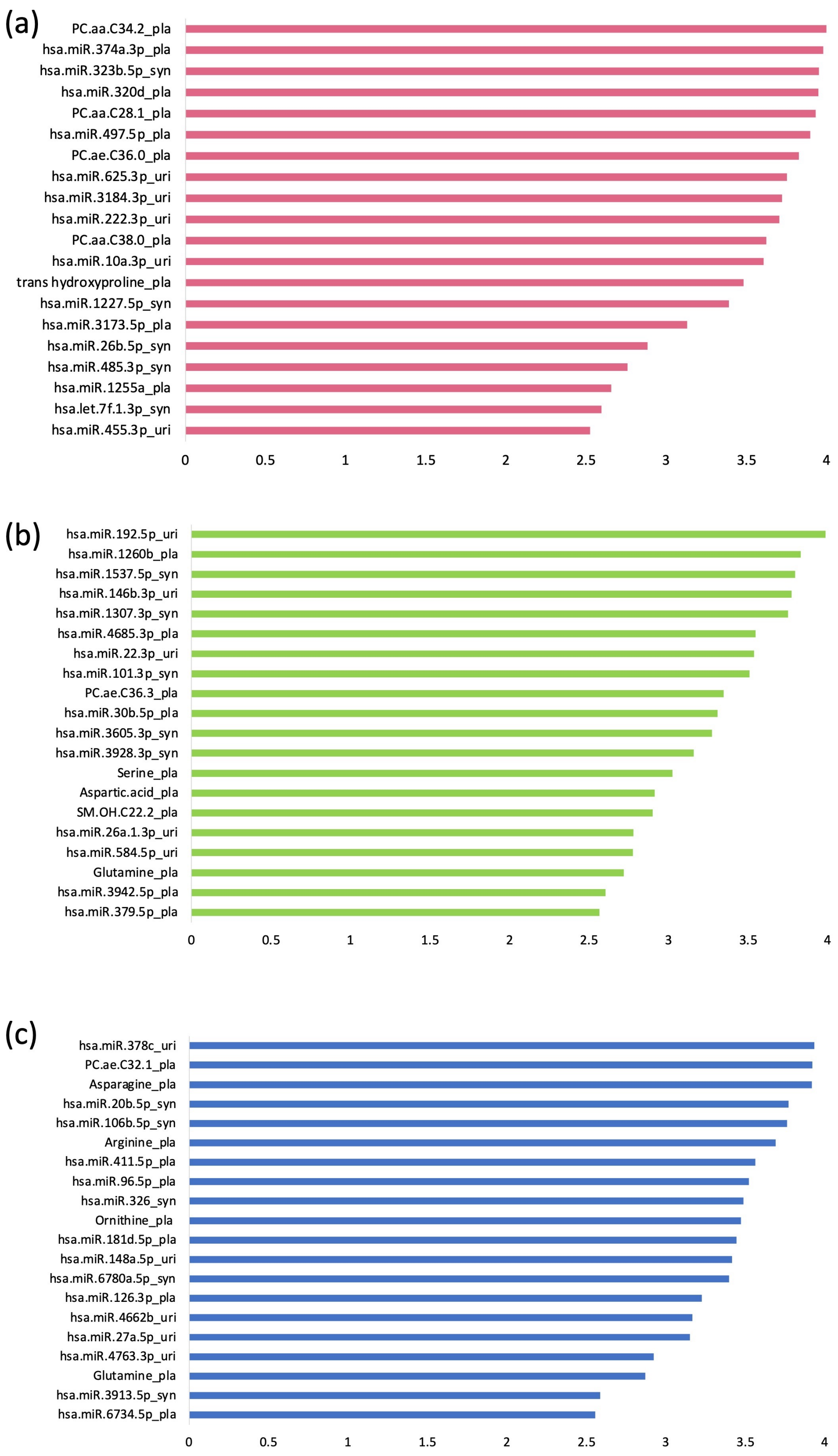


**Supplementary Figure 5. Cluster-wise feature importance plot obtained using integrative multimodal machine learning framework.** (a-c) mean Gini impurity-based feature importance for top-20 features across various domains, including clinical data, miRNA plasma, miRNA synovial, miRNA urine, and metabolite plasma, within the integrated model of WOMAC function response classification for Cluster 1, 2 and 3, respectively.

**Supplementary Table 1.** Summary statistics of N=414 TKR subjects at baseline.

| | Full Sample (n=414) |
|---|---|
| **Sex** | |
| Female [N, (%)] | 232 (56) |
| Male [N, (%)] | 182 (44) |
| **Age** | |
| Mean (sd) | 65.7 (8.4) |
| Median (Q1,Q3) | 66 (60, 71) |
| **BMI** | |
| Mean (sd) | 31.3 (7.0) |
| Median (Q1,Q3) | 29.8 (26.3, 35.0) |
| **HADS anxiety** | |
| normal [N, (%)] | 301 (74) |
| borderline [N, (%)] | 54 (13) |
| case [N, (%)] | 50 (12) |
| Missing (N) | 9 |
| **HADS depression** | |
| normal [N, (%)] | 324 (79) |
| borderline [N, (%)] | 48 (12) |
| case [N, (%)] | 37 (9) |
| Missing (N) | 5 |
| **painDETECT** | |
| neuropathic [N, (%)] | 55 (13) |
| nociceptive [N, (%)] | 255 (62) |
| unclear [N, (%)] | 81 (20) |
| Missing [N, (%)] | 23 (6) |
| **WOMAC pain baseline** | |
| Mean (sd) | 10.1 (3.5) |
| Median (Q1,Q3) | 10 (8, 12) |
| **WOMAC function baseline** | |
| Mean (sd) | 34.5 (11.9) |
| Median (Q1,Q3) | 35 (26, 42) |
| Missing (N) | 2 |

BMI, body mass index; HADS, hospital anxiety and depression scale; Q, quantile; sd, standard deviation.

**Supplementary Table 2. Signature Features for identified cluster endotypes.** Differential expression analysis and standardized mean difference were employed to identify significant features associated with each cluster showing both significant Standardized Mean Differences (SMD) and statistically significant differential expression (DE) within clusters [adjusted p-value (q-value) DE and q-value SMD <0.05]. The comparison includes Cluster 1 vs Clusters 2 and 3, Cluster 2 vs Clusters 1 and 3, and Cluster 3 vs Clusters 1 and 2. These signature features provide insights into the distinct molecular profiles characterizing different endotypes.

| **Cluster 1 vs 2 and 3** | | | | | |
|---|---|---|---|---|---|
| **Multiomics domain** | | **logFold Change** | **qvalue DE** | **SMD effect size** | **qvalue SMD** |
| **Metabolites plasma** | Glycine_pla | 0.105 | 0.019 | 0.213 | 0.039 |
| **miRNA plasma** | hsa-let-7b-5p_pla | 0.642 | 0.009 | 0.276 | 0.009 |
| | hsa-let-7c-5p_pla | 0.158 | 0.011 | 0.33 | 0.001 |
| | hsa-miR-143-3p_pla | 0.104 | 0.029 | 0.244 | 0.016 |
| | hsa-miR-1843_pla | 0.247 | 0.038 | 0.22 | 0.046 |
| | hsa-miR-3913-3p_pla | 0.241 | 0.042 | 0.216 | 0.045 |
| | hsa-miR-424-3p_pla | 0.39 | 0.002 | 0.266 | 0.028 |
| **miRNA synovial fluid** | hsa-miR-181a-5p _syn | 0.143 | 0.031 | 0.248 | 0.03 |
| | hsa-miR-181b-5p _syn | 0.168 | 0.024 | 0.281 | 0.022 |
| | hsa-miR-181d-5p _syn | 0.286 | 0.023 | 0.271 | 0.031 |
| | hsa-miR-18b-5p _syn | 0.298 | 0.029 | 0.279 | 0.025 |
| | hsa-miR-2053 _syn | 0.398 | 0.002 | 0.398 | <0.001 |
| | hsa-miR-223-3p _syn | 0.125 | 0.048 | 0.215 | 0.037 |
| | hsa-miR-3184-3p _syn | 0.168 | 0.025 | 0.22 | 0.033 |
| | hsa-miR-3614-5p _syn | 0.314 | 0.017 | 0.209 | 0.04 |
| | hsa-miR-365b-5p _syn | 0.263 | 0.039 | 0.227 | 0.032 |
| | hsa-miR-378c _syn | 0.262 | 0.041 | 0.21 | 0.038 |
| | hsa-miR-423-5p _syn | 0.158 | 0.008 | 0.25 | 0.015 |
| | hsa-miR-4429 _syn | 0.376 | 0.03 | 0.209 | 0.041 |
| | hsa-miR-4520-3p _syn | 0.338 | 0.016 | 0.254 | 0.013 |
| | hsa-miR-494-5p _syn | 0.295 | 0.039 | 0.249 | 0.011 |
| | hsa-miR-505-3p _syn | 0.326 | 0.006 | 0.255 | 0.015 |
| | hsa-miR-505-5p _syn | 0.291 | 0.009 | 0.267 | 0.008 |
| | hsa-miR-550a-3p _syn | 0.352 | 0.015 | 0.204 | 0.045 |
| **miRNA urine** | hsa-miR-205-3p_uri | 0.391 | 0.021 | 0.219 | 0.041 |
| | hsa-miR-23b-3p_uri | 0.152 | 0.008 | 0.28 | 0.034 |
| | hsa-miR-320d_uri | 0.295 | 0.022 | 0.201 | 0.049 |
| | hsa-miR-4449_uri | 0.282 | 0.023 | 0.291 | 0.032 |
| | hsa-miR-708-3p_uri | 0.243 | 0.046 | 0.217 | 0.041 |
| | hsa-miR-769-5p_uri | 0.156 | 0.017 | 0.213 | 0.043 |
| **Cluster 2 vs 1 and 3** | | | | | |
| **Multiomics domain** | | **logFold Change** | **qvalue DE** | **SMD effect size** | **qvalue SMD** |
| **Metabolites plasma** | Acetylornithoine_pla | 0.199 | 0.025 | 0.249 | 0.017 |
| **miRNA plasma** | hsa-miR-1246_pla | 0.26 | 0.043 | 0.22 | 0.039 |
| | hsa-miR-152-3p_pla | 0.131 | 0.025 | 0.214 | 0.041 |
| | hsa-miR-181a-3p_pla | 0.294 | 0.009 | 0.216 | 0.047 |
| | hsa-miR-205-5p_pla | 0.319 | 0.002 | 0.274 | 0.013 |
| | hsa-miR-23b-3p_pla | 0.098 | 0.037 | 0.277 | 0.012 |
| | hsa-miR-29a-3p_pla | 0.197 | 0.001 | 0.292 | 0.005 |
| | hsa-miR-320d_pla | 0.295 | 0.025 | 0.258 | 0.033 |
| | hsa-miR-34a-5p_pla | 0.229 | 0.018 | 0.217 | 0.044 |
| | hsa-miR-3679-5p_pla | 0.3 | 0.025 | 0.228 | 0.039 |
| | hsa-miR-874-3p_pla | 0.282 | 0.027 | 0.257 | 0.036 |
| **miRNA synovial fluid** | hsa-miR-411-5p _syn | 0.209 | 0.023 | 0.293 | 0.021 |
| | hsa-miR-487b-3p _syn | 0.303 | 0.01 | 0.215 | 0.048 |
| | hsa-miR-496 _syn | 0.331 | 0.017 | 0.338 | 0.001 |
| **miRNA urine** | hsa-miR-500b-3p_uri | 0.302 | 0.02 | 0.214 | 0.049 |
| | hsa-miR-501-3p_uri | 0.187 | 0.044 | 0.239 | 0.035 |
| **Cluster 3 vs 1 and 2** | | | | | |
| **Multiomics domain** | | **logFold Change** | **qvalue DE** | **SMD effect size** | **qvalue SMD** |
| **Metabolites plasma** | lysoPC.a.C16.0_pla | 0.06 | 0.049 | 0.274 | 0.011 |
| | lysoPC.a.C17.0_pla | 0.061 | 0.028 | 0.222 | 0.035 |
| | PC.ae.C34.0_pla | 0.047 | 0.034 | 0.269 | 0.015 |
| | PC.ae.C42.3_pla | 0.035 | 0.04 | 0.269 | 0.016 |
| | PC.ae.C42.4_pla | 0.042 | 0.037 | 0.273 | 0.012 |
| **miRNA plasma** | hsa-let-7d-3p_pla | 0.092 | 0.014 | 0.23 | 0.03 |
| | hsa-miR-106b-3p_pla | 0.096 | 0.026 | 0.229 | 0.047 |
| | hsa-miR-130a-3p_pla | 0.098 | 0.039 | 0.238 | 0.035 |
| | hsa-miR-20a-5p_pla | 0.092 | 0.028 | 0.263 | 0.02 |
| | hsa-miR-301a-3p_pla | 0.138 | 0.036 | 0.265 | 0.017 |
| | hsa-miR-340-3p_pla | 0.304 | 0.009 | 0.243 | 0.026 |

| | | | | | |
|---|---|---|---|---|---|
| | hsa-miR-425-3p_pla | 0.248 | 0.028 | 0.249 | 0.027 |
| | hsa-miR-486-3p_pla | 0.091 | 0.035 | 0.268 | 0.015 |
| | hsa-miR-548l_pla | 0.266 | 0.045 | 0.242 | 0.025 |
| | hsa-miR-570-3p_pla | 0.419 | 0.003 | 0.289 | 0.005 |
| | hsa-miR-6734-5p_pla | 0.305 | 0.004 | 0.317 | 0.006 |
| | hsa-miR-7.1-3p_pla | 0.283 | 0.028 | 0.262 | 0.017 |
| **miRNA synovial fluid** | hsa-miR-141-3p_syn | 0.137 | 0.018 | 0.221 | 0.046 |
| | hsa-miR-4540_syn | 0.235 | 0.016 | 0.285 | 0.007 |
| | hsa-miR-5011-5p_syn | 0.285 | 0.032 | 0.264 | 0.011 |
| | hsa-miR-582-5p_syn | 0.324 | 0.023 | 0.306 | 0.004 |
| | hsa-miR-7515_syn | 0.112 | 0.044 | 0.283 | 0.001 |
| **miRNA urine** | hsa-miR-301a-3p _uri | 0.377 | 0.01 | 0.249 | 0.023 |
| | hsa-miR-424-3p _uri | 0.243 | 0.033 | 0.275 | 0.01 |

**Supplementary Table 3. Pathway enrichment analysis of cluster 1 endotype putative microRNA gene targets.** Gene targets of the microRNAs specific for each cluster were obtained from mirDIP and used to perform pathway enrichment analysis using pathDIP.

| Pathway Source | Pathway Name | q-value (BH method) | Genes mapped | Category |
|---|---|---|---|---|
| **Cluster 1** | | | | |
| REACTOME | Signal Transduction | 1.18E-43 | 631 | Signal transduction |
| ACSN2 | PI3K AKT MTOR | 5.54E-16 | 107 | Signal transduction |
| REACTOME | Signaling by Receptor Tyrosine Kinases | 1.55E-15 | 155 | Signal transduction |
| KEGG | MAPK signaling pathway | 1.34E-14 | 101 | Signal transduction |
| REACTOME | Signaling by Rho GTPases, Miro GTPases and RHOBTB3 | 2.06E-14 | 186 | Signal transduction |
| ACSN2 | EMT REGULATORS | 4.20E-14 | 167 | Signal transduction |
| REACTOME | Signaling by Rho GTPases | 6.04E-14 | 181 | Signal transduction |
| REACTOME | Nervous system development | 2.94E-13 | 160 | Development and regeneration |
| REACTOME | Axon guidance | 5.38E-13 | 154 | Development and regeneration |
| REACTOME | Membrane Trafficking | 8.10E-13 | 169 | Transport and catabolism |
| KEGG | Axon guidance | 1.22E-12 | 70 | Development and regeneration |
| REACTOME | Developmental Biology | 3.74E-12 | 262 | Development and regeneration |
| REACTOME | RHO GTPase cycle | 5.28E-12 | 129 | Signal transduction |
| WikiPathways | Brain-derived neurotrophic factor (BDNF) signaling pathway | 5.36E-12 | 59 | Nervous system |
| WikiPathways | MAPK signaling pathway | 2.16E-11 | 82 | Signal transduction |
| WikiPathways | Focal adhesion: PI3K-Akt-mTOR-signaling pathway | 3.66E-11 | 95 | Cellular community |
| REACTOME | Intracellular signaling by second messengers | 3.67E-11 | 96 | Signal transduction |
| WikiPathways | EGF/EGFR signaling pathway | 3.68E-11 | 62 | Signal transduction |
| ACSN2 | STARVATION AUTOPHAGY | 6.26E-11 | 62 | Transport and catabolism |
| KEGG | Cellular senescence | 8.06E-11 | 59 | Cell growth and death |
| KEGG | PI3K-Akt signaling pathway | 1.26E-10 | 104 | Signal transduction |
| KEGG | Ras signaling pathway | 2.31E-10 | 77 | Signal transduction |
| KEGG | Autophagy - animal | 2.38E-10 | 54 | Transport and catabolism |
| REACTOME | MAPK family signaling cascades | 6.79E-10 | 96 | Signal transduction |
| ACSN2 | HEDGEHOG | 7.81E-10 | 88 | Signal transduction |
| WikiPathways | Mesodermal commitment pathway | 7.96E-10 | 57 | Development and regeneration |
| ACSN2 | DEATH RECEPTOR PATHWAYS | 1.75E-09 | 80 | Signal transduction |
| ACSN2 | MAPK | 1.96E-09 | 68 | Signal transduction |
| ACSN2 | MOMP REGULATION | 2.84E-09 | 59 | Cell growth and death |
| WikiPathways | Ras signaling | 2.88E-09 | 63 | Signal transduction |
| ACSN2 | WNT NON CANONICAL | 2.96E-09 | 114 | Signal transduction |
| ACSN2 | RCD GENES | 3.12E-09 | 51 | Cell growth and death |
| REACTOME | PIP3 activates AKT signaling | 7.02E-09 | 81 | Signal transduction |
| WikiPathways | PI3K-Akt signaling pathway | 7.73E-09 | 96 | Signal transduction |
| ACSN2 | APOPTOSIS | 7.86E-09 | 72 | Cell growth and death |
| WikiPathways | Focal adhesion | 1.21E-08 | 65 | Cellular community |
| REACTOME | Neuronal System | 1.38E-08 | 109 | Nervous system |
| REACTOME | Vesicle-mediated transport | 1.38E-08 | 177 | Transport and catabolism |
| KEGG | Focal adhesion | 2.28E-08 | 65 | Cellular community |
| WikiPathways | TGF-beta signaling pathway | 5.70E-08 | 48 | Signal transduction |
| WikiPathways | Neurogenesis regulation in the olfactory epithelium | 6.83E-08 | 28 | Sensory system |
| KEGG | FoxO signaling pathway | 7.52E-08 | 47 | Signal transduction |
| REACTOME | ESR-mediated signaling | 7.80E-08 | 61 | Signal transduction |
| WikiPathways | Ectoderm differentiation | 8.45E-08 | 50 | Development and regeneration |
| REACTOME | Signaling by TGFB family members | 9.76E-08 | 45 | Signal transduction |
| ACSN2 | IMMUNOSTIMULATORY CORE PATHWAYS | 9.83E-08 | 40 | Immune system |
| Panther_Pathway | Wnt signaling pathway | 9.90E-08 | 83 | Signal transduction |
| WikiPathways | Insulin signaling | 1.02E-07 | 54 | Endocrine system |
| KEGG | Neurotrophin signaling pathway | 1.27E-07 | 44 | Nervous system |
| WikiPathways | EGFR tyrosine kinase inhibitor resistance | 1.33E-07 | 35 | Signal transduction |
| KEGG | Apelin signaling pathway | 1.83E-07 | 48 | Signal transduction |
| Panther_Pathway | PDGF signaling pathway | 2.06E-07 | 45 | Signal transduction |
| KEGG | Rap1 signaling pathway | 2.34E-07 | 65 | Signal transduction |
| WikiPathways | VEGFA-VEGFR2 signaling | 2.37E-07 | 110 | Signal transduction |
| REACTOME | MAPK1/MAPK3 signaling | 2.56E-07 | 80 | Signal transduction |
| ACSN2 | SENESCENCE | 3.78E-07 | 36 | Cell growth and death |
| REACTOME | Signaling by Nuclear Receptors | 5.89E-07 | 75 | Signal transduction |
| WikiPathways | Endochondral ossification | 7.71E-07 | 28 | Development and regeneration |
| KEGG | Signaling pathways regulating pluripotency of stem cells | 7.83E-07 | 48 | Cellular community |
| WikiPathways | Endoderm differentiation | 7.83E-07 | 48 | Development and regeneration |
| KEGG | Endocytosis | 7.98E-07 | 71 | Transport and catabolism |
| Panther_Pathway | EGF receptor signaling pathway | 8.60E-07 | 41 | Signal transduction |

| | | | | |
|---|---|---|---|---|
| REACTOME | RAF/MAP kinase cascade | 9.42E-07 | 77 | Signal transduction |
| Panther_Pathway | CCKR signaling map | 1.03E-06 | 54 | Signal transduction |
| REACTOME | Cell Cycle, Mitotic | 1.14E-06 | 126 | Cell growth and death |
| REACTOME | Protein-protein interactions at synapses | 1.19E-06 | 33 | Nervous system |
| WikiPathways | ErbB signaling pathway | 1.58E-06 | 35 | Signal transduction |
| KEGG | Thyroid hormone signaling pathway | 1.66E-06 | 42 | Endocrine system |
| REACTOME | MET activates RAP1 and RAC1 | 1.69E-06 | 10 | Signal transduction |
| REACTOME | Signaling by NTRKs | 1.69E-06 | 45 | Signal transduction |
| REACTOME | MET promotes cell motility | 1.76E-06 | 21 | Signal transduction |
| ACSN2 | WNT CANONICAL | 3.08E-06 | 104 | Signal transduction |
| ACSN2 | CORE | 3.10E-06 | 28 | Signal transduction |
| WikiPathways | Hippocampal synaptogenesis and neurogenesis | 4.51E-06 | 15 | Signal transduction |
| REACTOME | Cell Cycle | 4.57E-06 | 148 | Cell growth and death |
| REACTOME | Rab regulation of trafficking | 5.92E-06 | 41 | Transport and catabolism |
| REACTOME | Signaling by TGF-beta Receptor Complex | 6.53E-06 | 34 | Signal transduction |
| KEGG | ErbB signaling pathway | 6.60E-06 | 32 | Signal transduction |
| KEGG | cGMP-PKG signaling pathway | 6.60E-06 | 51 | Signal transduction |
| WikiPathways | Clock-controlled autophagy in bone metabolism | 6.67E-06 | 31 | Transport and catabolism |
| WikiPathways | Hepatocyte growth factor receptor signaling | 7.26E-06 | 18 | Signal transduction |
| KEGG | TGF-beta signaling pathway | 8.27E-06 | 34 | Signal transduction |
| REACTOME | Signaling by NTRK1 (TRKA) | 8.29E-06 | 39 | Signal transduction |
| WikiPathways | TGF-beta receptor signaling | 8.35E-06 | 24 | Signal transduction |
| WikiPathways | Thermogenesis | 8.51E-06 | 38 | Thermogenesis |
| REACTOME | CDC42 GTPase cycle | 8.52E-06 | 48 | Signal transduction |
| WikiPathways | Gastrin signaling pathway | 1.01E-05 | 39 | Endocrine system |
| REACTOME | RAC1 GTPase cycle | 1.09E-05 | 54 | Signal transduction |
| REACTOME | Neurexins and neuroligins | 1.32E-05 | 23 | Nervous system |
| REACTOME | RND1 GTPase cycle | 1.35E-05 | 20 | Signal transduction |
| REACTOME | RAC2 GTPase cycle | 1.46E-05 | 32 | Signal transduction |
| REACTOME | RHOD GTPase cycle | 1.52E-05 | 23 | Signal transduction |
| REACTOME | Estrogen-dependent gene expression | 1.56E-05 | 39 | Signal transduction |
| WikiPathways | Adipogenesis | 1.84E-05 | 41 | Development and regeneration |
| REACTOME | Transmission across Chemical Synapses | 1.84E-05 | 71 | Nervous system |
| KEGG | Longevity regulating pathway | 1.86E-05 | 32 | Aging |
| BioCarta | BIOCARTA MET PATHWAY | 2.18E-05 | 17 | Development and regeneration |
| WikiPathways | BDNF-TrkB signaling | 2.18E-05 | 17 | Nervous system |
| Panther_Pathway | Integrin signalling pathway | 2.22E-05 | 49 | Signal transduction |
| REACTOME | RAC3 GTPase cycle | 2.24E-05 | 33 | Signal transduction |
| WikiPathways | PI3K-AKT-mTOR signaling pathway and therapeutic opportunities | 2.39E-05 | 16 | Signal transduction |
| WikiPathways | DYRK1A | 2.40E-05 | 25 | Development and regeneration |
| KEGG | Cell cycle | 2.54E-05 | 40 | Cell growth and death |
| REACTOME | RHOC GTPase cycle | 2.55E-05 | 28 | Signal transduction |
| REACTOME | RHOG GTPase cycle | 2.55E-05 | 28 | Signal transduction |
| WikiPathways | Cell cycle | 2.78E-05 | 39 | Cell growth and death |
| WikiPathways | Development of ureteric collection system | 4.33E-05 | 24 | Development and regeneration |
| REACTOME | Semaphorin interactions | 4.45E-05 | 25 | Development and regeneration |
| KEGG | Calcium signaling pathway | 4.45E-05 | 64 | Signal transduction |
| REACTOME | RHOBTB GTPase Cycle | 5.38E-05 | 17 | Signal transduction |
| KEGG | cAMP signaling pathway | 5.61E-05 | 59 | Signal transduction |
| KEGG | Oxytocin signaling pathway | 6.04E-05 | 46 | Endocrine system |
| WikiPathways | Embryonic stem cell pluripotency pathways | 6.19E-05 | 37 | Cellular community |
| REACTOME | RHOA GTPase cycle | 7.64E-05 | 44 | Signal transduction |
| WikiPathways | IL6 signaling pathway | 7.76E-05 | 19 | Immune system |
| REACTOME | RHOB GTPase cycle | 7.82E-05 | 26 | Signal transduction |
| WikiPathways | Glucocorticoid receptor pathway | 7.82E-05 | 26 | Endocrine system |
| Panther_Pathway | FGF signaling pathway | 7.93E-05 | 34 | Signal transduction |
| WikiPathways | Factors and pathways affecting insulin-like growth factor (IGF1)-Akt signaling | 8.04E-05 | 17 | Endocrine system |
| WikiPathways | Target of rapamycin signaling | 8.04E-05 | 17 | Signal transduction |
| REACTOME | RHOBTB1 GTPase cycle | 8.51E-05 | 13 | Signal transduction |
| KEGG | Insulin signaling pathway | 8.52E-05 | 41 | Endocrine system |
| WikiPathways | Hedgehog signaling pathway | 8.53E-05 | 20 | Signal transduction |
| ACSN2 | G2 M CHECKPOINT | 9.14E-05 | 28 | Cell growth and death |
| REACTOME | EPH-Ephrin signaling | 9.74E-05 | 31 | Development and regeneration |
| KEGG | Wnt signaling pathway | 9.89E-05 | 47 | Signal transduction |
| REACTOME | TBC/RABGAPs | 0.000109 | 19 | Transport and catabolism |
| REACTOME | Estrogen-dependent nuclear events downstream of ESR-membrane signaling | 0.000151 | 13 | Signal transduction |
| BioCarta | BIOCARTA MAPK PATHWAY | 0.000152 | 28 | Signal transduction |
| REACTOME | Signalling to ERKs | 0.000152 | 16 | Signal transduction |

| | | | | |
|---|---|---|---|---|
| REACTOME | RHO GTPases activate PAKs | 0.000159 | 12 | Signal transduction |
| REACTOME | Antigen processing: Ubiquitination & Proteasome degradation | 0.000161 | 75 | Immune system |
| WikiPathways | CKAP4 signaling pathway map | 0.000178 | 36 | Signal transduction |
| PathBank | Spermidine and Spermine Biosynthesis | 0.00018 | 6 | Metabolism of amino acids and derivatives |
| PharmGKB | VEGF Signaling Pathway | 0.00018 | 23 | Signal transduction |
| REACTOME | Clathrin-mediated endocytosis | 0.000181 | 42 | Transport and catabolism |
| REACTOME | Negative regulation of the PI3K/AKT network | 0.000194 | 35 | Signal transduction |
| REACTOME | Activation of HOX genes during differentiation | 0.000203 | 30 | Development and regeneration |
| REACTOME | Activation of anterior HOX genes in hindbrain development during early embryogenesis | 0.000203 | 30 | Development and regeneration |
| ACSN2 | ECM | 0.000204 | 38 | Cellular community |
| Panther_Pathway | Angiogenesis | 0.000227 | 44 | Development and regeneration |
| REACTOME | Netrin-1 signaling | 0.000228 | 20 | Development and regeneration |
| REACTOME | Cytokine Signaling in Immune system | 0.000233 | 149 | Immune system |
| BioCarta | BIOCARTA CTCF PATHWAY | 0.000244 | 13 | Immune system |
| Panther_Pathway | p53 pathway | 0.000247 | 27 | Cell growth and death |
| REACTOME | Signaling by MET | 0.000247 | 27 | Signal transduction |
| KEGG | AMPK signaling pathway | 0.000248 | 36 | Signal transduction |
| WikiPathways | Neural crest differentiation | 0.000249 | 32 | Development and regeneration |
| KEGG | Parathyroid hormone synthesis, secretion and action | 0.00025 | 34 | Endocrine system |
| WikiPathways | B cell receptor signaling pathway | 0.000273 | 31 | Immune system |
| KEGG | Adherens junction | 0.000273 | 25 | Cellular community |
| REACTOME | VEGFR2 mediated cell proliferation | 0.000278 | 11 | Signal transduction |
| WikiPathways | Nuclear receptors meta-pathway | 0.000286 | 75 | Endocrine system |
| ACSN2 | CASPASES | 0.000288 | 43 | Cell growth and death |
| REACTOME | Non-integrin membrane-ECM interactions | 0.000297 | 22 | Cellular community |
| BioCarta | BIOCARTA G1 PATHWAY | 0.00032 | 14 | Cell growth and death |
| REACTOME | L1CAM interactions | 0.000344 | 36 | Development and regeneration |
| WikiPathways | IL-5 signaling pathway | 0.000349 | 17 | Immune system |
| KEGG | Mitophagy - animal | 0.000362 | 24 | Transport and catabolism |
| BioCarta | BIOCARTA TCR PATHWAY | 0.000373 | 18 | Immune system |
| ACSN2 | TCR SIGNALING | 0.000375 | 52 | Immune system |
| KEGG | Phospholipase D signaling pathway | 0.000376 | 42 | Signal transduction |
| WikiPathways | White fat cell differentiation | 0.000385 | 14 | Immune system |
| REACTOME | Death Receptor Signaling | 0.000386 | 43 | Signal transduction |
| REACTOME | RAB GEFs exchange GTP for GDP on RABs | 0.000389 | 29 | Transport and catabolism |
| KEGG | Hippo signaling pathway | 0.000446 | 43 | Signal transduction |
| REACTOME | Prolonged ERK activation events | 0.000475 | 9 | Signal transduction |
| REACTOME | Signaling by NOTCH1 | 0.000544 | 25 | Signal transduction |
| ACSN2 | G1 S CHECKPOINT | 0.000571 | 15 | Cell growth and death |
| ACSN2 | GROWTH FACTORS SIGNALING PATHWAYS | 0.000571 | 15 | Signal transduction |
| KEGG | Prolactin signaling pathway | 0.000577 | 24 | Endocrine system |
| REACTOME | Syndecan interactions | 0.000588 | 13 | Cellular community |
| REACTOME | Signaling by VEGF | 0.000628 | 32 | Signal transduction |
| REACTOME | VEGFA-VEGFR2 Pathway | 0.000644 | 30 | Signal transduction |
| REACTOME | Extra-nuclear estrogen signaling | 0.000675 | 25 | Signal transduction |
| REACTOME | RHO GTPase Effectors | 0.000676 | 69 | Signal transduction |
| WikiPathways | Integrin-mediated cell adhesion | 0.000695 | 31 | Cellular community |
| WikiPathways | Thyroid hormones production and peripheral downstream signaling effects | 0.0007 | 29 | Metabolism of amino acids and derivatives |
| WikiPathways | Angiogenesis | 0.0007 | 12 | Development and regeneration |
| REACTOME | Cellular Senescence | 0.000794 | 44 | Cell growth and death |
| WikiPathways | Chemokine signaling pathway | 0.000794 | 44 | Immune system |
| Panther_Pathway | Insulin/IGF pathway-protein kinase B signaling cascade | 0.000801 | 15 | Endocrine system |
| REACTOME | Frs2-mediated activation | 0.000823 | 8 | Signal transduction |
| WikiPathways | Bone morphogenic protein signaling and regulation | 0.000823 | 8 | Signal transduction |
| WikiPathways | Leptin signaling pathway | 0.000825 | 25 | Signal transduction |
| WikiPathways | Prolactin signaling pathway | 0.000825 | 25 | Endocrine system |
| Panther_Pathway | B cell activation | 0.000836 | 21 | Immune system |
| REACTOME | NRAGE signals death through JNK | 0.000836 | 21 | Signal transduction |
| KEGG | Growth hormone synthesis, secretion and action | 0.000845 | 35 | Endocrine system |
| REACTOME | Intra-Golgi and retrograde Golgi-to-ER traffic | 0.000848 | 52 | Transport and catabolism |
| KEGG | Hedgehog signaling pathway | 0.000852 | 20 | Signal transduction |
| REACTOME | RHOJ GTPase cycle | 0.000852 | 20 | Signal transduction |
| REACTOME | MET activates PTPN11 | 0.000854 | 5 | Signal transduction |
| BioCarta | BIOCARTA CALCINEURIN PATHWAY | 0.000858 | 10 | Development and regeneration |
| PathBank | T Cell Receptor Signaling Pathway | 0.000867 | 18 | Immune system |

| | | | | |
|---|---|---|---|---|
| WikiPathways | Heart development | 0.000867 | 18 | Development and regeneration |
| REACTOME | Signaling by Activin | 0.00087 | 9 | Signal transduction |
| KEGG | Relaxin signaling pathway | 0.000873 | 37 | Endocrine system |
| KEGG | Oocyte meiosis | 0.000996 | 36 | Cell growth and death |
| WikiPathways | Disruption of postsynaptic signaling by CNV | 0.001075 | 13 | Nervous system |
| WikiPathways | Netrin-UNC5B signaling pathway | 0.001129 | 19 | Cell growth and death |
| REACTOME | Signaling by WNT | 0.001137 | 69 | Signal transduction |
| WikiPathways | Energy metabolism | 0.001141 | 18 | Energy metabolism |
| REACTOME | M Phase | 0.001146 | 85 | Cell growth and death |
| REACTOME | Signaling by FGFR | 0.001185 | 27 | Signal transduction |
| WikiPathways | G1 to S cell cycle control | 0.001215 | 22 | Cell growth and death |
| WikiPathways | Hippo signaling regulation pathways | 0.00123 | 30 | Signal transduction |
| REACTOME | PI5P, PP2A and IER3 Regulate PI3K/AKT Signaling | 0.0013 | 31 | Signal transduction |
| REACTOME | Neurotransmitter receptors and postsynaptic signal transmission | 0.001317 | 51 | Nervous system |
| WikiPathways | IL-18 signaling pathway | 0.001318 | 64 | Immune system |
| REACTOME | Cohesin Loading onto Chromatin | 0.001348 | 7 | Cell growth and death |
| REACTOME | MET receptor recycling | 0.001348 | 7 | Signal transduction |
| KEGG | Estrogen signaling pathway | 0.001368 | 38 | Endocrine system |
| REACTOME | Factors involved in megakaryocyte development and platelet production | 0.001405 | 42 | Immune system |
| BioCarta | BIOCARTA TGFB PATHWAY | 0.001408 | 10 | Signal transduction |
| REACTOME | Immune System | 0.001434 | 373 | Immune system |
| REACTOME | Retrograde transport at the Trans-Golgi-Network | 0.001455 | 18 | Transport and catabolism |
| REACTOME | DAG and IP3 signaling | 0.001476 | 16 | Signal transduction |
| REACTOME | Class I MHC mediated antigen processing & presentation | 0.001481 | 84 | Immune system |
| REACTOME | Signaling by Interleukins | 0.001488 | 98 | Immune system |
| KEGG | Dopaminergic synapse | 0.001523 | 36 | Nervous system |
| REACTOME | Mitotic Telophase/Cytokinesis | 0.001537 | 8 | Cell growth and death |
| WikiPathways | DNA damage response (only ATM dependent) | 0.001543 | 32 | Cell growth and death |
| KEGG | Longevity regulating pathway - multiple species | 0.001564 | 21 | Aging |
| REACTOME | Signaling by PDGF | 0.001679 | 20 | Signal transduction |
| BioCarta | BIOCARTA GPCR PATHWAY | 0.001712 | 13 | Signal transduction |
| KEGG | Long-term potentiation | 0.001821 | 22 | Nervous system |
| Panther_Pathway | T cell activation | 0.001837 | 24 | Immune system |
| REACTOME | Cell death signalling via NRAGE, NRIF and NADE | 0.001837 | 24 | Signal transduction |
| REACTOME | RAF-independent MAPK1/3 activation | 0.001846 | 11 | Signal transduction |
| REACTOME | RHOBTB2 GTPase cycle | 0.001846 | 11 | Signal transduction |
| SIGNOR 3.0 | Adipogenesis | 0.001846 | 11 | Development and regeneration |
| WikiPathways | Nuclear receptors | 0.001885 | 15 | Endocrine system |
| Panther_Pathway | p53 pathway feedback loops 2 | 0.001887 | 17 | Cell growth and death |
| WikiPathways | Translation inhibitors in chronically activated PDGFRA cells | 0.001887 | 17 | Signal transduction |
| ACSN2 | IMMUNOSUPPPRESSIVE CORE PATHWAYS | 0.001896 | 16 | Immune system |
| REACTOME | EPHB-mediated forward signaling | 0.001896 | 16 | Development and regeneration |
| REACTOME | RND3 GTPase cycle | 0.001896 | 16 | Signal transduction |
| WikiPathways | Splicing factor NOVA regulated synaptic proteins | 0.001896 | 16 | Nervous system |
| REACTOME | S Phase | 0.001957 | 42 | Cell growth and death |
| REACTOME | Translocation of SLC2A4 (GLUT4) to the plasma membrane | 0.001969 | 23 | Transport and catabolism |
| WikiPathways | Kit receptor signaling pathway | 0.002038 | 20 | Immune system |
| REACTOME | Inactivation of CDC42 and RAC1 | 0.002123 | 6 | Development and regeneration |
| REACTOME | Role of ABL in ROBO-SLIT signaling | 0.002123 | 6 | Development and regeneration |
| REACTOME | Ion transport by P-type ATPases | 0.002171 | 19 | Membrane transport |
| REACTOME | PTEN Regulation | 0.002217 | 37 | Signal transduction |
| KEGG | mTOR signaling pathway | 0.002223 | 40 | Signal transduction |
| KEGG | Gap junction | 0.002254 | 27 | Cellular community |
| WikiPathways | Synaptic vesicle pathway | 0.00228 | 18 | Nervous system |
| WikiPathways | Wnt signaling pathway | 0.00228 | 18 | Signal transduction |
| KEGG | GnRH signaling pathway | 0.002374 | 28 | Endocrine system |
| REACTOME | RND2 GTPase cycle | 0.002451 | 16 | Signal transduction |
| WikiPathways | TGF-beta signaling in thyroid cells for epithelial-mesenchymal transition | 0.002472 | 9 | Signal transduction |
| Panther_Pathway | Ras Pathway | 0.002628 | 22 | Signal transduction |
| REACTOME | Sema4D in semaphorin signaling | 0.002651 | 11 | Development and regeneration |
| REACTOME | Activated NTRK2 signals through FRS2 and FRS3 | 0.002682 | 7 | Signal transduction |
| REACTOME | Establishment of Sister Chromatid Cohesion | 0.002682 | 7 | Cell growth and death |
| REACTOME | AKT phosphorylates targets in the cytosol | 0.002703 | 8 | Signal transduction |
| WikiPathways | Hippo-Merlin signaling dysregulation | 0.002762 | 33 | Signal transduction |
| WikiPathways | p53 transcriptional gene network | 0.002776 | 28 | Cell growth and death |
| REACTOME | Signaling by BMP | 0.002998 | 12 | Signal transduction |
| BioCarta | BIOCARTA MTOR PATHWAY | 0.003276 | 10 | Signal transduction |
| Panther_Pathway | p53 pathway by glucose deprivation | 0.003276 | 10 | Cell growth and death |

| | | | | |
|---|---|---|---|---|
| SIGNOR 3.0 | TGF-beta Signaling | 0.003276 | 10 | Signal transduction |
| WikiPathways | p38 MAPK signaling pathway | 0.003282 | 13 | Signal transduction |
| ACSN2 | DEPENDANCE RECEPTORS | 0.003289 | 15 | Cell growth and death |
| WikiPathways | PDGF pathway | 0.003289 | 15 | Signal transduction |
| KEGG | p53 signaling pathway | 0.00346 | 23 | Cell growth and death |
| WikiPathways | Thyroxine (thyroid hormone) production | 0.003873 | 11 | Metabolism of amino acids and derivatives |
| REACTOME | PLC beta mediated events | 0.003894 | 17 | Signal transduction |
| WikiPathways | Interleukin-1 (IL-1) structural pathway | 0.003894 | 17 | Immune system |
| WikiPathways | Ciliary landscape | 0.00392 | 51 | Transport and catabolism |
| REACTOME | Regulation of cortical dendrite branching | 0.003938 | 4 | Development and regeneration |
| WikiPathways | Inhibition of exosome biogenesis and secretion by Manumycin A in CRPC cells | 0.003944 | 9 | Signal transduction |
| KEGG | Melanogenesis | 0.003945 | 29 | Endocrine system |
| KEGG | T cell receptor signaling pathway | 0.003945 | 29 | Immune system |
| WikiPathways | Wnt signaling | 0.004067 | 31 | Signal transduction |
| REACTOME | Mitotic G1 phase and G1/S transition | 0.004095 | 38 | Cell growth and death |
| WikiPathways | T-cell receptor signaling pathway | 0.004107 | 26 | Immune system |
| Panther_Pathway | Insulin/IGF pathway-mitogen activated protein kinase kinase/MAP kinase cascade | 0.004117 | 12 | Endocrine system |
| REACTOME | Myogenesis | 0.004117 | 12 | Development and regeneration |
| BioCarta | BIOCARTA P38MAPK PATHWAY | 0.004262 | 13 | Signal transduction |
| KEGG | Aldosterone-regulated sodium reabsorption | 0.004262 | 14 | Excretory system |
| REACTOME | Ca-dependent events | 0.004262 | 14 | Signal transduction |
| BioCarta | BIOCARTA BCR PATHWAY | 0.004264 | 13 | Immune system |
| REACTOME | Cargo recognition for clathrin-mediated endocytosis | 0.004429 | 29 | Transport and catabolism |
| REACTOME | G-protein mediated events | 0.00443 | 18 | Signal transduction |
| BioCarta | BIOCARTA CD40 PATHWAY | 0.004488 | 8 | Immune system |
| REACTOME | NCAM signaling for neurite out-growth | 0.00453 | 20 | Development and regeneration |
| BioCarta | BIOCARTA FBW7 PATHWAY | 0.00466 | 6 | Cell growth and death |
| PathBank | Rac 1 Cell Motility Signaling Pathway | 0.004713 | 10 | Signal transduction |
| WikiPathways | TROP2 regulatory signaling | 0.004717 | 17 | Signal transduction |
| WikiPathways | RalA downstream regulated genes | 0.004867 | 7 | Signal transduction |
| KEGG | Fc gamma R-mediated phagocytosis | 0.004868 | 27 | Immune system |
| REACTOME | TGF-beta receptor signaling activates SMADs | 0.005019 | 16 | Signal transduction |
| REACTOME | Deactivation of the beta-catenin transactivating complex | 0.005286 | 15 | Signal transduction |
| WikiPathways | EPO receptor signaling | 0.005287 | 11 | Signal transduction |
| WikiPathways | Estrogen signaling pathway | 0.005287 | 11 | Endocrine system |
| REACTOME | Signaling by FGFR2 | 0.005332 | 22 | Signal transduction |
| REACTOME | MET activates PTK2 signaling | 0.005534 | 12 | Signal transduction |
| SIGNOR 3.0 | P38 Signaling and Myogenesis | 0.005534 | 12 | Development and regeneration |
| WikiPathways | Extracellular vesicle-mediated signaling in recipient cells | 0.005534 | 12 | Immune system |
| BioCarta | BIOCARTA FMLP PATHWAY | 0.005549 | 13 | Immune system |
| PathBank | BCR Signaling Pathway | 0.005549 | 13 | Immune system |
| REACTOME | Oncogene Induced Senescence | 0.005549 | 13 | Cell growth and death |
| REACTOME | RHO GTPases Activate ROCKs | 0.005855 | 9 | Signal transduction |
| REACTOME | RHO GTPases activate CIT | 0.005855 | 9 | Signal transduction |
| KEGG | Platelet activation | 0.005917 | 33 | Immune system |
| KEGG | Tight junction | 0.005968 | 41 | Cellular community |
| WikiPathways | Hair follicle development: cytodifferentiation - part 3 of 3 | 0.006134 | 25 | Development and regeneration |
| KEGG | Endocrine and other factor-regulated calcium reabsorption | 0.006611 | 18 | Excretory system |
| REACTOME | Golgi Associated Vesicle Biogenesis | 0.006611 | 18 | Transport and catabolism |
| REACTOME | Signaling by SCF-KIT | 0.006658 | 15 | Signal transduction |
| KEGG | Chemokine signaling pathway | 0.00666 | 45 | Immune system |
| BioCarta | BIOCARTA FCER1 PATHWAY | 0.007049 | 14 | Immune system |
| REACTOME | Apoptotic execution phase | 0.007224 | 17 | Cell growth and death |
| REACTOME | G0 and Early G1 | 0.007231 | 11 | Cell growth and death |
| WikiPathways | Follicle stimulating hormone (FSH) signaling pathway | 0.007231 | 11 | Endocrine system |
| REACTOME | CaM pathway | 0.007256 | 13 | Signal transduction |
| REACTOME | Calmodulin induced events | 0.007256 | 13 | Signal transduction |
| REACTOME | Interaction between L1 and Ankyrins | 0.007275 | 12 | Development and regeneration |
| REACTOME | Activation of NMDA receptors and postsynaptic events | 0.007297 | 26 | Nervous system |
| WikiPathways | Corticotropin-releasing hormone signaling pathway | 0.007297 | 26 | Endocrine system |
| KEGG | C-type lectin receptor signaling pathway | 0.007508 | 28 | Immune system |
| REACTOME | Integration of energy metabolism | 0.007558 | 29 | Energy metabolism |
| ACSN2 | FATTY ACID BIOSYNTHESIS | 0.007726 | 16 | Lipid metabolism |
| REACTOME | NOTCH1 Intracellular Domain Regulates Transcription | 0.007726 | 16 | Signal transduction |
| BioCarta | BIOCARTA MELANOCYTE PATHWAY | 0.007991 | 5 | Development and regeneration |

| | | | | |
|---|---|---|---|---|
| BioCarta | BIOCARTA PCAF PATHWAY | 0.008209 | 7 | Immune system |
| BioCarta | BIOCARTA RB PATHWAY | 0.008209 | 7 | Cell growth and death |
| REACTOME | Chk1/Chk2(Cds1) mediated inactivation of Cyclin B:Cdk1 complex | 0.008209 | 7 | Cell growth and death |
| REACTOME | Opioid Signalling | 0.008225 | 25 | Signal transduction |
| WikiPathways | Genes controlling nephrogenesis | 0.008238 | 15 | Development and regeneration |
| REACTOME | FRS-mediated FGFR3 signaling | 0.008474 | 9 | Signal transduction |
| REACTOME | NOTCH4 Intracellular Domain Regulates Transcription | 0.008474 | 9 | Signal transduction |
| REACTOME | PKA-mediated phosphorylation of CREB | 0.008474 | 9 | Signal transduction |
| SIGNOR 3.0 | Axon guidance | 0.008474 | 9 | Development and regeneration |
| WikiPathways | Serotonin receptor 2 and ELK-SRF/GATA4 signaling | 0.008474 | 9 | Nervous system |
| REACTOME | RHO GTPases activate PKNs | 0.008489 | 19 | Signal transduction |
| BioCarta | BIOCARTA SKP2E2F PATHWAY | 0.008688 | 6 | Cell growth and death |
| REACTOME | MAP2K and MAPK activation | 0.008706 | 14 | Signal transduction |
| REACTOME | Signaling by FGFR3 | 0.008706 | 14 | Signal transduction |
| REACTOME | Signaling by NOTCH | 0.00888 | 47 | Signal transduction |
| BioCarta | BIOCARTA CELLCYCLE PATHWAY | 0.009181 | 10 | Cell growth and death |
| BioCarta | BIOCARTA VDR PATHWAY | 0.009181 | 10 | Endocrine system |
| PathBank | Ras Signaling Pathway | 0.009181 | 10 | Signal transduction |
| BioCarta | BIOCARTA PDGF PATHWAY | 0.009386 | 11 | Signal transduction |
| WikiPathways | Angiotensin II receptor type 1 pathway | 0.009386 | 11 | Endocrine system |
| WikiPathways | T cell receptor and co-stimulatory signaling | 0.009386 | 11 | Immune system |
| REACTOME | MyD88 cascade initiated on plasma membrane | 0.009459 | 26 | Immune system |
| REACTOME | Toll Like Receptor 10 (TLR10) Cascade | 0.009459 | 26 | Immune system |
| REACTOME | Toll Like Receptor 5 (TLR5) Cascade | 0.009459 | 26 | Immune system |

**Supplementary Table 4. Pathway enrichment analysis of cluster 2 endotype putative microRNA gene targets.** Gene targets of the microRNAs specific for each cluster were obtained from mirDIP and used to perform pathway enrichment analysis using pathDIP.

| Pathway Source | Pathway Name | q-value (BH method) | Genes mapped | Category |
|---|---|---|---|---|
| **Cluster 2** | | | | |
| REACTOME | Signal Transduction | 3.41E-32 | 452 | Signal transduction |
| REACTOME | Signaling by Receptor Tyrosine Kinases | 1.92E-21 | 135 | Signal transduction |
| ACSN2 | EMT REGULATORS | 5.42E-17 | 138 | Signal transduction |
| ACSN2 | PI3K AKT MTOR | 1.84E-15 | 85 | Signal transduction |
| WikiPathways | EGF/EGFR signaling pathway | 9.13E-15 | 57 | Signal transduction |
| REACTOME | Nervous system development | 5.14E-14 | 127 | Development and regeneration |
| REACTOME | Axon guidance | 3.40E-13 | 121 | Development and regeneration |
| KEGG | Axon guidance | 5.26E-13 | 58 | Development and regeneration |
| REACTOME | Developmental Biology | 1.07E-12 | 201 | Development and regeneration |
| ACSN2 | WNT NON CANONICAL | 3.86E-12 | 97 | Signal transduction |
| WikiPathways | EGFR tyrosine kinase inhibitor resistance | 1.92E-11 | 35 | Signal transduction |
| WikiPathways | VEGFA-VEGFR2 signaling | 1.59E-10 | 95 | Signal transduction |
| KEGG | Focal adhesion | 5.19E-10 | 56 | Cellular community |
| WikiPathways | Brain-derived neurotrophic factor (BDNF) signaling pathway | 8.53E-10 | 45 | Nervous system |
| Panther_Pathway | Integrin signalling pathway | 8.66E-10 | 49 | Signal transduction |
| REACTOME | Membrane Trafficking | 9.92E-10 | 122 | Transport and catabolism |
| ACSN2 | DEATH RECEPTOR PATHWAYS | 1.71E-09 | 64 | Signal transduction |
| REACTOME | Signaling by Rho GTPases | 1.71E-09 | 127 | Signal transduction |
| REACTOME | Signaling by Rho GTPases, Miro GTPases and RHOBTB3 | 1.74E-09 | 129 | Signal transduction |
| KEGG | PI3K-Akt signaling pathway | 2.38E-09 | 79 | Signal transduction |
| KEGG | Insulin signaling pathway | 4.50E-09 | 42 | Endocrine system |
| ACSN2 | HEDGEHOG | 4.82E-09 | 68 | Signal transduction |
| WikiPathways | Energy metabolism | 4.95E-09 | 23 | Energy metabolism |
| ACSN2 | APOPTOSIS | 5.20E-09 | 58 | Cell growth and death |
| WikiPathways | TGF-beta signaling pathway | 5.28E-09 | 41 | Signal transduction |
| KEGG | MAPK signaling pathway | 5.72E-09 | 69 | Signal transduction |
| WikiPathways | Insulin signaling | 6.54E-09 | 46 | Endocrine system |
| ACSN2 | RCD GENES | 8.00E-09 | 41 | Cell growth and death |
| Panther_Pathway | EGF receptor signaling pathway | 9.47E-09 | 37 | Signal transduction |
| KEGG | Long-term potentiation | 1.13E-08 | 27 | Nervous system |
| ACSN2 | STARVATION AUTOPHAGY | 1.38E-08 | 46 | Transport and catabolism |
| WikiPathways | Ectoderm differentiation | 1.42E-08 | 42 | Development and regeneration |
| REACTOME | Signaling by TGFB family members | 1.83E-08 | 38 | Signal transduction |
| REACTOME | Assembly of collagen fibrils and other multimeric structures | 3.02E-08 | 25 | Cellular community |
| WikiPathways | Focal adhesion | 3.69E-08 | 51 | Cellular community |
| REACTOME | Signaling by Nuclear Receptors | 3.72E-08 | 62 | Signal transduction |
| KEGG | Autophagy - animal | 4.77E-08 | 40 | Transport and catabolism |
| WikiPathways | Endoderm differentiation | 5.27E-08 | 41 | Development and regeneration |
| ACSN2 | MAPK | 5.65E-08 | 52 | Signal transduction |
| REACTOME | ESR-mediated signaling | 5.67E-08 | 49 | Signal transduction |
| ACSN2 | MOMP REGULATION | 5.69E-08 | 45 | Cell growth and death |
| REACTOME | Intracellular signaling by second messengers | 5.82E-08 | 68 | Signal transduction |
| REACTOME | VEGFA-VEGFR2 Pathway | 6.97E-08 | 32 | Signal transduction |
| REACTOME | Intrinsic Pathway for Apoptosis | 7.23E-08 | 23 | Cell growth and death |
| REACTOME | RHO GTPase cycle | 7.49E-08 | 89 | Signal transduction |
| WikiPathways | Mesodermal commitment pathway | 1.18E-07 | 42 | Development and regeneration |
| REACTOME | Collagen chain trimerization | 1.35E-07 | 20 | Cellular community |
| KEGG | Thyroid hormone signaling pathway | 1.47E-07 | 36 | Endocrine system |
| WikiPathways | Leptin signaling pathway | 1.81E-07 | 27 | Signal transduction |
| REACTOME | Signaling by VEGF | 1.82E-07 | 33 | Signal transduction |
| KEGG | Longevity regulating pathway - multiple species | 1.83E-07 | 24 | Aging |
| REACTOME | Vesicle-mediated transport | 1.85E-07 | 130 | Transport and catabolism |
| REACTOME | Signaling by PDGF | 2.09E-07 | 23 | Signal transduction |
| WikiPathways | PI3K-Akt signaling pathway | 2.14E-07 | 71 | Signal transduction |
| REACTOME | Signaling by NTRKs | 2.14E-07 | 38 | Signal transduction |
| KEGG | FoxO signaling pathway | 2.33E-07 | 37 | Signal transduction |
| WikiPathways | ErbB signaling pathway | 2.34E-07 | 30 | Signal transduction |
| REACTOME | Non-integrin membrane-ECM interactions | 2.83E-07 | 23 | Cellular community |
| KEGG | Dopaminergic synapse | 3.03E-07 | 37 | Nervous system |
| KEGG | Cellular senescence | 3.08E-07 | 41 | Cell growth and death |
| KEGG | Rap1 signaling pathway | 3.17E-07 | 51 | Signal transduction |

| | | | | |
|---|---|---|---|---|
| REACTOME | Collagen degradation | 3.17E-07 | 24 | Cellular community |
| WikiPathways | MAPK signaling pathway | 4.82E-07 | 56 | Signal transduction |
| REACTOME | Crosslinking of collagen fibrils | 5.16E-07 | 12 | Cellular community |
| REACTOME | PIP3 activates AKT signaling | 5.16E-07 | 59 | Signal transduction |
| KEGG | ErbB signaling pathway | 5.17E-07 | 28 | Signal transduction |
| WikiPathways | Focal adhesion: PI3K-Akt-mTOR-signaling pathway | 8.00E-07 | 64 | Cellular community |
| REACTOME | Collagen biosynthesis and modifying enzymes | 8.13E-07 | 24 | Cellular community |
| REACTOME | NCAM signaling for neurite out-growth | 1.03E-06 | 23 | Development and regeneration |
| REACTOME | Signaling by NTRK1 (TRKA) | 1.21E-06 | 33 | Signal transduction |
| REACTOME | L1CAM interactions | 1.32E-06 | 34 | Development and regeneration |
| REACTOME | Semaphorin interactions | 1.39E-06 | 23 | Development and regeneration |
| KEGG | Longevity regulating pathway | 1.41E-06 | 28 | Aging |
| REACTOME | Activation of BH3-only proteins | 1.68E-06 | 15 | Cell growth and death |
| REACTOME | Collagen formation | 1.79E-06 | 28 | Cellular community |
| Panther_Pathway | CCKR signaling map | 2.42E-06 | 42 | Signal transduction |
| ACSN2 | IMMUNOSTIMULATORY CORE PATHWAYS | 2.46E-06 | 30 | Immune system |
| KEGG | Adherens junction | 2.56E-06 | 24 | Cellular community |
| KEGG | Apelin signaling pathway | 3.16E-06 | 36 | Signal transduction |
| REACTOME | Signaling by TGF-beta Receptor Complex | 3.66E-06 | 28 | Signal transduction |
| ACSN2 | WNT CANONICAL | 4.56E-06 | 79 | Signal transduction |
| KEGG | Growth hormone synthesis, secretion and action | 4.86E-06 | 33 | Endocrine system |
| REACTOME | Extracellular matrix organization | 5.71E-06 | 61 | Cellular community |
| REACTOME | Anchoring fibril formation | 6.73E-06 | 10 | Cellular community |
| REACTOME | RAC1 GTPase cycle | 6.88E-06 | 43 | Signal transduction |
| Panther_Pathway | Insulin/IGF pathway-mitogen activated protein kinase kinase/MAP kinase cascade | 7.06E-06 | 14 | Endocrine system |
| REACTOME | Extra-nuclear estrogen signaling | 7.41E-06 | 24 | Signal transduction |
| KEGG | AMPK signaling pathway | 8.00E-06 | 32 | Signal transduction |
| Panther_Pathway | PDGF signaling pathway | 8.31E-06 | 33 | Signal transduction |
| REACTOME | NCAM1 interactions | 8.70E-06 | 17 | Development and regeneration |
| ACSN2 | TCR SIGNALING | 9.35E-06 | 45 | Immune system |
| KEGG | Tight junction | 9.76E-06 | 40 | Cellular community |
| ACSN2 | ECM | 1.16E-05 | 33 | Cellular community |
| BioCarta | BIOCARTA IGF1MTOR PATHWAY | 1.16E-05 | 11 | Endocrine system |
| Panther_Pathway | FGF signaling pathway | 1.17E-05 | 29 | Signal transduction |
| WikiPathways | Neurogenesis regulation in the olfactory epithelium | 1.22E-05 | 20 | Sensory system |
| KEGG | Oocyte meiosis | 1.38E-05 | 33 | Cell growth and death |
| REACTOME | Degradation of the extracellular matrix | 1.40E-05 | 35 | Cellular community |
| KEGG | TGF-beta signaling pathway | 1.41E-05 | 27 | Signal transduction |
| WikiPathways | CKAP4 signaling pathway map | 1.53E-05 | 31 | Signal transduction |
| ACSN2 | CASPASES | 1.59E-05 | 37 | Cell growth and death |
| REACTOME | VEGFR2 mediated vascular permeability | 1.64E-05 | 13 | Signal transduction |
| REACTOME | Signaling by MET | 1.81E-05 | 24 | Signal transduction |
| REACTOME | Integrin cell surface interactions | 2.08E-05 | 25 | Cellular community |
| REACTOME | Negative regulation of the PI3K/AKT network | 2.09E-05 | 30 | Signal transduction |
| KEGG | Neurotrophin signaling pathway | 2.13E-05 | 31 | Nervous system |
| KEGG | Relaxin signaling pathway | 2.60E-05 | 33 | Endocrine system |
| WikiPathways | Adipogenesis | 2.60E-05 | 33 | Development and regeneration |
| KEGG | Glucagon signaling pathway | 2.84E-05 | 29 | Endocrine system |
| WikiPathways | Translation inhibitors in chronically activated PDGFRA cells | 3.21E-05 | 17 | Signal transduction |
| WikiPathways | Thermogenesis | 4.16E-05 | 29 | Thermogenesis |
| REACTOME | Translocation of SLC2A4 (GLUT4) to the plasma membrane | 4.20E-05 | 22 | Transport and catabolism |
| KEGG | mTOR signaling pathway | 4.62E-05 | 36 | Signal transduction |
| REACTOME | Programmed Cell Death | 4.67E-05 | 45 | Cell growth and death |
| KEGG | cAMP signaling pathway | 4.87E-05 | 46 | Signal transduction |
| REACTOME | MAPK family signaling cascades | 5.43E-05 | 61 | Signal transduction |
| KEGG | Signaling pathways regulating pluripotency of stem cells | 5.63E-05 | 34 | Cellular community |
| Panther_Pathway | Angiogenesis | 6.04E-05 | 36 | Development and regeneration |
| ACSN2 | SENESCENCE | 7.17E-05 | 25 | Cell growth and death |
| REACTOME | Signaling by WNT | 7.27E-05 | 57 | Signal transduction |
| REACTOME | RHO GTPase Effectors | 8.03E-05 | 56 | Signal transduction |
| BioCarta | BIOCARTA MAPK PATHWAY | 9.16E-05 | 23 | Signal transduction |
| ACSN2 | DEPENDANCE RECEPTORS | 9.40E-05 | 15 | Cell growth and death |
| Panther_Pathway | Cadherin signaling pathway | 9.49E-05 | 35 | Cellular community |
| WikiPathways | Gastrin signaling pathway | 9.53E-05 | 29 | Endocrine system |
| REACTOME | Syndecan interactions | 9.90E-05 | 12 | Cellular community |
| Panther_Pathway | T cell activation | 9.93E-05 | 22 | Immune system |
| REACTOME | Apoptosis | 0.000111 | 39 | Cell growth and death |

| | | | | |
|---|---|---|---|---|
| WikiPathways | Factors and pathways affecting insulin-like growth factor (IGF1)-Akt signaling | 0.000111 | 14 | Endocrine system |
| KEGG | cGMP-PKG signaling pathway | 0.000112 | 37 | Signal transduction |
| KEGG | Hippo signaling pathway | 0.000143 | 35 | Signal transduction |
| WikiPathways | DYRK1A | 0.000163 | 19 | Development and regeneration |
| REACTOME | Deactivation of the beta-catenin transactivating complex | 0.000172 | 15 | Signal transduction |
| KEGG | Sphingolipid signaling pathway | 0.000178 | 29 | Signal transduction |
| KEGG | Ras signaling pathway | 0.000203 | 46 | Signal transduction |
| Panther_Pathway | Ras Pathway | 0.000233 | 20 | Signal transduction |
| BioCarta | BIOCARTA MTOR PATHWAY | 0.00026 | 10 | Signal transduction |
| REACTOME | RHO GTPases activate PAKs | 0.00026 | 10 | Signal transduction |
| REACTOME | Neuronal System | 0.000264 | 70 | Nervous system |
| REACTOME | Post NMDA receptor activation events | 0.000271 | 22 | Nervous system |
| REACTOME | PPARA activates gene expression | 0.000288 | 28 | Lipid metabolism |
| Panther_Pathway | Wnt signaling pathway | 0.000292 | 54 | Signal transduction |
| REACTOME | PI5P, PP2A and IER3 Regulate PI3K/AKT Signaling | 0.000349 | 26 | Signal transduction |
| REACTOME | Activation of NMDA receptors and postsynaptic events | 0.00035 | 24 | Nervous system |
| WikiPathways | Ras signaling | 0.000355 | 38 | Signal transduction |
| WikiPathways | CCL18 signaling pathway | 0.00037 | 13 | Immune system |
| KEGG | Oxytocin signaling pathway | 0.000371 | 34 | Endocrine system |
| REACTOME | Regulation of lipid metabolism by PPARalpha | 0.000384 | 28 | Lipid metabolism |
| WikiPathways | SREBF and miR33 in cholesterol and lipid homeostasis | 0.000386 | 9 | Lipid metabolism |
| REACTOME | Signaling by EGFR | 0.000386 | 16 | Signal transduction |
| REACTOME | Signaling by ERBB2 | 0.000386 | 16 | Signal transduction |
| BioCarta | BIOCARTA PPARG PATHWAY | 0.000392 | 6 | Endocrine system |
| REACTOME | Clathrin-mediated endocytosis | 0.000411 | 32 | Transport and catabolism |
| REACTOME | Nuclear Events (kinase and transcription factor activation) | 0.000417 | 18 | Signal transduction |
| Panther_Pathway | Apoptosis signaling pathway | 0.00046 | 26 | Cell growth and death |
| ACSN2 | CORE | 0.000464 | 19 | Signal transduction |
| WikiPathways | Target of rapamycin signaling | 0.000489 | 13 | Signal transduction |
| KEGG | Estrogen signaling pathway | 0.000492 | 31 | Endocrine system |
| REACTOME | MET promotes cell motility | 0.000505 | 14 | Signal transduction |
| WikiPathways | Canonical and non-canonical Notch signaling | 0.000519 | 11 | Signal transduction |
| KEGG | HIF-1 signaling pathway | 0.000526 | 26 | Signal transduction |
| ACSN2 | CELL CELL ADHESIONS | 0.000535 | 25 | Cellular community |
| REACTOME | Protein-protein interactions at synapses | 0.000535 | 22 | Nervous system |
| REACTOME | Activation of BAD and translocation to mitochondria | 0.000547 | 8 | Cell growth and death |
| KEGG | Adipocytokine signaling pathway | 0.000548 | 19 | Endocrine system |
| BioCarta | BIOCARTA HER2 PATHWAY | 0.00057 | 10 | Signal transduction |
| REACTOME | Gastrulation | 0.000589 | 16 | Development and regeneration |
| KEGG | Parathyroid hormone synthesis, secretion and action | 0.00059 | 26 | Endocrine system |
| REACTOME | Other semaphorin interactions | 0.000591 | 9 | Development and regeneration |
| KEGG | B cell receptor signaling pathway | 0.000596 | 21 | Immune system |
| Panther_Pathway | PI3 kinase pathway | 0.00061 | 15 | Signal transduction |
| REACTOME | EPH-Ephrin signaling | 0.000612 | 23 | Development and regeneration |
| REACTOME | Cell Cycle, Mitotic | 0.00064 | 84 | Cell growth and death |
| WikiPathways | Thyroid stimulating hormone (TSH) signaling pathway | 0.000643 | 19 | Endocrine system |
| KEGG | Phospholipase D signaling pathway | 0.000716 | 32 | Signal transduction |
| ACSN2 | CELL MATRIX ADHESIONS | 0.000731 | 18 | Cellular community |
| REACTOME | Estrogen-dependent gene expression | 0.000734 | 27 | Signal transduction |
| WikiPathways | RalA downstream regulated genes | 0.000736 | 7 | Signal transduction |
| WikiPathways | Somatroph axis (GH) and its relationship to dietary restriction and aging | 0.000748 | 5 | Aging |
| WikiPathways | DNA damage response (only ATM dependent) | 0.000761 | 26 | Cell growth and death |
| BioCarta | BIOCARTA MET PATHWAY | 0.000771 | 12 | Development and regeneration |
| REACTOME | CD28 co-stimulation | 0.000771 | 12 | Immune system |
| WikiPathways | BDNF-TrkB signaling | 0.000771 | 12 | Nervous system |
| REACTOME | Estrogen-dependent nuclear events downstream of ESR-membrane signaling | 0.000791 | 10 | Signal transduction |
| REACTOME | Transcriptional regulation of pluripotent stem cells | 0.000791 | 10 | Development and regeneration |
| WikiPathways | Kit receptor signaling pathway | 0.000792 | 17 | Immune system |
| REACTOME | ECM proteoglycans | 0.000793 | 20 | Cellular community |
| KEGG | Cholinergic synapse | 0.000856 | 26 | Nervous system |
| SIGNOR 3.0 | NOTCH Signaling | 0.000857 | 8 | Signal transduction |
| PathBank | Lysophosphatidic Acid LPA1 Signalling | 0.000857 | 6 | Signal transduction |
| REACTOME | Regulation of PTEN mRNA translation | 0.000857 | 6 | Signal transduction |
| WikiPathways | Interferon type I signaling pathways | 0.000859 | 16 | Immune system |
| WikiPathways | Sterol regulatory element-binding proteins (SREBP) signaling | 0.000891 | 19 | Lipid metabolism |

| | | | | |
|---|---|---|---|---|
| KEGG | Inflammatory mediator regulation of TRP channels | 0.000899 | 24 | Sensory system |
| REACTOME | Ca2+ pathway | 0.00095 | 17 | Signal transduction |
| REACTOME | Oncogene Induced Senescence | 0.001001 | 12 | Cell growth and death |
| WikiPathways | Hepatocyte growth factor receptor signaling | 0.001001 | 12 | Signal transduction |
| WikiPathways | AGE/RAGE pathway | 0.001015 | 18 | Immune system |
| WikiPathways | Integrin-mediated cell adhesion | 0.001036 | 24 | Cellular community |
| WikiPathways | Hippo-Merlin signaling dysregulation | 0.001038 | 27 | Signal transduction |
| REACTOME | RHOJ GTPase cycle | 0.001044 | 16 | Signal transduction |
| KEGG | Fc gamma R-mediated phagocytosis | 0.001054 | 23 | Immune system |
| KEGG | GnRH signaling pathway | 0.001054 | 23 | Endocrine system |
| WikiPathways | Apoptosis | 0.001066 | 22 | Cell growth and death |
| REACTOME | CDC42 GTPase cycle | 0.001189 | 32 | Signal transduction |
| KEGG | Gap junction | 0.001213 | 22 | Cellular community |
| REACTOME | Activation of RAC1 | 0.001223 | 7 | Development and regeneration |
| WikiPathways | Lactate shuttle in glial cells | 0.001223 | 7 | Nervous system |
| ACSN2 | LYSOSOME ENDOSOME | 0.001226 | 9 | Transport and catabolism |
| Panther_Pathway | p53 pathway by glucose deprivation | 0.001226 | 9 | Cell growth and death |
| SIGNOR 3.0 | MTOR Signaling | 0.001226 | 9 | Signal transduction |
| REACTOME | Signaling by NOTCH | 0.001237 | 39 | Signal transduction |
| Panther_Pathway | Insulin/IGF pathway-protein kinase B signaling cascade | 0.001242 | 12 | Endocrine system |
| WikiPathways | Leukocyte-intrinsic Hippo pathway functions | 0.001242 | 12 | Signal transduction |
| WikiPathways | Embryonic stem cell pluripotency pathways | 0.001245 | 26 | Cellular community |
| BioCarta | BIOCARTA GPCR PATHWAY | 0.001247 | 11 | Signal transduction |
| REACTOME | Laminin interactions | 0.001247 | 11 | Cellular community |
| SIGNOR 3.0 | P38 Signaling and Myogenesis | 0.001247 | 11 | Development and regeneration |
| WikiPathways | Hippocampal synaptogenesis and neurogenesis | 0.001247 | 10 | Signal transduction |
| WikiPathways | PI3K-AKT-mTOR signaling pathway and therapeutic opportunities | 0.001247 | 11 | Signal transduction |
| KEGG | Melanogenesis | 0.001273 | 24 | Endocrine system |
| BioCarta | BIOCARTA NDKDYNAMIN PATHWAY | 0.001281 | 8 | Nervous system |
| WikiPathways | Canonical and non-canonical TGF-B signaling | 0.001281 | 8 | Signal transduction |
| WikiPathways | TGF-beta signaling in thyroid cells for epithelial-mesenchymal transition | 0.001281 | 8 | Signal transduction |
| REACTOME | RHOC GTPase cycle | 0.001398 | 19 | Signal transduction |
| REACTOME | Signaling by NOTCH1 | 0.001398 | 19 | Signal transduction |
| REACTOME | Cargo recognition for clathrin-mediated endocytosis | 0.001471 | 24 | Transport and catabolism |
| WikiPathways | Endochondral ossification | 0.001575 | 17 | Development and regeneration |
| WikiPathways | Clock-controlled autophagy in bone metabolism | 0.001662 | 20 | Transport and catabolism |
| REACTOME | Antigen processing: Ubiquitination & Proteasome degradation | 0.001667 | 53 | Immune system |
| REACTOME | NR1H2 and NR1H3-mediated signaling | 0.001838 | 14 | Signal transduction |
| ACSN2 | NECROPTOSIS | 0.001845 | 25 | Cell growth and death |
| BioCarta | BIOCARTA MELANOCYTE PATHWAY | 0.001854 | 5 | Development and regeneration |
| BioCarta | BIOCARTA CALCINEURIN PATHWAY | 0.001984 | 8 | Development and regeneration |
| Panther_Pathway | Axon guidance mediated by Slit/Robo | 0.001984 | 8 | Development and regeneration |
| WikiPathways | miR-509-3p alteration of YAP1/ECM axis | 0.001984 | 8 | Cellular community |
| REACTOME | RHOD GTPase cycle | 0.002006 | 15 | Signal transduction |
| BioCarta | BIOCARTA PYK2 PATHWAY | 0.002009 | 10 | Signal transduction |
| WikiPathways | Hippo signaling regulation pathways | 0.002021 | 23 | Signal transduction |
| WikiPathways | Wnt signaling pathway and pluripotency | 0.002021 | 23 | Cellular community |
| REACTOME | NR1H3 & NR1H2 regulate gene expression linked to cholesterol transport and efflux | 0.002048 | 12 | Signal transduction |
| KEGG | Leukocyte transendothelial migration | 0.002049 | 25 | Immune system |
| ACSN2 | GLUCOSE METABOLISM | 0.002335 | 25 | Carbohydrate metabolism |
| REACTOME | Transmission across Chemical Synapses | 0.002475 | 47 | Nervous system |
| REACTOME | Integration of energy metabolism | 0.002477 | 24 | Energy metabolism |
| REACTOME | Signaling by SCF-KIT | 0.002477 | 13 | Signal transduction |
| WikiPathways | IL6 signaling pathway | 0.002477 | 13 | Immune system |
| REACTOME | Opioid Signalling | 0.002494 | 21 | Signal transduction |
| REACTOME | Transport of small molecules | 0.0026 | 105 | Membrane transport |
| PathBank | Intracellular Signalling Through Adenosine Receptor A2a and Adenosine | 0.002628 | 12 | Signal transduction |
| PathBank | Intracellular Signalling Through Adenosine Receptor A2b and Adenosine | 0.002628 | 12 | Signal transduction |
| WikiPathways | Photodynamic therapy-induced HIF-1 survival signaling | 0.002628 | 12 | Signal transduction |
| REACTOME | Signaling by BMP | 0.002653 | 10 | Signal transduction |
| BioCarta | BIOCARTA AGR PATHWAY | 0.002686 | 11 | Nervous system |
| WikiPathways | Alpha 6 beta 4 signaling pathway | 0.002686 | 11 | Cellular community |
| REACTOME | Cytokine Signaling in Immune system | 0.002842 | 104 | Immune system |
| Panther_Pathway | Axon guidance mediated by semaphorins | 0.002849 | 8 | Development and regeneration |

| | | | | |
|---|---|---|---|---|
| REACTOME | VEGFR2 mediated cell proliferation | 0.002849 | 8 | Signal transduction |
| WikiPathways | TGF-beta receptor signaling | 0.002865 | 15 | Signal transduction |
| KEGG | T cell receptor signaling pathway | 0.002876 | 23 | Immune system |
| BioCarta | BIOCARTA LEPTIN PATHWAY | 0.00288 | 6 | Signal transduction |
| BioCarta | BIOCARTA VITCB PATHWAY | 0.00288 | 6 | Nervous system |
| REACTOME | Signaling by Leptin | 0.00288 | 6 | Signal transduction |
| Panther_Pathway | Heterotrimeric G-protein signaling pathway-Gi alpha and Gs alpha mediated pathway | 0.002956 | 30 | Signal transduction |
| WikiPathways | Notch signaling pathway | 0.002956 | 16 | Signal transduction |
| Panther_Pathway | p53 pathway | 0.002972 | 19 | Cell growth and death |
| REACTOME | Regulation of insulin secretion | 0.002972 | 19 | Energy metabolism |
| WikiPathways | AMP-activated protein kinase signaling | 0.002991 | 17 | Signal transduction |
| BioCarta | BIOCARTA PGC1A PATHWAY | 0.003003 | 7 | Energy metabolism |
| PathBank | Leucine Stimulation on Insulin Signaling | 0.003003 | 7 | Endocrine system |
| REACTOME | Signaling by Activin | 0.003003 | 7 | Signal transduction |
| Panther_Pathway | TGF-beta signaling pathway | 0.003139 | 21 | Signal transduction |
| WikiPathways | TROP2 regulatory signaling | 0.003191 | 14 | Signal transduction |
| PathBank | Ras Signaling Pathway | 0.003213 | 9 | Signal transduction |
| SIGNOR 3.0 | WNT Signaling and Myogenesis | 0.003213 | 9 | Signal transduction |
| ACSN2 | G1 S CHECKPOINT | 0.003325 | 11 | Cell growth and death |
| REACTOME | RAF activation | 0.003325 | 11 | Signal transduction |
| REACTOME | Golgi Associated Vesicle Biogenesis | 0.003346 | 15 | Transport and catabolism |
| REACTOME | PTEN Regulation | 0.003458 | 28 | Signal transduction |
| REACTOME | Oxidative Stress Induced Senescence | 0.003563 | 21 | Cell growth and death |
| REACTOME | Cell Cycle | 0.003642 | 96 | Cell growth and death |
| BioCarta | BIOCARTA TERT PATHWAY | 0.003751 | 5 | Aging |
| REACTOME | Competing endogenous RNAs (ceRNAs) regulate PTEN translation | 0.003751 | 5 | Signal transduction |
| WikiPathways | Caloric restriction and aging | 0.003751 | 5 | Aging |
| HumanCyc | BMP Signalling Pathway | 0.003813 | 4 | Signal transduction |
| WikiPathways | Wnt signaling pathway | 0.00382 | 14 | Signal transduction |
| REACTOME | MAP kinase activation | 0.004024 | 16 | Immune system |
| REACTOME | Amplification of signal from the kinetochores | 0.004034 | 21 | Cell growth and death |
| REACTOME | Amplification of signal from unattached kinetochores via a MAD2 inhibitory signal | 0.004034 | 21 | Cell growth and death |
| KEGG | ECM-receptor interaction | 0.004238 | 20 | Cellular community |
| REACTOME | RAC2 GTPase cycle | 0.004238 | 20 | Signal transduction |
| WikiPathways | Hair follicle development: cytodifferentiation - part 3 of 3 | 0.004238 | 19 | Development and regeneration |
| BioCarta | BIOCARTA KERATINOCYTE PATHWAY | 0.004256 | 13 | Development and regeneration |
| REACTOME | TGF-beta receptor signaling activates SMADs | 0.004256 | 13 | Signal transduction |
| BioCarta | BIOCARTA BAD PATHWAY | 0.004262 | 9 | Cell growth and death |
| BioCarta | BIOCARTA CARM ER PATHWAY | 0.004262 | 9 | Endocrine system |
| REACTOME | MET activates PTK2 signaling | 0.004276 | 10 | Signal transduction |
| WikiPathways | Extracellular vesicle-mediated signaling in recipient cells | 0.004276 | 10 | Immune system |
| BioCarta | BIOCARTA PITX2 PATHWAY | 0.004364 | 7 | Development and regeneration |
| REACTOME | CRMPs in Sema3A signaling | 0.004364 | 7 | Development and regeneration |
| REACTOME | Sema3A PAK dependent Axon repulsion | 0.004364 | 7 | Development and regeneration |
| BioCarta | BIOCARTA PPARA PATHWAY | 0.004435 | 14 | Endocrine system |
| WikiPathways | G protein signaling pathways | 0.004443 | 21 | Signal transduction |
| KEGG | Cell adhesion molecules | 0.004477 | 27 | Cellular community |
| PathBank | Nitric Oxide Signaling Pathway | 0.004478 | 6 | Signal transduction |
| REACTOME | Frs2-mediated activation | 0.004478 | 6 | Signal transduction |
| SIGNOR 3.0 | PI3K/AKT Signaling | 0.004478 | 6 | Signal transduction |
| WikiPathways | Disruption of postsynaptic signaling by CNV | 0.005136 | 10 | Nervous system |
| KEGG | VEGF signaling pathway | 0.005465 | 15 | Signal transduction |
| REACTOME | EGFR downregulation | 0.005481 | 10 | Signal transduction |
| SIGNOR 3.0 | P38 Signaling | 0.005481 | 10 | Signal transduction |
| REACTOME | Cellular Senescence | 0.005521 | 31 | Cell growth and death |
| REACTOME | TCF dependent signaling in response to WNT | 0.00557 | 36 | Signal transduction |
| REACTOME | Class I MHC mediated antigen processing & presentation | 0.005607 | 60 | Immune system |
| WikiPathways | B cell receptor signaling pathway | 0.005691 | 21 | Immune system |
| KEGG | Glutamatergic synapse | 0.005713 | 24 | Nervous system |
| REACTOME | EML4 and NUDC in mitotic spindle formation | 0.005713 | 24 | Cell growth and death |
| REACTOME | RND1 GTPase cycle | 0.005727 | 12 | Signal transduction |
| REACTOME | MAPK1/MAPK3 signaling | 0.006021 | 47 | Signal transduction |
| KEGG | Apoptosis | 0.006033 | 27 | Cell growth and death |
| REACTOME | NOTCH1 Intracellular Domain Regulates Transcription | 0.006087 | 13 | Signal transduction |
| REACTOME | Mitotic Spindle Checkpoint | 0.006172 | 23 | Cell growth and death |
| WikiPathways | Leptin-insulin signaling overlap | 0.006274 | 7 | Endocrine system |

| | | | | |
|---|---|---|---|---|
| WikiPathways | MAP3K1 role in promoting and blocking gonadal determination | 0.006274 | 7 | Development and regeneration |
| WikiPathways | NOTCH1 regulation of endothelial cell calcification | 0.006274 | 7 | Signal transduction |
| BioCarta | BIOCARTA IL2RB PATHWAY | 0.006277 | 11 | Immune system |
| WikiPathways | Serotonin HTR1 group and FOS pathway | 0.006277 | 11 | Nervous system |
| REACTOME | RAF/MAP kinase cascade | 0.006547 | 46 | Signal transduction |
| PathBank | Excitatory Neural Signalling Through 5-HTR 4 and Serotonin | 0.006634 | 5 | Nervous system |
| PathBank | Excitatory Neural Signalling Through 5-HTR 6 and Serotonin | 0.006634 | 5 | Nervous system |
| PathBank | Excitatory Neural Signalling Through 5-HTR 7 and Serotonin | 0.006634 | 5 | Nervous system |
| PathBank | Intracellular Signalling Through Histamine H2 Receptor and Histamine | 0.006634 | 5 | Signal transduction |
| PathBank | Lysophosphatidic Acid LPA2 Signalling | 0.006634 | 5 | Signal transduction |
| ACSN2 | ADHERENS JUNCTIONS | 0.006737 | 10 | Cellular community |
| REACTOME | Resolution of Sister Chromatid Cohesion | 0.006853 | 25 | Cell growth and death |
| SIGNOR 3.0 | Glutamatergic synapse | 0.006853 | 6 | Nervous system |
| KEGG | Progesterone-mediated oocyte maturation | 0.006861 | 21 | Endocrine system |
| BioCarta | BIOCARTA CREB PATHWAY | 0.00688 | 8 | Signal transduction |
| BioCarta | BIOCARTA EDG1 PATHWAY | 0.00688 | 8 | Signal transduction |
| REACTOME | SHC1 events in ERBB2 signaling | 0.00688 | 8 | Signal transduction |
| SIGNOR 3.0 | Insulin Signaling | 0.00688 | 8 | Endocrine system |
| WikiPathways | Transcription factor regulation in adipogenesis | 0.00688 | 8 | Development and regeneration |
| REACTOME | G0 and Early G1 | 0.006897 | 9 | Cell growth and death |
| REACTOME | Signaling by ALK | 0.006897 | 9 | Signal transduction |
| REACTOME | RHO GTPases Activate Formins | 0.006955 | 27 | Signal transduction |
| KEGG | Hedgehog signaling pathway | 0.007076 | 14 | Signal transduction |
| REACTOME | Ion transport by P-type ATPases | 0.007076 | 14 | Membrane transport |
| REACTOME | Neurotransmitter receptors and postsynaptic signal transmission | 0.007196 | 36 | Nervous system |
| Panther_Pathway | Interleukin signaling pathway | 0.007288 | 18 | Immune system |
| WikiPathways | Thyroid hormones production and peripheral downstream signaling effects | 0.0073 | 20 | Metabolism of amino acids and derivatives |
| REACTOME | FLT3 Signaling | 0.007404 | 11 | Immune system |
| WikiPathways | Nuclear receptors | 0.007404 | 11 | Endocrine system |
| KEGG | Platelet activation | 0.007517 | 25 | Immune system |
| REACTOME | RHOG GTPase cycle | 0.007587 | 17 | Signal transduction |
| REACTOME | Ion channel transport | 0.007639 | 33 | Membrane transport |
| BioCarta | BIOCARTA TCR PATHWAY | 0.007877 | 12 | Immune system |
| KEGG | Mitophagy - animal | 0.007881 | 16 | Transport and catabolism |
| WikiPathways | Cell cycle | 0.008077 | 24 | Cell growth and death |
| Panther_Pathway | p38 MAPK pathway | 0.008099 | 10 | Signal transduction |
| WikiPathways | CAMKK2 pathway | 0.008099 | 10 | Signal transduction |
| WikiPathways | MAPK cascade | 0.008099 | 10 | Signal transduction |
| ACSN2 | TNF RESPONSE | 0.008102 | 13 | Signal transduction |
| REACTOME | Signaling by FGFR1 | 0.008102 | 13 | Signal transduction |
| PathBank | Dopamine Activation of Neurological Reward System | 0.008167 | 4 | Nervous system |
| BioCarta | BIOCARTA MEF2D PATHWAY | 0.008206 | 7 | Immune system |
| REACTOME | Gastrin-CREB signalling pathway via PKC and MAPK | 0.008206 | 7 | Signal transduction |
| WikiPathways | Inhibition of exosome biogenesis and secretion by Manumycin A in CRPC cells | 0.008206 | 7 | Signal transduction |
| BioCarta | BIOCARTA PDGF PATHWAY | 0.00852 | 9 | Signal transduction |
| REACTOME | CREB1 phosphorylation through NMDA receptor-mediated activation of RAS signaling | 0.00852 | 9 | Nervous system |
| BioCarta | BIOCARTA IGF1R PATHWAY | 0.008684 | 8 | Endocrine system |
| REACTOME | Formation of paraxial mesoderm | 0.008684 | 8 | Development and regeneration |
| BioCarta | BIOCARTA FCER1 PATHWAY | 0.008782 | 11 | Immune system |
| REACTOME | NGF-stimulated transcription | 0.008782 | 11 | Signal transduction |
| WikiPathways | Chemokine signaling pathway | 0.009399 | 30 | Immune system |
| BioCarta | BIOCARTA PLATELETAPP PATHWAY | 0.009727 | 6 | Immune system |
| REACTOME | Prolonged ERK activation events | 0.009727 | 6 | Signal transduction |
| REACTOME | SEMA3A-Plexin repulsion signaling by inhibiting Integrin adhesion | 0.009727 | 6 | Development and regeneration |
| SIGNOR 3.0 | NOTCH Signaling and Myogenesis | 0.009727 | 6 | Signal transduction |
| WikiPathways | H19 action Rb-E2F1 signaling and CDK-Beta-catenin activity | 0.009727 | 6 | Signal transduction |
| WikiPathways | Prolactin signaling pathway | 0.009741 | 17 | Endocrine system |
| REACTOME | Cell-Cell communication | 0.00975 | 25 | Cellular community |
| BioCarta | BIOCARTA FMLP PATHWAY | 0.009769 | 10 | Immune system |
| REACTOME | Signalling to ERKs | 0.009769 | 10 | Signal transduction |
| REACTOME | MyD88 cascade initiated on plasma membrane | 0.009956 | 20 | Immune system |
| REACTOME | Toll Like Receptor 10 (TLR10) Cascade | 0.009956 | 20 | Immune system |
| REACTOME | Toll Like Receptor 5 (TLR5) Cascade | 0.009956 | 20 | Immune system |

**Supplementary Table 5. Pathway enrichment analysis of cluster 3 endotype putative microRNA gene targets.** Gene targets of the microRNAs specific for each cluster were obtained from mirDIP and used to perform pathway enrichment analysis using pathDIP.

| Pathway Source | Pathway Name | q-value (BH method) | Genes mapped | Category |
|---|---|---|---|---|
| **Cluster 3** | | | | |
| REACTOME | Signal Transduction | 2.23E-38 | 483 | Signal transduction |
| ACSN2 | EMT REGULATORS | 6.83E-17 | 142 | Signal transduction |
| REACTOME | Membrane Trafficking | 4.92E-13 | 136 | Transport and catabolism |
| ACSN2 | WNT CANONICAL | 1.82E-12 | 102 | Signal transduction |
| REACTOME | Signaling by Rho GTPases, Miro GTPases and RHOBTB3 | 1.25E-11 | 141 | Signal transduction |
| ACSN2 | WNT NON CANONICAL | 1.50E-11 | 96 | Signal transduction |
| REACTOME | Signaling by Rho GTPases | 1.52E-11 | 138 | Signal transduction |
| REACTOME | Signaling by Receptor Tyrosine Kinases | 1.62E-11 | 115 | Signal transduction |
| WikiPathways | Mesodermal commitment pathway | 1.64E-11 | 51 | Development and regeneration |
| ACSN2 | PI3K AKT MTOR | 3.15E-11 | 78 | Signal transduction |
| REACTOME | RHO GTPase cycle | 4.57E-11 | 102 | Signal transduction |
| WikiPathways | Brain-derived neurotrophic factor (BDNF) signaling pathway | 1.14E-09 | 46 | Nervous system |
| KEGG | MAPK signaling pathway | 1.39E-09 | 73 | Signal transduction |
| REACTOME | Nervous system development | 2.47E-09 | 117 | Development and regeneration |
| REACTOME | Developmental Biology | 6.17E-09 | 193 | Development and regeneration |
| WikiPathways | Adipogenesis | 8.07E-09 | 42 | Development and regeneration |
| WikiPathways | TGF-beta signaling pathway | 8.07E-09 | 42 | Signal transduction |
| KEGG | Axon guidance | 1.28E-08 | 51 | Development and regeneration |
| Panther_Pathway | Angiogenesis | 1.33E-08 | 46 | Development and regeneration |
| WikiPathways | Energy metabolism | 1.33E-08 | 23 | Energy metabolism |
| WikiPathways | VEGFA-VEGFR2 signaling | 1.38E-08 | 92 | Signal transduction |
| REACTOME | Intracellular signaling by second messengers | 1.56E-08 | 72 | Signal transduction |
| REACTOME | Vesicle-mediated transport | 1.92E-08 | 138 | Transport and catabolism |
| KEGG | Wnt signaling pathway | 2.70E-08 | 47 | Signal transduction |
| REACTOME | MAPK family signaling cascades | 6.08E-08 | 73 | Signal transduction |
| KEGG | Cellular senescence | 6.57E-08 | 44 | Cell growth and death |
| REACTOME | Signaling by TGFB family members | 7.43E-08 | 38 | Signal transduction |
| WikiPathways | MAPK signaling pathway | 9.84E-08 | 60 | Signal transduction |
| WikiPathways | EGF/EGFR signaling pathway | 1.29E-07 | 45 | Signal transduction |
| KEGG | Oxytocin signaling pathway | 1.40E-07 | 41 | Endocrine system |
| ACSN2 | HEDGEHOG | 1.53E-07 | 66 | Signal transduction |
| WikiPathways | Endochondral ossification | 1.78E-07 | 25 | Development and regeneration |
| KEGG | Signaling pathways regulating pluripotency of stem cells | 2.15E-07 | 41 | Cellular community |
| REACTOME | Axon guidance | 2.32E-07 | 106 | Development and regeneration |
| REACTOME | Ca2+ pathway | 2.80E-07 | 24 | Signal transduction |
| ACSN2 | MAPK | 2.83E-07 | 52 | Signal transduction |
| KEGG | Dopaminergic synapse | 4.59E-07 | 35 | Nervous system |
| REACTOME | RHOD GTPase cycle | 4.99E-07 | 22 | Signal transduction |
| ACSN2 | RCD GENES | 9.45E-07 | 38 | Cell growth and death |
| WikiPathways | TGF-beta receptor signaling | 1.07E-06 | 22 | Signal transduction |
| REACTOME | PIP3 activates AKT signaling | 1.53E-06 | 60 | Signal transduction |
| WikiPathways | Endoderm differentiation | 1.86E-06 | 39 | Development and regeneration |
| WikiPathways | Hippo signaling regulation pathways | 2.38E-06 | 28 | Signal transduction |
| KEGG | PI3K-Akt signaling pathway | 3.36E-06 | 72 | Signal transduction |
| WikiPathways | Estrogen signaling pathway | 3.88E-06 | 11 | Endocrine system |
| REACTOME | Signaling by TGF-beta Receptor Complex | 4.06E-06 | 29 | Signal transduction |
| REACTOME | RHOQ GTPase cycle | 4.11E-06 | 22 | Signal transduction |
| WikiPathways | Ectoderm differentiation | 4.19E-06 | 38 | Development and regeneration |
| BioCarta | BIOCARTA GPCR PATHWAY | 4.39E-06 | 15 | Signal transduction |
| KEGG | Focal adhesion | 4.40E-06 | 48 | Cellular community |
| REACTOME | RHOJ GTPase cycle | 4.52E-06 | 21 | Signal transduction |
| KEGG | TGF-beta signaling pathway | 4.59E-06 | 29 | Signal transduction |
| WikiPathways | Focal adhesion: PI3K-Akt-mTOR-signaling pathway | 4.71E-06 | 64 | Cellular community |
| KEGG | Parathyroid hormone synthesis, secretion and action | 4.72E-06 | 29 | Endocrine system |
| WikiPathways | Embryonic stem cell pluripotency pathways | 5.22E-06 | 33 | Cellular community |
| WikiPathways | G protein signaling pathways | 5.52E-06 | 26 | Signal transduction |
| KEGG | Endocytosis | 6.48E-06 | 55 | Transport and catabolism |
| KEGG | FoxO signaling pathway | 7.54E-06 | 35 | Signal transduction |
| ACSN2 | IMMUNOSTIMULATORY CORE PATHWAYS | 7.89E-06 | 30 | Immune system |
| ACSN2 | STARVATION AUTOPHAGY | 7.90E-06 | 41 | Transport and catabolism |
| KEGG | cAMP signaling pathway | 8.39E-06 | 47 | Signal transduction |

| | | | | |
|---|---|---|---|---|
| KEGG | Renin secretion | 8.81E-06 | 21 | Endocrine system |
| Panther_Pathway | CCKR signaling map | 8.84E-06 | 42 | Signal transduction |
| REACTOME | Signaling by BMP | 8.95E-06 | 14 | Signal transduction |
| KEGG | Cell cycle | 8.99E-06 | 34 | Cell growth and death |
| WikiPathways | Wnt signaling | 9.02E-06 | 32 | Signal transduction |
| REACTOME | ESR-mediated signaling | 9.07E-06 | 45 | Signal transduction |
| WikiPathways | Insulin signaling | 9.66E-06 | 40 | Endocrine system |
| KEGG | Glucagon signaling pathway | 9.73E-06 | 28 | Endocrine system |
| REACTOME | CDC42 GTPase cycle | 1.10E-05 | 39 | Signal transduction |
| WikiPathways | Cell cycle | 1.17E-05 | 33 | Cell growth and death |
| WikiPathways | Focal adhesion | 1.24E-05 | 46 | Cellular community |
| REACTOME | RHOV GTPase cycle | 1.37E-05 | 16 | Signal transduction |
| REACTOME | Intra-Golgi and retrograde Golgi-to-ER traffic | 1.58E-05 | 46 | Transport and catabolism |
| REACTOME | Estrogen-dependent gene expression | 1.81E-05 | 32 | Signal transduction |
| WikiPathways | PI3K-Akt signaling pathway | 1.89E-05 | 67 | Signal transduction |
| REACTOME | Cellular Senescence | 1.94E-05 | 40 | Cell growth and death |
| WikiPathways | Hippocampal synaptogenesis and neurogenesis | 2.05E-05 | 13 | Signal transduction |
| WikiPathways | Hedgehog signaling pathway | 2.10E-05 | 18 | Signal transduction |
| Panther_Pathway | EGF receptor signaling pathway | 2.37E-05 | 31 | Signal transduction |
| KEGG | Long-term potentiation | 2.84E-05 | 22 | Nervous system |
| KEGG | Melanogenesis | 2.93E-05 | 26 | Endocrine system |
| BioCarta | BIOCARTA TCR PATHWAY | 3.49E-05 | 17 | Immune system |
| WikiPathways | Thermogenesis | 3.84E-05 | 27 | Thermogenesis |
| REACTOME | Signaling by NTRKs | 3.86E-05 | 34 | Signal transduction |
| Panther_Pathway | Wnt signaling pathway | 3.95E-05 | 59 | Signal transduction |
| ACSN2 | APOPTOSIS | 4.17E-05 | 49 | Cell growth and death |
| REACTOME | RHOG GTPase cycle | 4.37E-05 | 23 | Signal transduction |
| WikiPathways | Thyroid stimulating hormone (TSH) signaling pathway | 4.46E-05 | 19 | Endocrine system |
| REACTOME | EGR2 and SOX10-mediated initiation of Schwann cell myelination | 4.61E-05 | 13 | Development and regeneration |
| ACSN2 | MOMP REGULATION | 5.04E-05 | 39 | Cell growth and death |
| KEGG | Calcium signaling pathway | 5.09E-05 | 48 | Signal transduction |
| REACTOME | Signaling by NOTCH | 5.43E-05 | 45 | Signal transduction |
| Panther_Pathway | PDGF signaling pathway | 5.56E-05 | 32 | Signal transduction |
| REACTOME | RHO GTPases activate PAKs | 6.06E-05 | 11 | Signal transduction |
| SIGNOR 3.0 | TGF-beta Signaling | 6.06E-05 | 11 | Signal transduction |
| Panther_Pathway | TGF-beta signaling pathway | 6.44E-05 | 26 | Signal transduction |
| BioCarta | BIOCARTA BCR PATHWAY | 6.57E-05 | 14 | Immune system |
| BioCarta | BIOCARTA MET PATHWAY | 6.57E-05 | 14 | Development and regeneration |
| REACTOME | Golgi Associated Vesicle Biogenesis | 6.87E-05 | 19 | Transport and catabolism |
| REACTOME | Signaling by NTRK1 (TRKA) | 6.99E-05 | 30 | Signal transduction |
| KEGG | Thyroid hormone signaling pathway | 7.23E-05 | 31 | Endocrine system |
| ACSN2 | ECM | 7.39E-05 | 32 | Cellular community |
| BioCarta | BIOCARTA CALCINEURIN PATHWAY | 7.93E-05 | 10 | Development and regeneration |
| REACTOME | RAC1 GTPase cycle | 0.000102 | 41 | Signal transduction |
| BioCarta | BIOCARTA VIP PATHWAY | 0.000106 | 12 | Immune system |
| WikiPathways | EGFR tyrosine kinase inhibitor resistance | 0.000111 | 24 | Signal transduction |
| REACTOME | Immune System | 0.000112 | 289 | Immune system |
| BioCarta | BIOCARTA FCER1 PATHWAY | 0.000115 | 15 | Immune system |
| WikiPathways | Wnt signaling pathway and pluripotency | 0.000117 | 27 | Cellular community |
| REACTOME | Signaling by WNT | 0.000125 | 58 | Signal transduction |
| REACTOME | RHOC GTPase cycle | 0.000127 | 22 | Signal transduction |
| KEGG | GnRH signaling pathway | 0.00013 | 23 | Endocrine system |
| KEGG | Autophagy - animal | 0.000134 | 33 | Transport and catabolism |
| BioCarta | BIOCARTA TGFB PATHWAY | 0.000136 | 10 | Signal transduction |
| ACSN2 | SENESCENCE | 0.000144 | 25 | Cell growth and death |
| REACTOME | Beta-catenin independent WNT signaling | 0.000152 | 34 | Signal transduction |
| REACTOME | Clathrin-mediated endocytosis | 0.000176 | 34 | Transport and catabolism |
| KEGG | Hedgehog signaling pathway | 0.000183 | 18 | Signal transduction |
| REACTOME | MAPK1/MAPK3 signaling | 0.000228 | 55 | Signal transduction |
| BioCarta | BIOCARTA PDGF PATHWAY | 0.000233 | 12 | Signal transduction |
| HumanCyc | BMP Signalling Pathway | 0.000233 | 5 | Signal transduction |
| REACTOME | RAC3 GTPase cycle | 0.000249 | 25 | Signal transduction |
| WikiPathways | Corticotropin-releasing hormone signaling pathway | 0.000249 | 22 | Endocrine system |
| ACSN2 | TCR SIGNALING | 0.000269 | 42 | Immune system |
| KEGG | Ras signaling pathway | 0.00027 | 47 | Signal transduction |
| REACTOME | RND1 GTPase cycle | 0.000278 | 15 | Signal transduction |
| WikiPathways | Neurogenesis regulation in the olfactory epithelium | 0.000284 | 18 | Sensory system |
| BioCarta | BIOCARTA PPARA PATHWAY | 0.000287 | 17 | Endocrine system |

| | | | | |
|---|---|---|---|---|
| KEGG | Vasopressin-regulated water reabsorption | 0.00029 | 13 | Excretory system |
| PathBank | T Cell Receptor Signaling Pathway | 0.00029 | 16 | Immune system |
| WikiPathways | Vasopressin-regulated water reabsorption | 0.00029 | 13 | Excretory system |
| KEGG | Aldosterone synthesis and secretion | 0.000291 | 23 | Endocrine system |
| WikiPathways | CAMKK2 pathway | 0.000294 | 13 | Signal transduction |
| WikiPathways | White fat cell differentiation | 0.000294 | 13 | Immune system |
| REACTOME | Golgi-to-ER retrograde transport | 0.000299 | 31 | Transport and catabolism |
| KEGG | Neurotrophin signaling pathway | 0.000305 | 29 | Nervous system |
| KEGG | Hippo signaling pathway | 0.00031 | 35 | Signal transduction |
| REACTOME | Opioid Signalling | 0.000312 | 24 | Signal transduction |
| WikiPathways | PDGFR-beta pathway | 0.000314 | 12 | Signal transduction |
| ACSN2 | G2 M CHECKPOINT | 0.000319 | 22 | Cell growth and death |
| REACTOME | Signaling by MET | 0.000319 | 22 | Signal transduction |
| KEGG | Relaxin signaling pathway | 0.000334 | 28 | Endocrine system |
| BioCarta | BIOCARTA MTOR PATHWAY | 0.000341 | 10 | Signal transduction |
| REACTOME | CD209 (DC-SIGN) signaling | 0.000341 | 10 | Immune system |
| WikiPathways | Galanin receptor pathway | 0.000341 | 10 | Signal transduction |
| REACTOME | Signaling by NOTCH1 | 0.000342 | 21 | Signal transduction |
| KEGG | Gap junction | 0.000358 | 21 | Cellular community |
| PathBank | BCR Signaling Pathway | 0.000374 | 13 | Immune system |
| REACTOME | Oncogene Induced Senescence | 0.000374 | 13 | Cell growth and death |
| WikiPathways | Hepatocyte growth factor receptor signaling | 0.000374 | 13 | Signal transduction |
| WikiPathways | Hippo-Merlin signaling dysregulation | 0.000388 | 29 | Signal transduction |
| REACTOME | RAF/MAP kinase cascade | 0.000406 | 53 | Signal transduction |
| REACTOME | Non-integrin membrane-ECM interactions | 0.000414 | 18 | Cellular community |
| REACTOME | EPH-Ephrin signaling | 0.000417 | 24 | Development and regeneration |
| WikiPathways | ErbB signaling pathway | 0.000417 | 24 | Signal transduction |
| REACTOME | Signaling by Nuclear Receptors | 0.000426 | 51 | Signal transduction |
| PathBank | GnRH Signaling Pathway | 0.000436 | 16 | Endocrine system |
| KEGG | Growth hormone synthesis, secretion and action | 0.000437 | 26 | Endocrine system |
| REACTOME | RHOB GTPase cycle | 0.000438 | 20 | Signal transduction |
| BioCarta | BIOCARTA PPARG PATHWAY | 0.000447 | 6 | Endocrine system |
| WikiPathways | Leptin signaling pathway | 0.000476 | 21 | Signal transduction |
| REACTOME | Oxidative Stress Induced Senescence | 0.000477 | 24 | Cell growth and death |
| BioCarta | BIOCARTA MEF2D PATHWAY | 0.000477 | 9 | Immune system |
| REACTOME | RHOBTB GTPase Cycle | 0.000484 | 13 | Signal transduction |
| BioCarta | BIOCARTA CCR3 PATHWAY | 0.000487 | 7 | Immune system |
| WikiPathways | Development of ureteric collection system | 0.000494 | 18 | Development and regeneration |
| KEGG | cGMP-PKG signaling pathway | 0.000496 | 36 | Signal transduction |
| KEGG | Rap1 signaling pathway | 0.0005 | 40 | Signal transduction |
| WikiPathways | PDGF pathway | 0.000519 | 14 | Signal transduction |
| WikiPathways | DNA damage response (only ATM dependent) | 0.000571 | 27 | Cell growth and death |
| REACTOME | Nuclear Events (kinase and transcription factor activation) | 0.000614 | 18 | Signal transduction |
| REACTOME | trans-Golgi Network Vesicle Budding | 0.000628 | 20 | Transport and catabolism |
| BioCarta | BIOCARTA EGF PATHWAY | 0.000664 | 11 | Signal transduction |
| BioCarta | BIOCARTA PYK2 PATHWAY | 0.000664 | 11 | Signal transduction |
| WikiPathways | Ciliary landscape | 0.000675 | 43 | Transport and catabolism |
| REACTOME | TGF-beta receptor signaling activates SMADs | 0.000692 | 15 | Signal transduction |
| WikiPathways | DYRK1A | 0.000751 | 18 | Development and regeneration |
| Panther_Pathway | p53 pathway | 0.000809 | 21 | Cell growth and death |
| WikiPathways | Serotonin HTR1 group and FOS pathway | 0.000863 | 13 | Nervous system |
| PathBank | Dopamine Activation of Neurological Reward System | 0.000867 | 5 | Nervous system |
| REACTOME | Cell Cycle, Mitotic | 0.000889 | 86 | Cell growth and death |
| WikiPathways | Splicing factor NOVA regulated synaptic proteins | 0.000893 | 14 | Nervous system |
| KEGG | Oocyte meiosis | 0.000955 | 29 | Cell growth and death |
| REACTOME | Signaling by PDGF | 0.001004 | 17 | Signal transduction |
| WikiPathways | Gastrin signaling pathway | 0.001004 | 27 | Endocrine system |
| REACTOME | Cell Cycle | 0.001041 | 103 | Cell growth and death |
| BioCarta | BIOCARTA AGR PATHWAY | 0.001046 | 12 | Nervous system |
| REACTOME | Regulation of PTEN mRNA translation | 0.001046 | 6 | Signal transduction |
| BioCarta | BIOCARTA TPO PATHWAY | 0.001066 | 10 | Immune system |
| REACTOME | DARPP-32 events | 0.001066 | 10 | Signal transduction |
| SIGNOR 3.0 | Oxytocin signaling | 0.001066 | 10 | Endocrine system |
| REACTOME | Rap1 signalling | 0.001087 | 8 | Signal transduction |
| SIGNOR 3.0 | NOTCH Signaling | 0.001087 | 8 | Signal transduction |
| WikiPathways | Clock-controlled autophagy in bone metabolism | 0.001098 | 21 | Transport and catabolism |
| KEGG | Glutamatergic synapse | 0.001108 | 24 | Nervous system |
| REACTOME | Transmission across Chemical Synapses | 0.001111 | 50 | Nervous system |

| | | | | |
|---|---|---|---|---|
| WikiPathways | Thyroid hormones production and peripheral downstream signaling effects | 0.001142 | 23 | Metabolism of amino acids and derivatives |
| WikiPathways | Kit receptor signaling pathway | 0.001177 | 17 | Immune system |
| REACTOME | Neuronal System | 0.001186 | 69 | Nervous system |
| KEGG | Apelin signaling pathway | 0.001279 | 30 | Signal transduction |
| REACTOME | RAC2 GTPase cycle | 0.001314 | 22 | Signal transduction |
| ACSN2 | GROWTH FACTORS SIGNALING PATHWAYS | 0.001368 | 12 | Signal transduction |
| BioCarta | BIOCARTA FMLP PATHWAY | 0.001368 | 12 | Immune system |
| REACTOME | Pre-NOTCH Expression and Processing | 0.001455 | 20 | Signal transduction |
| BioCarta | BIOCARTA CARM ER PATHWAY | 0.001464 | 10 | Endocrine system |
| BioCarta | BIOCARTA CTCF PATHWAY | 0.001464 | 10 | Immune system |
| BioCarta | BIOCARTA PCAF PATHWAY | 0.00151 | 7 | Immune system |
| REACTOME | Mitotic Telophase/Cytokinesis | 0.00151 | 7 | Cell growth and death |
| KEGG | Longevity regulating pathway | 0.001521 | 22 | Aging |
| REACTOME | MAPK6/MAPK4 signaling | 0.001521 | 22 | Signal transduction |
| WikiPathways | Neural crest differentiation | 0.001524 | 24 | Development and regeneration |
| BioCarta | BIOCARTA IGF1 PATHWAY | 0.00161 | 9 | Endocrine system |
| BioCarta | BIOCARTA IL6 PATHWAY | 0.00161 | 9 | Immune system |
| SIGNOR 3.0 | MTOR Signaling | 0.00161 | 9 | Signal transduction |
| REACTOME | Activation of BH3-only proteins | 0.00163 | 11 | Cell growth and death |
| REACTOME | Signaling by ERBB2 | 0.001634 | 15 | Signal transduction |
| REACTOME | Hemostasis | 0.001635 | 105 | Immune system |
| PharmGKB | VEGF Signaling Pathway | 0.001677 | 17 | Signal transduction |
| Panther_Pathway | Insulin/IGF pathway-protein kinase B signaling cascade | 0.001739 | 12 | Endocrine system |
| ACSN2 | CORE | 0.001805 | 18 | Signal transduction |
| KEGG | Phosphatidylinositol signaling system | 0.001943 | 23 | Signal transduction |
| REACTOME | MET receptor recycling | 0.001951 | 6 | Signal transduction |
| SIGNOR 3.0 | Glucocorticoid receptor Signaling | 0.001951 | 6 | Endocrine system |
| PathBank | EGF Signalling Pathway | 0.001967 | 10 | Signal transduction |
| REACTOME | Downregulation of TGF-beta receptor signaling | 0.001967 | 10 | Signal transduction |
| KEGG | mTOR signaling pathway | 0.001968 | 32 | Signal transduction |
| REACTOME | Regulation of insulin secretion | 0.001973 | 19 | Energy metabolism |
| REACTOME | Rab regulation of trafficking | 0.00198 | 27 | Transport and catabolism |
| BioCarta | BIOCARTA KERATINOCYTE PATHWAY | 0.002107 | 14 | Development and regeneration |
| WikiPathways | LDLRAD4 and what we know about it | 0.002151 | 5 | Signal transduction |
| REACTOME | DAG and IP3 signaling | 0.002196 | 13 | Signal transduction |
| WikiPathways | Factors and pathways affecting insulin-like growth factor (IGF1)-Akt signaling | 0.002216 | 12 | Endocrine system |
| REACTOME | G alpha (12/13) signalling events | 0.002222 | 20 | Signal transduction |
| BioCarta | BIOCARTA CHREBP PATHWAY | 0.002246 | 6 | Signal transduction |
| REACTOME | SHC1 events in ERBB2 signaling | 0.002246 | 9 | Signal transduction |
| SIGNOR 3.0 | Insulin Signaling | 0.002246 | 9 | Endocrine system |
| KEGG | Insulin signaling pathway | 0.002252 | 29 | Endocrine system |
| REACTOME | PPARA activates gene expression | 0.0023 | 26 | Lipid metabolism |
| REACTOME | NCAM signaling for neurite out-growth | 0.002342 | 17 | Development and regeneration |
| BioCarta | BIOCARTA AGPCR PATHWAY | 0.002409 | 4 | Signal transduction |
| WikiPathways | SREBF and miR33 in cholesterol and lipid homeostasis | 0.002429 | 8 | Lipid metabolism |
| REACTOME | Cyclin D associated events in G1 | 0.002564 | 14 | Cell growth and death |
| REACTOME | G1 Phase | 0.002564 | 14 | Cell growth and death |
| BioCarta | BIOCARTA MAPK PATHWAY | 0.002565 | 20 | Signal transduction |
| REACTOME | EPHB-mediated forward signaling | 0.00271 | 13 | Development and regeneration |
| REACTOME | RHOF GTPase cycle | 0.00271 | 13 | Signal transduction |
| REACTOME | RND3 GTPase cycle | 0.00271 | 13 | Signal transduction |
| WikiPathways | IL-2 signaling pathway | 0.00271 | 13 | Immune system |
| REACTOME | Semaphorin interactions | 0.002757 | 17 | Development and regeneration |
| REACTOME | COPI-independent Golgi-to-ER retrograde traffic | 0.002862 | 15 | Transport and catabolism |
| KEGG | AMPK signaling pathway | 0.002894 | 26 | Signal transduction |
| REACTOME | Regulation of lipid metabolism by PPARalpha | 0.002894 | 26 | Lipid metabolism |
| REACTOME | Platelet activation, signaling and aggregation | 0.002909 | 47 | Immune system |
| WikiPathways | Prolactin signaling pathway | 0.002917 | 19 | Endocrine system |
| WikiPathways | Hair follicle development: cytodifferentiation - part 3 of 3 | 0.002919 | 20 | Development and regeneration |
| REACTOME | Negative regulation of the PI3K/AKT network | 0.002989 | 25 | Signal transduction |
| Panther_Pathway | B cell activation | 0.003027 | 16 | Immune system |
| REACTOME | PTEN Regulation | 0.003035 | 29 | Signal transduction |
| BioCarta | BIOCARTA HER2 PATHWAY | 0.003044 | 9 | Signal transduction |
| REACTOME | RHOBTB1 GTPase cycle | 0.003044 | 9 | Signal transduction |
| REACTOME | RHOBTB2 GTPase cycle | 0.003044 | 9 | Signal transduction |
| REACTOME | RHO GTPase Effectors | 0.003137 | 51 | Signal transduction |

| | | | | |
|---|---|---|---|---|
| KEGG | Inflammatory mediator regulation of TRP channels | 0.003158 | 20 | Sensory system |
| KEGG | Insulin secretion | 0.003318 | 18 | Endocrine system |
| REACTOME | G-protein mediated events | 0.003319 | 15 | Signal transduction |
| KEGG | Estrogen signaling pathway | 0.003322 | 26 | Endocrine system |
| REACTOME | CLEC7A (Dectin-1) induces NFAT activation | 0.003342 | 6 | Immune system |
| REACTOME | Establishment of Sister Chromatid Cohesion | 0.003342 | 6 | Cell growth and death |
| REACTOME | MET activates RAP1 and RAC1 | 0.003342 | 6 | Signal transduction |
| WikiPathways | Nuclear receptors | 0.003368 | 12 | Endocrine system |
| REACTOME | MyD88-independent TLR4 cascade | 0.003374 | 24 | Immune system |
| REACTOME | TRIF(TICAM1)-mediated TLR4 signaling | 0.003374 | 24 | Immune system |
| REACTOME | Cytokine Signaling in Immune system | 0.003378 | 107 | Immune system |
| REACTOME | RHO GTPases Activate ROCKs | 0.003382 | 8 | Signal transduction |
| REACTOME | Transcription of E2F targets under negative control by DREAM complex | 0.003382 | 8 | Cell growth and death |
| REACTOME | L1CAM interactions | 0.003464 | 26 | Development and regeneration |
| BioCarta | BIOCARTA PGC1A PATHWAY | 0.003561 | 7 | Energy metabolism |
| REACTOME | Antigen processing: Ubiquitination & Proteasome degradation | 0.003606 | 53 | Immune system |
| REACTOME | Class I MHC mediated antigen processing & presentation | 0.00362 | 63 | Immune system |
| REACTOME | Transcriptional regulation of white adipocyte differentiation | 0.003734 | 20 | Development and regeneration |
| REACTOME | TBC/RABGAPs | 0.003922 | 13 | Transport and catabolism |
| REACTOME | Regulation of cholesterol biosynthesis by SREBP (SREBF) | 0.003928 | 15 | Lipid metabolism |
| BioCarta | BIOCARTA WNT PATHWAY | 0.003997 | 9 | Signal transduction |
| REACTOME | Sema4D in semaphorin signaling | 0.003997 | 9 | Development and regeneration |
| SIGNOR 3.0 | WNT Signaling | 0.003997 | 9 | Signal transduction |
| SIGNOR 3.0 | WNT Signaling and Myogenesis | 0.003997 | 9 | Signal transduction |
| WikiPathways | Notch signaling pathway | 0.004124 | 16 | Signal transduction |
| REACTOME | Signaling by Interleukins | 0.004144 | 73 | Immune system |
| WikiPathways | Ras signaling | 0.004154 | 35 | Signal transduction |
| REACTOME | MET activates PTPN11 | 0.004167 | 4 | Signal transduction |
| PathBank | Intracellular Signalling Through Prostacyclin Receptor and Prostacyclin | 0.00417 | 5 | Signal transduction |
| REACTOME | Competing endogenous RNAs (ceRNAs) regulate PTEN translation | 0.00417 | 5 | Signal transduction |
| KEGG | ErbB signaling pathway | 0.004216 | 20 | Signal transduction |
| ACSN2 | G1 S CHECKPOINT | 0.004221 | 11 | Cell growth and death |
| WikiPathways | TROP2 regulatory signaling | 0.004255 | 14 | Signal transduction |
| KEGG | C-type lectin receptor signaling pathway | 0.004359 | 23 | Immune system |
| Panther_Pathway | FGF signaling pathway | 0.004359 | 23 | Signal transduction |
| WikiPathways | CKAP4 signaling pathway map | 0.004451 | 25 | Signal transduction |
| KEGG | Phospholipase D signaling pathway | 0.004464 | 27 | Signal transduction |
| REACTOME | p75 NTR receptor-mediated signalling | 0.004492 | 22 | Signal transduction |
| REACTOME | Programmed Cell Death | 0.004505 | 39 | Cell growth and death |
| KEGG | Endocrine and other factor-regulated calcium reabsorption | 0.004529 | 12 | Excretory system |
| BioCarta | BIOCARTA GSK3 PATHWAY | 0.004585 | 8 | Immune system |
| WikiPathways | BMP signaling in eyelid development | 0.004585 | 8 | Development and regeneration |
| BioCarta | BIOCARTA PITX2 PATHWAY | 0.005229 | 7 | Development and regeneration |
| REACTOME | Transcription of E2F targets under negative control by p107 (RBL1) and p130 (RBL2) in complex with HDAC1 | 0.005229 | 7 | Cell growth and death |
| BioCarta | BIOCARTA CARM1 PATHWAY | 0.005266 | 6 | Endocrine system |
| PathBank | Activation of cAMP-dependent protein kinase, PKA | 0.005266 | 5 | Signal transduction |
| WikiPathways | Bone morphogenic protein signaling and regulation | 0.005266 | 6 | Signal transduction |
| REACTOME | CaM pathway | 0.005281 | 11 | Signal transduction |
| REACTOME | Calmodulin induced events | 0.005281 | 11 | Signal transduction |
| WikiPathways | CCL18 signaling pathway | 0.005281 | 11 | Immune system |
| REACTOME | Signaling by VEGF | 0.005462 | 23 | Signal transduction |
| REACTOME | MAP kinase activation | 0.005538 | 16 | Immune system |
| Panther_Pathway | p53 pathway feedback loops 2 | 0.005606 | 13 | Cell growth and death |
| WikiPathways | Translation inhibitors in chronically activated PDGFRA cells | 0.005606 | 13 | Signal transduction |
| REACTOME | Activation of NMDA receptors and postsynaptic events | 0.005973 | 21 | Nervous system |
| REACTOME | MET promotes cell motility | 0.006169 | 12 | Signal transduction |
| REACTOME | Platelet homeostasis | 0.006186 | 19 | Immune system |
| BioCarta | BIOCARTA INSULIN PATHWAY | 0.006364 | 8 | Endocrine system |
| BioCarta | BIOCARTA NOS1 PATHWAY | 0.006364 | 8 | Signal transduction |
| PathBank | Cadmium Induces DNA Synthesis and Proliferation in Macrophages | 0.006364 | 8 | Immune system |
| KEGG | Platelet activation | 0.006392 | 23 | Immune system |
| WikiPathways | Target of rapamycin signaling | 0.006559 | 11 | Signal transduction |
| REACTOME | Adaptive Immune System | 0.006775 | 122 | Immune system |
| REACTOME | Activated NOTCH1 Transmits Signal to the Nucleus | 0.006803 | 10 | Signal transduction |
| REACTOME | MAPK targets/ Nuclear events mediated by MAP kinases | 0.006803 | 10 | Immune system |

| | | | | |
|---|---|---|---|---|
| HumanCyc | triacylglycerol biosynthesis | 0.006815 | 9 | Lipid metabolism |
| Panther_Pathway | Hypoxia response via HIF activation | 0.006815 | 9 | Signal transduction |
| REACTOME | MHC class II antigen presentation | 0.00703 | 25 | Immune system |
| REACTOME | Synthesis of PIPs at the plasma membrane | 0.007136 | 14 | Lipid metabolism |
| ACSN2 | GLUCOSE METABOLISM | 0.007171 | 24 | Carbohydrate metabolism |
| REACTOME | RHOA GTPase cycle | 0.00728 | 29 | Signal transduction |
| REACTOME | Integration of energy metabolism | 0.007281 | 22 | Energy metabolism |
| ACSN2 | DEATH RECEPTOR PATHWAYS | 0.007294 | 44 | Signal transduction |
| BioCarta | BIOCARTA CACAM PATHWAY | 0.007298 | 5 | Signal transduction |
| PathBank | Excitatory Neural Signalling Through 5-HTR 4 and Serotonin | 0.007298 | 5 | Nervous system |
| PathBank | Excitatory Neural Signalling Through 5-HTR 6 and Serotonin | 0.007298 | 5 | Nervous system |
| PathBank | Excitatory Neural Signalling Through 5-HTR 7 and Serotonin | 0.007298 | 5 | Nervous system |
| PathBank | Intracellular Signalling Through Histamine H2 Receptor and Histamine | 0.007298 | 5 | Signal transduction |
| REACTOME | Calcineurin activates NFAT | 0.007298 | 5 | Immune system |
| REACTOME | ROBO receptors bind AKAP5 | 0.007298 | 5 | Development and regeneration |
| UniProt_Pathways | Purine metabolism; 3',5'-cyclic AMP degradation; AMP from 3',5'-cyclic AMP: step 1/1. | 0.007298 | 5 | Nucleotide metabolism |
| ACSN2 | DNA DAMAGE RESPONSE | 0.007301 | 18 | Cell growth and death |
| REACTOME | Sensory processing of sound | 0.007301 | 18 | Sensory system |
| BioCarta | BIOCARTA CCR5 PATHWAY | 0.007302 | 7 | Immune system |
| BioCarta | BIOCARTA NDKDYNAMIN PATHWAY | 0.007302 | 7 | Nervous system |
| WikiPathways | Sterol regulatory element-binding proteins (SREBP) signaling | 0.007312 | 17 | Lipid metabolism |
| REACTOME | Activation of gene expression by SREBF (SREBP) | 0.007314 | 12 | Lipid metabolism |
| REACTOME | NCAM1 interactions | 0.007314 | 12 | Development and regeneration |
| REACTOME | MET activates STAT3 | 0.007323 | 3 | Signal transduction |
| WikiPathways | Lamin A-processing pathway | 0.007323 | 3 | Cell growth and death |
| REACTOME | Neurotransmitter receptors and postsynaptic signal transmission | 0.007452 | 37 | Nervous system |
| KEGG | Serotonergic synapse | 0.007669 | 21 | Nervous system |
| REACTOME | Ca-dependent events | 0.007718 | 11 | Signal transduction |
| BioCarta | BIOCARTA DREAM PATHWAY | 0.007723 | 6 | Nervous system |
| KEGG | Thyroid hormone synthesis | 0.008066 | 15 | Endocrine system |
| BioCarta | BIOCARTA CREB PATHWAY | 0.008074 | 8 | Signal transduction |
| REACTOME | NOTCH2 Activation and Transmission of Signal to the Nucleus | 0.008074 | 8 | Signal transduction |
| WikiPathways | Serotonin receptor 4/6/7 and NR3C signaling | 0.008074 | 5 | Nervous system |
| REACTOME | VEGFA-VEGFR2 Pathway | 0.008079 | 21 | Signal transduction |
| WikiPathways | Interferon type I signaling pathways | 0.008089 | 14 | Immune system |
| WikiPathways | Hair follicle development: organogenesis - part 2 of 3 | 0.008114 | 10 | Development and regeneration |
| REACTOME | Regulation of PTEN gene transcription | 0.008159 | 15 | Signal transduction |
| WikiPathways | AGE/RAGE pathway | 0.008178 | 16 | Immune system |
| BioCarta | BIOCARTA AT1R PATHWAY | 0.008282 | 9 | Signal transduction |
| REACTOME | G0 and Early G1 | 0.008282 | 9 | Cell growth and death |
| REACTOME | Syndecan interactions | 0.008282 | 9 | Cellular community |
| WikiPathways | Follicle stimulating hormone (FSH) signaling pathway | 0.008282 | 9 | Endocrine system |
| REACTOME | Protein-protein interactions at synapses | 0.008283 | 19 | Nervous system |
| REACTOME | Toll Like Receptor 3 (TLR3) Cascade | 0.008301 | 22 | Immune system |
| Panther_Pathway | Metabotropic glutamate receptor group II pathway | 0.008374 | 12 | Metabolism of amino acids and derivatives |
| WikiPathways | IL6 signaling pathway | 0.008374 | 12 | Immune system |
| REACTOME | Toll Like Receptor 4 (TLR4) Cascade | 0.008384 | 28 | Immune system |
| Panther_Pathway | Heterotrimeric G-protein signaling pathway-Gi alpha and Gs alpha mediated pathway | 0.008406 | 29 | Signal transduction |
| Panther_Pathway | Integrin signalling pathway | 0.008551 | 31 | Signal transduction |
| REACTOME | Interleukin-4 and Interleukin-13 signaling | 0.008735 | 23 | Immune system |
| PathBank | Intracellular Signalling Through PGD2 receptor and Prostaglandin D2 | 0.008795 | 4 | Signal transduction |
| REACTOME | MET activates PI3K/AKT signaling | 0.008795 | 4 | Signal transduction |
| REACTOME | PLC beta mediated events | 0.008816 | 13 | Signal transduction |
| WikiPathways | IL-3 signaling pathway | 0.008816 | 13 | Immune system |
| WikiPathways | Phosphoinositides metabolism | 0.008816 | 13 | Lipid metabolism |
| SIGNOR 3.0 | Thyroid Hormone Metabolism | 0.009091 | 11 | Metabolism of amino acids and derivatives |
| WikiPathways | G13 signaling pathway | 0.009091 | 11 | Signal transduction |
| WikiPathways | mBDNF and proBDNF regulation of GABA neurotransmission | 0.009091 | 11 | Nervous system |
| REACTOME | Cargo recognition for clathrin-mediated endocytosis | 0.009178 | 22 | Transport and catabolism |
| REACTOME | Toll-like Receptor Cascades | 0.009179 | 31 | Immune system |
| REACTOME | Apoptosis | 0.009192 | 33 | Cell growth and death |
| REACTOME | Activation of SMO | 0.009421 | 7 | Signal transduction |

| | | | | |
|---|---|---|---|---|
| REACTOME | Pre-NOTCH Processing in Golgi | 0.009421 | 7 | Signal transduction |
| WikiPathways | SRF and miRs in smooth muscle differentiation and proliferation | 0.009421 | 7 | Development and regeneration |
| Panther Pathway | Enkephalin release | 0.009742 | 10 | Endocrine system |
| REACTOME | Signaling by NOTCH2 | 0.009742 | 10 | Signal transduction |
| WikiPathways | Alpha 6 beta 4 signaling pathway | 0.009742 | 10 | Cellular community |

**Supplementary Table 6. Pathway annotation of cluster 1 endotype metabolites.** Metabolites were used to identify pathways they are annotated with in pathDIP.

| Compound | CHEBI ID | HMDB ID | Pathway Source | Pathway Name | Category |
|---|---|---|---|---|---|
| **Cluster 1** | | | | | |
| GLYCINE | 57305 | HMDB0000123 | REACTOME | Neurotransmitter receptors and postsynaptic signal transmission | Nervous system |
| GLYCINE | 57305 | HMDB0000123 | REACTOME | Transmission across Chemical Synapses | Nervous system |
| GLYCINE | 57305 | HMDB0000123 | REACTOME | Neuronal System | Nervous system |
| GLYCINE | 57305 | HMDB0000123 | REACTOME | Developmental Biology | Development and regeneration |
| GLYCINE | 57305 | HMDB0000123 | REACTOME | Glutathione conjugation | Metabolism of amino acids and derivatives |
| GLYCINE | 57305 | HMDB0000123 | REACTOME | Metabolism of nucleotides | Nucleotide metabolism |
| GLYCINE | 57305 | HMDB0000123 | REACTOME | Recycling of bile acids and salts | Lipid metabolism |
| GLYCINE | 57305 | HMDB0000123 | REACTOME | Signal Transduction | Signal transduction |
| GLYCINE | 57305 | HMDB0000123 | REACTOME | Glutathione synthesis and recycling | Metabolism of amino acids and derivatives |
| GLYCINE | 57305 | HMDB0000123 | REACTOME | Metabolism of porphyrins | Metabolism of cofactors and vitamins |
| GLYCINE | 57305 | HMDB0000123 | REACTOME | Heme biosynthesis | Metabolism of cofactors and vitamins |
| GLYCINE | 57305 | HMDB0000123 | REACTOME | Synthesis of bile acids and bile salts | Lipid metabolism |
| GLYCINE | 57305 | HMDB0000123 | REACTOME | Synthesis of bile acids and bile salts via 7alpha-hydroxycholesterol | Lipid metabolism |
| GLYCINE | 57305 | HMDB0000123 | REACTOME | Bile acid and bile salt metabolism | Lipid metabolism |
| GLYCINE | 57305 | HMDB0000123 | REACTOME | Metabolism of folate and pterines | Metabolism of cofactors and vitamins |
| GLYCINE | 57305 | HMDB0000123 | REACTOME | Metabolism of water-soluble vitamins and cofactors | Metabolism of cofactors and vitamins |
| GLYCINE | 57305 | HMDB0000123 | REACTOME | Metabolism of vitamins and cofactors | Metabolism of cofactors and vitamins |
| GLYCINE | 57305 | HMDB0000123 | REACTOME | Synthesis of Leukotrienes (LT) and Eoxins (EX) | Lipid metabolism |
| GLYCINE | 57305 | HMDB0000123 | REACTOME | Arachidonic acid metabolism | Lipid metabolism |
| GLYCINE | 57305 | HMDB0000123 | REACTOME | Metabolism of ingested SeMet, Sec, MeSec into H2Se | Metabolism of amino acids and derivatives |
| GLYCINE | 57305 | HMDB0000123 | REACTOME | Selenoamino acid metabolism | Metabolism of amino acids and derivatives |
| GLYCINE | 57305 | HMDB0000123 | REACTOME | EPH-Ephrin signaling | Development and regeneration |
| GLYCINE | 57305 | HMDB0000123 | REACTOME | Amino acid transport across the plasma membrane | Membrane transport |
| GLYCINE | 57305 | HMDB0000123 | REACTOME | Transport of small molecules | Membrane transport |
| GLYCINE | 57305 | HMDB0000123 | REACTOME | Glyoxylate metabolism and glycine degradation | Metabolism of amino acids and derivatives |
| GLYCINE | 57305 | HMDB0000123 | REACTOME | EPHB-mediated forward signaling | Development and regeneration |
| GLYCINE | 57305 | HMDB0000123 | REACTOME | Axon guidance | Development and regeneration |
| GLYCINE | 57305 | HMDB0000123 | REACTOME | Transport of bile salts and organic acids, metal ions and amine compounds | Membrane transport |
| GLYCINE | 57305 | HMDB0000123 | REACTOME | Transport of inorganic cations/anions and amino acids/oligopeptides | Membrane transport |
| GLYCINE | 57305 | HMDB0000123 | REACTOME | SLC-mediated transmembrane transport | Membrane transport |
| GLYCINE | 57305 | HMDB0000123 | REACTOME | Proton-coupled neutral amino acid transporters | Membrane transport |
| GLYCINE | 57305 | HMDB0000123 | REACTOME | Post NMDA receptor activation events | Nervous system |
| GLYCINE | 57305 | HMDB0000123 | REACTOME | Unblocking of NMDA receptors, glutamate binding and activation | Nervous system |
| GLYCINE | 57305 | HMDB0000123 | REACTOME | Na+/Cl- dependent neurotransmitter transporters | Membrane transport |
| GLYCINE | 57305 | HMDB0000123 | REACTOME | CREB1 phosphorylation through NMDA receptor-mediated activation of RAS signaling | Nervous system |
| GLYCINE | 57305 | HMDB0000123 | REACTOME | Activation of NMDA receptors and postsynaptic events | Nervous system |
| GLYCINE | 57305 | HMDB0000123 | REACTOME | Ras activation upon Ca2+ influx through NMDA receptor | Nervous system |
| GLYCINE | 57305 | HMDB0000123 | REACTOME | Metabolism of lipids | Lipid metabolism |
| GLYCINE | 57305 | HMDB0000123 | REACTOME | RAF/MAP kinase cascade | Signal transduction |
| GLYCINE | 57305 | HMDB0000123 | REACTOME | MAPK family signaling cascades | Signal transduction |
| GLYCINE | 57305 | HMDB0000123 | REACTOME | MAPK1/MAPK3 signaling | Signal transduction |
| GLYCINE | 57305 | HMDB0000123 | REACTOME | Glycine degradation | Metabolism of amino acids and derivatives |
| GLYCINE | 57305 | HMDB0000123 | REACTOME | Protein-protein interactions at synapses | Nervous system |
| GLYCINE | 57305 | HMDB0000123 | REACTOME | Choline catabolism | Metabolism of amino acids and derivatives |
| GLYCINE | 57305 | HMDB0000123 | REACTOME | Carnitine synthesis | Metabolism of amino acids and derivatives |

| | | | | | |
|---|---|---|---|---|---|
| GLYCINE | 57305 | HMDB0000123 | REACTOME | Creatine metabolism | Metabolism of amino acids and derivatives |
| GLYCINE | 57305 | HMDB0000123 | REACTOME | Metabolism of amino acids and derivatives | Metabolism of amino acids and derivatives |
| GLYCINE | 57305 | HMDB0000123 | REACTOME | Purine ribonucleoside monophosphate biosynthesis | Nucleotide metabolism |
| GLYCINE | 57305 | HMDB0000123 | REACTOME | Threonine catabolism | Metabolism of amino acids and derivatives |
| GLYCINE | 57305 | HMDB0000123 | REACTOME | Synaptic adhesion-like molecules | Nervous system |
| GLYCINE | 57305 | HMDB0000123 | REACTOME | Nucleotide biosynthesis | Nucleotide metabolism |
| GLYCINE | 57305 | HMDB0000123 | REACTOME | Metabolism of steroids | Lipid metabolism |
| GLYCINE | 57305 | HMDB0000123 | REACTOME | Fatty acid metabolism | Lipid metabolism |
| GLYCINE | 57305 | HMDB0000123 | REACTOME | Biosynthesis of DHA-derived SPMs | Lipid metabolism |
| GLYCINE | 57305 | HMDB0000123 | REACTOME | Biosynthesis of specialized proresolving mediators (SPMs) | Lipid metabolism |
| GLYCINE | 57305 | HMDB0000123 | REACTOME | Biosynthesis of DHA-derived sulfido conjugates | Lipid metabolism |
| GLYCINE | 57305 | HMDB0000123 | REACTOME | Biosynthesis of maresin conjugates in tissue regeneration (MCTR) | Lipid metabolism |
| GLYCINE | 57305 | HMDB0000123 | REACTOME | Biosynthesis of protectin and resolvin conjugates in tissue regeneration (PCTR and RCTR) | Lipid metabolism |
| GLYCINE | 57305 | HMDB0000123 | REACTOME | Negative regulation of NMDA receptor-mediated neuronal transmission | Nervous system |
| GLYCINE | 57305 | HMDB0000123 | REACTOME | Long-term potentiation | Nervous system |
| GLYCINE | 57305 | HMDB0000123 | REACTOME | Nervous system development | Development and regeneration |
| GLYCINE | 57305 | HMDB0000123 | SIGNOR 3.0 | Carnitine biosynthesis | Lipid metabolism |

**Supplementary Table 7. Pathway annotation of cluster 2 endotype metabolites.** Metabolites were used to identify pathways they are annotated with in pathDIP.

| Compound | CHEBI ID | HMDB ID | Pathway Source | Pathway Name | Category |
|---|---|---|---|---|---|
| **Cluster 2** | | | | | |
| ACETYLORNITHINE | 16543 | HMDB0003357 | KEGG | Arginine biosynthesis | Metabolism of amino acids and derivatives |
| ACETYLORNITHINE | 16543 | HMDB0003357 | MetabolicAtlas | arginine_and_proline_metabolism | Metabolism of amino acids and derivatives |
| ACETYLORNITHINE | 16543 | HMDB0003357 | MetabolicAtlas | transport_reactions | Membrane transport |

**Supplementary Table 8. Pathway annotation of cluster 3 endotype metabolites.** Metabolites were used to identify pathways they are annotated with in pathDIP.

| Compound | CHEBI ID | HMDB ID | Pathway Source | Pathway Name | Category |
|---|---|---|---|---|---|
| **Cluster 3** | | | | | |
| LYSOPHOSPHATIDYLCHOLINE | 17504 | HMDB0010397 | KEGG | Glycerophospholipid metabolism | Lipid metabolism |
| LYSOPHOSPHATIDYLCHOLINE | 16728 | | KEGG | Glycerophospholipid metabolism | Lipid metabolism |
| LYSOPHOSPHATIDYLCHOLINE | 16728 | HMDB0062192 | REACTOME | Acyl chain remodelling of PC | Lipid metabolism |
| LYSOPHOSPHATIDYLCHOLINE | 16728 | HMDB0062192 | REACTOME | Hydrolysis of LPC | Lipid metabolism |
| LYSOPHOSPHATIDYLCHOLINE | 16728 | HMDB0062192 | REACTOME | Glycerophospholipid biosynthesis | Lipid metabolism |
| LYSOPHOSPHATIDYLCHOLINE | 16728 | HMDB0062192 | REACTOME | Phospholipid metabolism | Lipid metabolism |
| LYSOPHOSPHATIDYLCHOLINE | 16728 | HMDB0062192 | REACTOME | Metabolism of lipids | Lipid metabolism |
| LYSOPHOSPHATIDYLCHOLINE | 17504 | HMDB0010397 | REACTOME | Opioid Signalling | Signal transduction |
| LYSOPHOSPHATIDYLCHOLINE | 17504 | HMDB0010397 | REACTOME | phospho-PLA2 pathway | Signal transduction |
| LYSOPHOSPHATIDYLCHOLINE | 17504 | HMDB0010397 | REACTOME | Ca-dependent events | Signal transduction |
| LYSOPHOSPHATIDYLCHOLINE | 17504 | HMDB0010397 | REACTOME | G-protein mediated events | Signal transduction |
| LYSOPHOSPHATIDYLCHOLINE | 17504 | HMDB0010397 | REACTOME | PLC beta mediated events | Signal transduction |
| LYSOPHOSPHATIDYLCHOLINE | 17504 | HMDB0010397 | REACTOME | Acyl chain remodelling of PC | Lipid metabolism |
| LYSOPHOSPHATIDYLCHOLINE | 17504 | HMDB0010397 | REACTOME | Acyl chain remodeling of CL | Lipid metabolism |
| LYSOPHOSPHATIDYLCHOLINE | 17504 | HMDB0010397 | REACTOME | Hydrolysis of LPC | Lipid metabolism |
| LYSOPHOSPHATIDYLCHOLINE | 17504 | HMDB0010397 | REACTOME | Glycerophospholipid biosynthesis | Lipid metabolism |
| LYSOPHOSPHATIDYLCHOLINE | 17504 | HMDB0010397 | REACTOME | Phospholipid metabolism | Lipid metabolism |
| LYSOPHOSPHATIDYLCHOLINE | 17504 | HMDB0010397 | REACTOME | Signal Transduction | Signal transduction |
| LYSOPHOSPHATIDYLCHOLINE | 17504 | HMDB0010397 | REACTOME | Innate Immune System | Immune system |
| LYSOPHOSPHATIDYLCHOLINE | 17504 | HMDB0010397 | REACTOME | Immune System | Immune system |
| LYSOPHOSPHATIDYLCHOLINE | 17504 | HMDB0010397 | REACTOME | Plasma lipoprotein assembly, remodeling, and clearance | Membrane transport |
| LYSOPHOSPHATIDYLCHOLINE | 17504 | HMDB0010397 | REACTOME | Membrane Trafficking | Transport and catabolism |
| LYSOPHOSPHATIDYLCHOLINE | 17504 | HMDB0010397 | REACTOME | Fcgamma receptor (FCGR) dependent phagocytosis | Immune system |
| LYSOPHOSPHATIDYLCHOLINE | 17504 | HMDB0010397 | REACTOME | Role of phospholipids in phagocytosis | Immune system |
| LYSOPHOSPHATIDYLCHOLINE | 17504 | HMDB0010397 | REACTOME | Arachidonic acid metabolism | Lipid metabolism |
| LYSOPHOSPHATIDYLCHOLINE | 17504 | HMDB0010397 | REACTOME | Signaling by GPCR | Signal transduction |
| LYSOPHOSPHATIDYLCHOLINE | 17504 | HMDB0010397 | REACTOME | Transport of small molecules | Membrane transport |
| LYSOPHOSPHATIDYLCHOLINE | 17504 | HMDB0010397 | REACTOME | GPCR downstream signalling | Signal transduction |
| LYSOPHOSPHATIDYLCHOLINE | 17504 | HMDB0010397 | REACTOME | G alpha (i) signalling events | Signal transduction |
| LYSOPHOSPHATIDYLCHOLINE | 17504 | HMDB0010397 | REACTOME | Metabolism of lipids | Lipid metabolism |
| LYSOPHOSPHATIDYLCHOLINE | 17504 | HMDB0010397 | REACTOME | Vesicle-mediated transport | Transport and catabolism |
| LYSOPHOSPHATIDYLCHOLINE | 17504 | HMDB0010397 | REACTOME | COPI-independent Golgi-to-ER retrograde traffic | Transport and catabolism |
| LYSOPHOSPHATIDYLCHOLINE | 17504 | HMDB0010397 | REACTOME | Intra-Golgi and retrograde Golgi-to-ER traffic | Transport and catabolism |
| LYSOPHOSPHATIDYLCHOLINE | 17504 | HMDB0010397 | REACTOME | Golgi-to-ER retrograde transport | Transport and catabolism |
| LYSOPHOSPHATIDYLCHOLINE | 17504 | HMDB0010397 | REACTOME | Plasma lipoprotein remodeling | Membrane transport |
| LYSOPHOSPHATIDYLCHOLINE | 17504 | HMDB0010397 | REACTOME | HDL remodeling | Membrane transport |
| LYSOPHOSPHATIDYLCHOLINE | 17504 | HMDB0010397 | REACTOME | Fatty acid metabolism | Lipid metabolism |
| PHOSPHATIDYLCHOLINE | 57643 | | REACTOME | Opioid Signalling | Signal transduction |
| PHOSPHATIDYLCHOLINE | 57643 | | REACTOME | phospho-PLA2 pathway | Signal transduction |
| PHOSPHATIDYLCHOLINE | 57643 | | REACTOME | Ca-dependent events | Signal transduction |
| PHOSPHATIDYLCHOLINE | 57643 | | REACTOME | G-protein mediated events | Signal transduction |
| PHOSPHATIDYLCHOLINE | 57643 | | REACTOME | PLC beta mediated events | Signal transduction |
| PHOSPHATIDYLCHOLINE | 57643 | | REACTOME | Cytokine Signaling in Immune system | Immune system |
| PHOSPHATIDYLCHOLINE | 57643 | | REACTOME | ABC transporters in lipid homeostasis | Membrane transport |
| PHOSPHATIDYLCHOLINE | 57643 | | REACTOME | Acyl chain remodelling of PC | Lipid metabolism |
| PHOSPHATIDYLCHOLINE | 57643 | | REACTOME | Acyl chain remodeling of CL | Lipid metabolism |
| PHOSPHATIDYLCHOLINE | 57643 | | REACTOME | Acyl chain remodelling of PE | Lipid metabolism |
| PHOSPHATIDYLCHOLINE | 57643 | | REACTOME | Synthesis of PS | Lipid metabolism |
| PHOSPHATIDYLCHOLINE | 57643 | | REACTOME | Synthesis of PG | Lipid metabolism |
| PHOSPHATIDYLCHOLINE | 57643 | | REACTOME | Synthesis of PA | Lipid metabolism |
| PHOSPHATIDYLCHOLINE | 57643 | | REACTOME | Synthesis of PC | Lipid metabolism |

| | | | | | |
|---|---|---|---|---|---|
| PHOSPHATIDYLCHOLINE | 57643 | | REACTOME | PI and PC transport between ER and Golgi membranes | Lipid metabolism |
| PHOSPHATIDYLCHOLINE | 57643 | | REACTOME | Glycerophospholipid biosynthesis | Lipid metabolism |
| PHOSPHATIDYLCHOLINE | 57643 | | REACTOME | PI Metabolism | Lipid metabolism |
| PHOSPHATIDYLCHOLINE | 57643 | | REACTOME | Phospholipid metabolism | Lipid metabolism |
| PHOSPHATIDYLCHOLINE | 57643 | | REACTOME | Signal Transduction | Signal transduction |
| PHOSPHATIDYLCHOLINE | 57643 | | REACTOME | Sphingolipid de novo biosynthesis | Lipid metabolism |
| PHOSPHATIDYLCHOLINE | 57643 | | REACTOME | Innate Immune System | Immune system |
| PHOSPHATIDYLCHOLINE | 57643 | | REACTOME | Immune System | Immune system |
| PHOSPHATIDYLCHOLINE | 57643 | | REACTOME | Plasma lipoprotein assembly, remodeling, and clearance | Membrane transport |
| PHOSPHATIDYLCHOLINE | 57643 | | REACTOME | Membrane Trafficking | Transport and catabolism |
| PHOSPHATIDYLCHOLINE | 57643 | | REACTOME | Fcgamma receptor (FCGR) dependent phagocytosis | Immune system |
| PHOSPHATIDYLCHOLINE | 57643 | | REACTOME | Role of phospholipids in phagocytosis | Immune system |
| PHOSPHATIDYLCHOLINE | 57643 | | REACTOME | Arachidonic acid metabolism | Lipid metabolism |
| PHOSPHATIDYLCHOLINE | 57643 | | REACTOME | Signaling by GPCR | Signal transduction |
| PHOSPHATIDYLCHOLINE | 57643 | | REACTOME | Transport of small molecules | Membrane transport |
| PHOSPHATIDYLCHOLINE | 57643 | | REACTOME | ABC-family proteins mediated transport | Membrane transport |
| PHOSPHATIDYLCHOLINE | 57643 | | REACTOME | GPCR downstream signalling | Signal transduction |
| PHOSPHATIDYLCHOLINE | 57643 | | REACTOME | G alpha (i) signalling events | Signal transduction |
| PHOSPHATIDYLCHOLINE | 57643 | | REACTOME | Sphingolipid metabolism | Lipid metabolism |
| PHOSPHATIDYLCHOLINE | 57643 | | REACTOME | Metabolism of lipids | Lipid metabolism |
| PHOSPHATIDYLCHOLINE | 57643 | | REACTOME | Vesicle-mediated transport | Transport and catabolism |
| PHOSPHATIDYLCHOLINE | 57643 | | REACTOME | COPI-independent Golgi-to-ER retrograde traffic | Transport and catabolism |
| PHOSPHATIDYLCHOLINE | 57643 | | REACTOME | Intra-Golgi and retrograde Golgi-to-ER traffic | Transport and catabolism |
| PHOSPHATIDYLCHOLINE | 57643 | | REACTOME | Golgi-to-ER retrograde transport | Transport and catabolism |
| PHOSPHATIDYLCHOLINE | 57643 | | REACTOME | Plasma lipoprotein remodeling | Membrane transport |
| PHOSPHATIDYLCHOLINE | 57643 | | REACTOME | HDL remodeling | Membrane transport |
| PHOSPHATIDYLCHOLINE | 57643 | | REACTOME | Fatty acid metabolism | Lipid metabolism |
| PHOSPHATIDYLCHOLINE | 57643 | | REACTOME | Signaling by CSF1 (M-CSF) in myeloid cells | Immune system |
| LYSOPHOSPHATIDYLCHOLINE | 60479 | | REACTOME | Hydrolysis of LPC | Lipid metabolism |
| LYSOPHOSPHATIDYLCHOLINE | 60479 | | REACTOME | Glycerophospholipid biosynthesis | Lipid metabolism |
| LYSOPHOSPHATIDYLCHOLINE | 60479 | | REACTOME | PI Metabolism | Lipid metabolism |
| LYSOPHOSPHATIDYLCHOLINE | 60479 | | REACTOME | Phospholipid metabolism | Lipid metabolism |
| LYSOPHOSPHATIDYLCHOLINE | 60479 | | REACTOME | Innate Immune System | Immune system |
| LYSOPHOSPHATIDYLCHOLINE | 60479 | | REACTOME | Immune System | Immune system |
| LYSOPHOSPHATIDYLCHOLINE | 60479 | | REACTOME | Toll-like Receptor Cascades | Immune system |
| LYSOPHOSPHATIDYLCHOLINE | 60479 | | REACTOME | Binding and Uptake of Ligands by Scavenger Receptors | Transport and catabolism |
| LYSOPHOSPHATIDYLCHOLINE | 60479 | | REACTOME | Scavenging by Class B Receptors | Transport and catabolism |
| LYSOPHOSPHATIDYLCHOLINE | 60479 | | REACTOME | Scavenging by Class A Receptors | Transport and catabolism |
| LYSOPHOSPHATIDYLCHOLINE | 60479 | | REACTOME | Scavenging by Class F Receptors | Transport and catabolism |
| LYSOPHOSPHATIDYLCHOLINE | 60479 | | REACTOME | Metabolism of lipids | Lipid metabolism |
| LYSOPHOSPHATIDYLCHOLINE | 60479 | | REACTOME | Vesicle-mediated transport | Transport and catabolism |
| LYSOPHOSPHATIDYLCHOLINE | 60479 | | REACTOME | Regulation of TLR by endogenous ligand | Immune system |
| LYSOPHOSPHATIDYLCHOLINE | 60479 | | REACTOME | Glycerophospholipid catabolism | Lipid metabolism |
| LYSOPHOSPHATIDYLCHOLINE | 64483 | | REACTOME | Synthesis of PC | Lipid metabolism |
| LYSOPHOSPHATIDYLCHOLINE | 64483 | | REACTOME | Glycerophospholipid biosynthesis | Lipid metabolism |
| LYSOPHOSPHATIDYLCHOLINE | 64483 | | REACTOME | Phospholipid metabolism | Lipid metabolism |
| LYSOPHOSPHATIDYLCHOLINE | 64483 | | REACTOME | Metabolism of lipids | Lipid metabolism |
| LYSOPHOSPHATIDYLCHOLINE | 64489 | HMDB0010379 | PathBank | Arachidonic Acid Metabolism | Lipid metabolism |

| | | | | | |
|---|---|---|---|---|---|
| LYSOPHOSPHATIDYLCHOLINE | 72998 | HMDB0010382 | PathBank | Phospholipid Biosynthesis | Lipid metabolism |
| PHOSPHATIDYLCHOLINE | 72999 | HMDB0000564 | PathBank | Phospholipid Biosynthesis | Lipid metabolism |

**Supplementary Table 9. Mean Area Under the Curve (AUC) Comparison of Machine Learning Approaches applied individually to Metabolite plasma and miRNA Domains.** This table displays the mean Area Under the Curve (AUC) values obtained while classifying WOMAC pain and function responses using logistic regression, lasso regression, ridge regression, support vector machines, and random forests. Note the consistent higher AUC (in bold) provided by random forests highlighting its efficacy in response classification modeling for metabolite and miRNA data.

| **Outcome** | **ML models** | **Metabolite plasma** | **miRNA plasma** | **miRNA synovial fluid** | **miRNA urine** | **Clinical** |
|---|---|---|---|---|---|---|
| | | **mean AUC** | **mean AUC** | **mean AUC** | **mean AUC** | **mean AUC** |
| **WOMAC pain response** | Random Forest | **0.623** | **0.691** | **0.687** | **0.692** | **0.7** |
| | Logistic Regression | 0.606 | 0.674 | 0.67 | 0.675 | 0.683 |
| | Lasso Regression | 0.612 | 0.68 | 0.676 | 0.681 | 0.689 |
| | Ridge Regression | 0.609 | 0.672 | 0.673 | 0.678 | 0.686 |
| | Support Vector Machines | 0.61 | 0.678 | 0.673 | 0.673 | 0.655 |
| **WOMAC function response** | Random Forest | **0.536** | **0.652** | **0.644** | **0.586** | **0.738** |
| | Logistic Regression | 0.521 | 0.637 | 0.629 | 0.571 | 0.723 |
| | Lasso Regression | 0.524 | 0.64 | 0.632 | 0.574 | 0.726 |
| | Ridge Regression | 0.526 | 0.641 | 0.638 | 0.557 | 0.711 |
| | Support Vector Machines | 0.517 | 0.632 | 0.629 | 0.548 | 0.714 |

**Supplementary Table 10. Cluster-wise Variable Selection Criteria for Unimodal Machine Learning Models classifying WOMAC pain response.** Variables within multiomic data domains were selected based on criteria: absolute(log fold change) > 1.1 for plasma metabolites and > 1.5 for miRNAs. These criteria ensured the inclusion of biologically significant features for further analysis. Subsequently, selected variables were integrated into the integrative machine learning framework to classify WOMAC pain response within each cluster.

| **Cluster 1 vs 2 and 3** | | |
|---|---|---|
| **Multiomic data domain** | **Features** | **logFold Change** |
| **Metabolites plasma** | alpha.Aminoadipicacid_pla | 0.183 |
| | trans.hydroxyproline_pla | 0.224 |
| | Citruline_pla | -0.161 |
| **miRNA plasma** | hsa-miR-4429_pla | 0.819 |
| | hsa-miR-208b-3p_pla | -0.594 |
| **miRNA synovial fluid** | hsa-miR-5698_syn | 1.206 |
| | hsa-miR-6758-3p_syn | 1.005 |
| | hsa-miR-1273h-5p_syn | -0.737 |
| | hsa-let-7f-1-3p_syn | -0.761 |
| | hsa-miR-2116-5p_syn | 0.904 |
| | hsa-miR-1251-5p_syn | 0.785 |
| | hsa-miR-6799-5p_syn | 0.879 |
| | hsa-miR-4688_syn | 0.874 |
| | hsa-miR-654-5p_syn | 0.893 |
| | hsa-miR-3150a-5p_syn | 0.849 |
| | hsa-miR-6837-3p_syn | 0.807 |
| | hsa-miR-548ax_syn | 0.854 |
| | hsa-miR-181d-3p_syn | 0.869 |
| | hsa-miR-873-5p_syn | 0.899 |
| | hsa-miR-619-5p_syn | 0.866 |
| | hsa-miR-3918_syn | 0.863 |
| | hsa-miR-1227-5p_syn | 0.863 |
| | hsa-miR-6729-5p_syn | 0.855 |
| | hsa-miR-4717-3p_syn | 0.825 |
| | hsa-miR-378d_syn | 0.823 |
| | hsa-miR-4670-5p_syn | 0.826 |
| | hsa-miR-4724-5p_syn | 0.834 |
| | hsa-miR-508-3p_syn | 0.801 |
| | hsa-miR-6820-3p_syn | 0.836 |
| | hsa-miR-12116_syn | 0.866 |
| | hsa-miR-4687-5p_syn | 0.829 |
| | hsa-miR-6871-5p_syn | 0.83 |
| | hsa-miR-3164_syn | 0.779 |
| | hsa-miR-4659b-5p_syn | 0.785 |
| | hsa-miR-129-5p_syn | 0.827 |
| | hsa-miR-4785_syn | 0.774 |
| | hsa-miR-4649-5p_syn | 0.786 |
| | hsa-miR-4689_syn | 0.801 |
| | hsa-miR-6730-3p_syn | 0.825 |
| | hsa-miR-548b-5p_syn | 0.799 |
| | hsa-miR-4776-3p_syn | 0.849 |
| | hsa-miR-4516_syn | 0.844 |
| | hsa-miR-6721-5p_syn | 0.83 |
| | hsa-miR-6762-3p_syn | 0.848 |
| | hsa-miR-93-5p_syn | 0.805 |
| | hsa-miR-145-3p_syn | -1.104 |
| | hsa-miR-3201_syn | -0.643 |
| | hsa-miR-711_syn | 0.805 |
| | hsa-miR-5571-3p_syn | 0.79 |
| | hsa-miR-378i_syn | 0.823 |
| | hsa-miR-6511a-3p_syn | 0.803 |
| | hsa-miR-1301-3p_syn | 0.801 |
| | hsa-miR-4429_syn | 0.868 |
| | hsa-miR-4664-3p_syn | 0.791 |
| | hsa-miR-122b-3p_syn | 0.767 |
| | hsa-miR-1306-5p_syn | 0.778 |
| | hsa-miR-6818-3p_syn | 0.736 |
| | hsa-miR-196b-3p_syn | 0.734 |
| | hsa-miR-6843-3p_syn | 0.757 |
| | hsa-miR-5704_syn | 0.757 |

| | | |
|---|---|---|
| | hsa-miR-3529-5p_syn | 0.77 |
| | hsa-miR-1236-5p_syn | 0.767 |
| | hsa-miR-6769b-3p_syn | 0.75 |
| | hsa-miR-106a-3p_syn | 0.755 |
| | hsa-miR-4715-3p_syn | 0.782 |
| | hsa-miR-3158-3p_syn | 0.694 |
| | hsa-miR-3174_syn | 0.677 |
| | hsa-miR-548u_syn | 0.753 |
| | hsa-miR-4703-3p_syn | 0.732 |
| | hsa-miR-488-3p_syn | 0.744 |
| | hsa-miR-6810-3p_syn | 0.746 |
| | hsa-miR-3085-5p_syn | 0.76 |
| | hsa-miR-557_syn | 0.74 |
| | hsa-miR-4714-3p_syn | 0.763 |
| | hsa-miR-10525-3p_syn | 0.682 |
| | hsa-miR-187-5p_syn | 0.769 |
| | hsa-miR-10395-3p_syn | 0.742 |
| | hsa-miR-3126-5p_syn | 0.743 |
| | hsa-miR-3667-3p_syn | 0.704 |
| | hsa-miR-6509-5p_syn | 0.77 |
| | hsa-miR-19b-1-5p_syn | 0.727 |
| | hsa-miR-6748-5p_syn | 0.709 |
| | hsa-miR-4716-3p_syn | 0.71 |
| | hsa-miR-4680-3p_syn | 0.731 |
| | hsa-miR-548w_syn | 0.718 |
| | hsa-miR-5191_syn | 0.708 |
| | hsa-miR-4286_syn | 0.647 |
| | hsa-miR-938_syn | 0.72 |
| | hsa-miR-30b-3p_syn | 0.68 |
| | hsa-miR-615-5p_syn | 0.716 |
| | hsa-miR-6785-5p_syn | 0.725 |
| | hsa-miR-4449_syn | 0.744 |
| | hsa-miR-4687-3p_syn | 0.676 |
| | hsa-miR-4677-5p_syn | 0.718 |
| | hsa-miR-4511_syn | 0.69 |
| | hsa-miR-6763-3p_syn | 0.676 |
| | hsa-miR-3150b-3p_syn | 0.692 |
| | hsa-miR-451a_syn | -1.041 |
| | hsa-miR-148a-3p_syn | 0.728 |
| | hsa-miR-6873-3p_syn | 0.62 |
| | hsa-miR-5091_syn | 0.708 |
| | hsa-miR-4709-3p_syn | 0.698 |
| | hsa-miR-511-3p_syn | 0.689 |
| | hsa-miR-5193_syn | 0.701 |
| | hsa-miR-6826-3p_syn | 0.701 |
| | hsa-miR-3939_syn | 0.684 |
| | hsa-miR-6740-5p_syn | 0.662 |
| | hsa-miR-2117_syn | 0.711 |
| | hsa-miR-6766-3p_syn | 0.676 |
| | hsa-miR-7113-3p_syn | 0.685 |
| | hsa-miR-2682-5p_syn | 0.691 |
| | hsa-miR-1199-5p_syn | 0.651 |
| | hsa-miR-877-3p_syn | 0.69 |
| | hsa-miR-6810-5p_syn | 0.694 |
| | hsa-miR-3613-5p_syn | 0.715 |
| | hsa-miR-6826-5p_syn | 0.689 |
| | hsa-miR-3192-5p_syn | 0.658 |
| | hsa-miR-6802-5p_syn | 0.676 |
| | hsa-miR-3616-3p_syn | 0.667 |
| | hsa-miR-7702_syn | 0.648 |
| | hsa-miR-5699-3p_syn | 0.684 |
| | hsa-miR-6787-3p_syn | 0.687 |
| | hsa-miR-7847-3p_syn | 0.676 |
| | hsa-miR-539-5p_syn | 0.663 |
| | hsa-miR-6732-3p_syn | 0.69 |
| | hsa-miR-6800-3p_syn | 0.634 |
| | hsa-miR-3139_syn | 0.704 |
| | hsa-miR-100-5p_syn | 0.673 |
| | hsa-miR-345-3p_syn | 0.663 |
| | hsa-miR-4715-5p_syn | 0.656 |
| | hsa-miR-4727-3p_syn | 0.674 |

| | | |
|---|---|---|
| | hsa-miR-6796-5p_syn | 0.698 |
| | hsa-miR-4742-5p_syn | 0.632 |
| | hsa-miR-6767-5p_syn | 0.637 |
| | hsa-miR-6754-5p_syn | 0.666 |
| | hsa-miR-6755-3p_syn | 0.669 |
| | hsa-miR-2114-3p_syn | 0.692 |
| | hsa-miR-6812-3p_syn | 0.672 |
| | hsa-miR-887-5p_syn | 0.648 |
| | hsa-miR-2116-3p_syn | 0.651 |
| | hsa-miR-582-5p_syn | 0.644 |
| | hsa-miR-6842-5p_syn | 0.634 |
| | hsa-miR-6866-3p_syn | 0.622 |
| | hsa-miR-3667-5p_syn | 0.639 |
| | hsa-miR-4467_syn | 0.69 |
| | hsa-miR-4670-3p_syn | 0.646 |
| | hsa-miR-4645-3p_syn | 0.613 |
| | hsa-miR-4714-5p_syn | 0.652 |
| | hsa-miR-3911_syn | 0.645 |
| | hsa-miR-4632-5p_syn | 0.666 |
| | hsa-miR-2681-3p_syn | 0.643 |
| | hsa-miR-4489_syn | 0.607 |
| | hsa-miR-6882-3p_syn | 0.632 |
| | hsa-miR-19a-5p_syn | 0.647 |
| | hsa-miR-3181_syn | 0.621 |
| | hsa-miR-4526_syn | 0.596 |
| | hsa-miR-4757-5p_syn | 0.629 |
| | hsa-miR-6753-3p_syn | 0.624 |
| | hsa-miR-6883-3p_syn | 0.629 |
| | hsa-miR-1305_syn | 0.622 |
| | hsa-miR-6876-5p_syn | 0.629 |
| | hsa-miR-4802-5p_syn | 0.595 |
| | hsa-miR-7156-3p_syn | 0.624 |
| | hsa-miR-6771-5p_syn | 0.621 |
| | hsa-miR-4669_syn | 0.646 |
| | hsa-miR-618_syn | 0.634 |
| | hsa-miR-4762-5p_syn | 0.65 |
| | hsa-miR-548t-5p_syn | 0.612 |
| | hsa-miR-3912-5p_syn | 0.62 |
| | hsa-miR-5684_syn | 0.611 |
| | hsa-miR-1281_syn | 0.591 |
| | hsa-miR-219a-1-3p_syn | 0.636 |
| | hsa-miR-3059-3p_syn | 0.589 |
| | hsa-miR-3616-5p_syn | 0.616 |
| | hsa-miR-1343-5p_syn | 0.634 |
| | hsa-miR-4726-5p_syn | 0.605 |
| | hsa-miR-4732-5p_syn | -0.87 |
| | hsa-miR-202-3p_syn | 0.613 |
| | hsa-miR-891a-5p_syn | 0.592 |
| | hsa-miR-548c-3p_syn | 0.598 |
| | hsa-miR-149-5p_syn | 0.614 |
| | hsa-miR-1250-3p_syn | 0.736 |
| | hsa-miR-1912-5p_syn | 0.595 |
| | hsa-miR-4716-5p_syn | 0.587 |
| | hsa-miR-708-5p_syn | 0.587 |
| | hsa-miR-144-5p_syn | -0.943 |
| | hsa-miR-10392-3p_syn | 0.59 |
| | hsa-miR-4748_syn | 0.587 |
| | hsa-miR-766-3p_syn | -0.654 |
| | hsa-miR-548aq-5p_syn | 0.608 |
| | hsa-miR-1255b-5p_syn | 0.593 |
| | hsa-miR-1226-5p_syn | 0.607 |
| | hsa-miR-6741-3p_syn | 0.592 |
| | hsa-miR-3617-5p_syn | 0.594 |
| | hsa-miR-6130_syn | 0.6 |
| | hsa-miR-765_syn | 0.588 |
| | hsa-miR-1825_syn | -0.814 |
| | hsa-miR-4732-3p_syn | -0.615 |
| | hsa-miR-486-3p_syn | -0.74 |
| | hsa-miR-183-5p_syn | -0.708 |
| | hsa-miR-486-5p_syn | -0.756 |
| | hsa-miR-2053_syn | 0.718 |

| | | |
|---|---|---|
| **miRNA urine** | hsa-miR-577_uri | 0.832 |
| | hsa-miR-301a-3p_uri | 0.93 |
| | hsa-miR-653-3p_uri | 0.944 |
| | hsa-miR-411-5p_uri | 0.946 |
| | hsa-miR-489-3p_uri | 0.923 |
| | hsa-miR-100-5p_uri | 0.781 |
| | hsa-miR-29a-3p_uri | 1.316 |
| | hsa-miR-30a-3p_uri | 1.308 |
| | hsa-miR-95-3p_uri | 0.887 |
| | hsa-miR-654-3p_uri | 0.823 |
| | hsa-miR-542-3p_uri | 0.883 |
| | hsa-miR-122-5p_uri | 0.853 |
| | hsa-miR-514a-3p_uri | 0.794 |
| | hsa-miR-127-3p_uri | 0.8 |
| | hsa-miR-136-5p_uri | 0.716 |
| | hsa-miR-451a_uri | 1.154 |
| | hsa-miR-96-5p_uri | 0.769 |
| | hsa-miR-454-3p_uri | 0.705 |
| | hsa-miR-135b-5p_uri | 0.626 |
| | hsa-miR-144-3p_uri | 0.863 |
| | hsa-miR-122b-3p_uri | 0.759 |
| | hsa-miR-204-3p_uri | 0.755 |
| | hsa-miR-598-3p_uri | 0.604 |
| | hsa-miR-11181-3p_uri | -1.486 |
| | hsa-miR-340-5p_uri | 0.755 |
| | hsa-miR-494-3p_uri | 0.609 |
| | hsa-miR-379-5p_uri | 0.615 |
| | hsa-miR-2052_uri | -1.147 |
| | hsa-miR-320a-5p_uri | -0.942 |
| | hsa-miR-381-3p_uri | 0.599 |
| | hsa-miR-1269a_uri | 0.635 |
| | hsa-miR-150-3p_uri | -0.673 |
| | hsa-miR-20b-3p_uri | -0.976 |
| | hsa-miR-661_uri | -0.932 |
| | hsa-miR-183-5p_uri | 0.672 |
| | hsa-miR-193a-3p_uri | -1.139 |
| | hsa-miR-222-5p_uri | -0.65 |
| | hsa-miR-9851-3p_uri | -0.825 |
| | hsa-miR-7515_uri | -0.801 |
| | hsa-miR-1281_uri | -0.831 |
| | hsa-miR-148a-3p_uri | 0.902 |
| | hsa-miR-920_uri | -0.743 |
| | hsa-miR-152-5p_uri | -0.743 |
| | hsa-miR-196a-1-3p_uri | -0.594 |
| | hsa-miR-224-5p_uri | 0.755 |
| | hsa-miR-19a-5p_uri | -0.601 |
| | hsa-miR-3074-3p_uri | -0.671 |
| | hsa-let-7g-5p_uri | 0.656 |
| **Cluster 2 vs 1 and 3** | | |
| **Multiomic data domain** | **Features** | **logFold Change** |
| **Metabolites plasma** | Alanine_pla | -0.19602393 |
| | Citruline_pla | 0.192801609 |
| | Ornithine_pla | 0.148444912 |
| | lysoPC.a.C20.4_pla | -0.138867491 |
| | Proline_pla | -0.165024088 |
| | Aspartic.acid_pla | -0.141704097 |
| | Acetylornithoine_pla | -0.161160047 |
| **miRNA plasma** | hsa-miR-379-5p_pla | 0.805901574 |
| | hsa-miR-3942-5p_pla | 0.886157981 |
| | hsa-miR-431-5p_pla | 0.795891185 |
| | hsa-miR-1299_pla | 1.033803959 |
| | hsa-miR-33b-5p_pla | 0.59262222 |
| | hsa-miR-4669_pla | 0.605706599 |
| | hsa-miR-181c-3p_pla | 0.614393663 |
| | hsa-miR-370-3p_pla | 0.619005368 |
| **miRNA synovial fluid** | hsa-miR-185-5p_syn | -0.647704065 |
| | hsa-miR-19b-3p_syn | -0.617703 |
| | hsa-miR-194-5p_syn | -0.830331527 |
| | hsa-miR-1-5p_syn | -0.588838911 |
| | hsa-miR-7106-3p_syn | -0.738287213 |

| | | |
|---|---|---|
| | hsa-miR-363-3p_syn | -0.847804634 |
| | hsa-miR-20b-5p_syn | -0.977391629 |
| | hsa-miR-3622b-3p_syn | 0.877940029 |
| | hsa-miR-19a-3p_syn | -0.611264334 |
| | hsa-miR-362-5p_syn | 0.860053341 |
| | hsa-miR-145-3p_syn | -1.1751026 |
| | hsa-miR-484_syn | -0.614649798 |
| | hsa-miR-323a-3p_syn | 0.737620678 |
| | hsa-miR-374b-3p_syn | 0.796331618 |
| | hsa-miR-767-5p_syn | 0.739136807 |
| | hsa-miR-34b-3p_syn | 0.803327676 |
| | hsa-miR-642a-5p_syn | 0.795823218 |
| | hsa-miR-451a_syn | -0.99694219 |
| | hsa-miR-26a-1-3p_syn | 0.700934986 |
| | hsa-miR-503-5p_syn | -0.640687414 |
| | hsa-miR-376a-3p_syn | 0.685050791 |
| | hsa-miR-192-5p_syn | -0.685586546 |
| | hsa-miR-4662b_syn | 0.595177023 |
| | hsa-miR-184_syn | 1.236817591 |
| | hsa-miR-505-3p_syn | -0.601493048 |
| | hsa-miR-133b_syn | -1.109406436 |
| | hsa-miR-1238-3p_syn | 0.724252871 |
| | hsa-miR-758-5p_syn | 0.649030084 |
| | hsa-miR-1226-5p_syn | 0.690704978 |
| | hsa-miR-642b-5p_syn | 0.651752555 |
| | hsa-miR-3144-3p_syn | -0.62489778 |
| | hsa-miR-6843-3p_syn | 0.621721868 |
| | hsa-miR-4773_syn | 0.679421214 |
| | hsa-miR-100-3p_syn | -1.021075018 |
| | hsa-miR-6883-3p_syn | 0.606539074 |
| | hsa-miR-3059-5p_syn | 0.660044252 |
| | hsa-miR-6855-3p_syn | 0.585585126 |
| | hsa-miR-1257_syn | -0.754262005 |
| | hsa-miR-766-5p_syn | -0.611348235 |
| | hsa-miR-329-3p_syn | 0.593284343 |
| | hsa-miR-642b-3p_syn | 0.669286972 |
| | hsa-miR-642a-3p_syn | 0.640719567 |
| | hsa-miR-96-5p_syn | -0.825662518 |
| | hsa-miR-1265_syn | 0.59716453 |
| | hsa-miR-16.2-3p_syn | -0.658010277 |
| | hsa-miR-493-3p_syn | 0.609129518 |
| | hsa-miR-144-5p_syn | -0.88608149 |
| | hsa-miR-4772-3p_syn | -0.627811396 |
| | hsa-miR-4732-3p_syn | -0.651323877 |
| | hsa-miR-486-3p_syn | -0.70568388 |
| | hsa-miR-206_syn | -0.957778105 |
| | hsa-miR-486-5p_syn | -0.676589218 |
| | hsa-miR-183-5p_syn | -0.602459444 |
| | hsa-miR-2053_syn | 0.586661134 |
| **miRNA urine** | hsa-miR-203b-5p_uri | 0.78355127 |
| | hsa-miR-203a-3p_uri | 0.757154442 |
| | hsa-miR-221-5p_uri | 0.857362746 |
| | hsa-miR-30d-3p_uri | -0.678335667 |
| | hsa-miR-944_uri | 0.785980944 |
| | hsa-miR-196a-3p_uri | 0.629577481 |
| | hsa-miR-891a-5p_uri | -0.813206853 |
| | hsa-miR-30a-3p_uri | -0.670010603 |
| | hsa-miR-150-5p_uri | 0.681518848 |
| | hsa-miR-143-3p_uri | 0.848808729 |
| | hsa-miR-615-3p_uri | -0.624304341 |
| | hsa-miR-142-5p_uri | 0.872630233 |
| | hsa-miR-223-5p_uri | 0.978134807 |
| | hsa-miR-145-5p_uri | 0.610436672 |
| | hsa-miR-193a-3p_uri | 0.68722247 |
| | hsa-miR-11181-3p_uri | 0.752669874 |
| | hsa-miR-223-3p_uri | 0.763571932 |
| | hsa-miR-2052_uri | 0.588831819 |
| **Cluster 3 vs 1 and 2** | | |
| **Multiomic data domain** | **Features** | **logFold Change** |
| **Metabolites plasma** | Acetylornithoine_pla | -0.543807467 |

| | | |
|---|---|---|
| | Kynurenine_pla | 0.187924966 |
| | Serine_pla | -0.174376755 |
| | Citruline_pla | 0.220034241 |
| | Aspartic.acid_pla | 0.154171025 |
| **miRNA plasma** | hsa-miR-196a-5p_pla | 0.687390149 |
| | hsa-miR-5010-3p_pla | -0.626710563 |
| | hsa-miR-145-3p_pla | -0.653096356 |
| | hsa-miR-6842-3p_pla | -0.730318495 |
| | hsa-miR-889-3p_pla | -0.739780333 |
| | hsa-miR-3942-5p_pla | 0.645039443 |
| | hsa-miR-570-3p_pla | -0.596234956 |
| | hsa-miR-642a-3p_pla | 0.618706237 |
| **miRNA synovial fluid** | hsa-miR-6500-3p_syn | -0.931828525 |
| | hsa-miR-618_syn | -1.001735537 |
| | hsa-miR-1-3p_syn | -0.655301308 |
| | hsa-miR-642b-5p_syn | 0.797607901 |
| | hsa-miR-151a-3p_syn | 0.709428234 |
| | hsa-miR-4685-3p_syn | -0.785122123 |
| | hsa-miR-1301-5p_syn | -1.087080782 |
| | hsa-miR-642a-3p_syn | 0.722229652 |
| | hsa-miR-1265_syn | -0.649982857 |
| | hsa-miR-1287-3p_syn | -0.591893221 |
| | hsa-miR-3059-3p_syn | -0.696505321 |
| | hsa-miR-1249-3p_syn | -0.819423588 |
| | hsa-miR-6862-5p_syn | -0.670604908 |
| | hsa-miR-106b-5p_syn | -0.622459298 |
| | hsa-miR-335-3p_syn | -0.790416622 |
| | hsa-miR-3188_syn | -0.658659465 |
| | hsa-miR-3909_syn | -0.601883189 |
| | hsa-miR-539-3p_syn | -0.638961605 |
| | hsa-miR-376a-2-5p_syn | -0.631040942 |
| | hsa-miR-4742-3p_syn | -0.60347597 |
| | hsa-miR-500b-3p_syn | -0.627205486 |
| | hsa-miR-181c-5p_syn | 0.58994652 |
| | hsa-miR-499b-3p_syn | -0.616214301 |
| | hsa-miR-379-3p_syn | -0.604064155 |
| | hsa-miR-133b_syn | 0.792782058 |
| | hsa-miR-100-3p_syn | 0.606221526 |
| | hsa-miR-206_syn | -0.615037123 |
| **miRNA urine** | hsa-miR-142-3p_uri | 1.010874437 |
| | hsa-miR-376b-3p_uri | -0.58954565 |
| | hsa-miR-203a-3p_uri | 0.679382798 |
| | hsa-miR-6734-5p_uri | -0.614429853 |
| | hsa-miR-203b-5p_uri | 0.661793679 |
| | hsa-miR-223-3p_uri | 0.926336398 |
| | hsa-miR-1299_uri | -0.692232451 |
| | hsa-miR-122-5p_uri | -0.612502615 |
| | hsa-miR-142-5p_uri | 0.689438138 |

**Supplementary Table 11. Cluster-wise Variable Selection Criteria for Unimodal Machine Learning Models classifying WOMAC function response.** Variables within multiomic data domains were selected based on criteria: absolute(log fold change) > 1.1 for plasma metabolites and > 1.5 for miRNAs. These criteria ensured the inclusion of biologically significant features for further analysis. Subsequently, selected variables were integrated into the integrative machine learning framework to classify WOMAC function response within each cluster.

| Cluster 1 vs 2 and 3 | | |
|---|---|---|
| **Multiomic data domain** | **Features** | **logFold Change** |
| Metabolites plasma | PC.aa.C38.4_pla | -0.2448322 |
| | PC.aa.C38.0_pla | -0.1640032 |
| | lysoPC.a.C20.4_pla | -0.195903 |
| | PC.aa.C40.6_pla | -0.175797 |
| | PC.aa.C36.0_pla | -0.1487664 |
| | Tyrosine_pla | 0.15726127 |
| | PC.aa.C38.6_pla | -0.142914 |
| | trans.hydroxyproline_pla | -0.2280933 |
| | PC.aa.C34.2_pla | -0.1389794 |
| | PC.aa.C28.1_pla | 0.17180618 |
| miRNA plasma | hsa-miR-1255a_pla | -0.7779667 |
| | hsa-miR-3173-5p_pla | -0.6327021 |
| | hsa-miR-497-5p_pla | -0.6133439 |
| | hsa-miR-320d_pla_pla | -0.6101234 |
| | hsa-miR-374a-3p_pla | -0.6137109 |
| miRNA synovial fluid | hsa-let-7f-1-3p_syn | -0.7710164 |
| | hsa-miR-766-3p_syn | -0.77498 |
| | hsa-miR-921_syn | -0.6520734 |
| | hsa-miR-153-5p_syn | -0.6510752 |
| | hsa-miR-500a-5p_syn | -0.5972108 |
| | hsa-miR-1227-5p_syn | -0.6913695 |
| | hsa-miR-26b-5p_syn | -0.5907307 |
| | hsa-miR-323b-5p_syn | 0.67846597 |
| | hsa-miR-3609_syn | -0.6718589 |
| | hsa-miR-429_syn | -0.5958588 |
| | hsa-miR-2053_syn | 0.81961278 |
| | hsa-miR-144-5p_syn | -0.6762372 |
| | hsa-miR-485-3p_syn | 0.61568296 |
| | hsa-miR-206_syn | 0.66806986 |
| miRNA urine | hsa-miR-155-5p_uri | -0.700368 |
| | hsa-miR-577_uri | 0.65666098 |
| | hsa-miR-142-3p_uri | -1.0615248 |
| | hsa-miR-455-3p_uri | 0.64311925 |
| | hsa-miR-654-3p_uri | 0.97122973 |
| | hsa-miR-122-5p_uri | 1.0042383 |
| | hsa-miR-379-5p_uri | 0.85751469 |
| | hsa-miR-10a-3p_uri | 0.7753546 |
| | hsa-miR-222-3p_uri | 0.86905109 |
| | hsa-miR-3184-3p_uri | 0.6765934 |
| | hsa-miR-136-5p_uri | 0.79327408 |
| | hsa-miR-125b-1-3p_uri | 0.69640183 |
| | hsa-miR-449c-5p_uri | 0.64601143 |
| | hsa-miR-375-3p_uri | -0.6837023 |
| | hsa-miR-100-3p_uri | 0.64770301 |
| | hsa-miR-377-3p_uri | 0.73420827 |
| | hsa-miR-219a-5p_uri | 0.5873169 |
| | hsa-miR-4755-3p_uri | 0.741147 |
| | hsa-miR-625-3p_uri | 0.80792905 |
| | hsa-miR-487b-3p_uri | 0.68395882 |
| | hsa-miR-127-3p_uri | 0.72940688 |
| | hsa-miR-494-3p_uri | 0.66168647 |
| | hsa-miR-411-5p_uri | 0.71966711 |
| | hsa-miR-1271-5p_uri | 0.64012458 |
| | hsa-miR-653-3p_uri | 0.66544816 |
| | hsa-miR-664a-3p_uri | 0.61926672 |
| | hsa-miR-142-5p_uri | -0.8907906 |
| | hsa-miR-432-5p_uri | 0.65443453 |
| | hsa-miR-204-3p_uri | 0.65667564 |
| | hsa-miR-489-3p_uri | 0.60482567 |
| | hsa-miR-514a-3p_uri | 0.59532036 |

| | | |
|---|---|---|
| | hsa-miR-409-3p_uri | 0.59112903 |
| | hsa-miR-223-3p_uri | -0.8004769 |
| | hsa-miR-191-5p_uri | -0.6690025 |
| | hsa-miR-30a-3p_uri | 0.69180818 |
| **Cluster 2 vs 1 and 3** | | |
| **Multiomic data domain** | **Features** | **logFold Change** |
| **Metabolites plasma** | SM.OH.C22.2_pla | -0.1945517 |
| | Glutamine_pla | -0.284101 |
| | PC.ae.C36.3_pla | -0.2345231 |
| | Aspartic.acid_pla | -0.2513432 |
| | Serine_pla | -0.153093 |
| **miRNA plasma** | hsa-miR-3942-5p_pla | 1.07769226 |
| | hsa-miR-1976_pla | -0.6799886 |
| | hsa-miR-379-5p_pla | 0.71521287 |
| | hsa-miR-30b-5p_pla | 0.60956471 |
| | hsa-miR-4685-3p_pla | 0.6133091 |
| | hsa-miR-1260b_pla | -0.6690885 |
| | hsa-miR-676-3p_pla | 0.63994308 |
| **miRNA synovial fluid** | hsa-miR-941_syn | 0.91330523 |
| | hsa-miR-378c_syn | 0.79392715 |
| | hsa-miR-1265_syn | 0.75987122 |
| | hsa-miR-6516-5p_syn | 0.64035949 |
| | hsa-miR-642a-5p_syn | 0.79515866 |
| | hsa-miR-33a-3p_syn | 0.64468682 |
| | hsa-miR-3928-3p_syn | 0.68665691 |
| | hsa-miR-3605-3p_syn | -0.6794698 |
| | hsa-miR-101-3p_syn | 0.64754275 |
| | hsa-miR-1307-3p_syn | 0.61992754 |
| | hsa-miR-642b-3p_syn | 0.7254036 |
| | hsa-miR-660-3p_syn | 0.59704036 |
| | hsa-miR-548aw_syn | 0.60073948 |
| | hsa-miR-1251-5p_syn | 0.61993007 |
| | hsa-miR-1537-5p_syn | 0.60713288 |
| | hsa-miR-548aq-3p_syn | 0.61829381 |
| | hsa-miR-758-5p_syn | 0.61900261 |
| | hsa-miR-495-3p_syn | 0.64796257 |
| | hsa-miR-20b-5p_syn | -0.7302366 |
| | hsa-miR-496_syn | 0.62806053 |
| | hsa-miR-4454_syn | 0.63190015 |
| | hsa-miR-362-5p_syn | 0.60575784 |
| | hsa-miR-4732-3p_syn | -0.794658 |
| | hsa-miR-34b-3p_syn | 0.61564846 |
| | hsa-miR-100-3p_syn | -0.9292159 |
| | hsa-miR-184_syn | 0.98616005 |
| | hsa-miR-206_syn | -1.230743 |
| | hsa-miR-145-3p_syn | -0.7278827 |
| | hsa-miR-144-5p_syn | -0.5903612 |
| **miRNA urine** | hsa-miR-584-5p_uri | 0.65465799 |
| | hsa-miR-26a-1-3p_uri | 0.60122339 |
| | hsa-miR-22-3p_uri | 0.5910385 |
| | hsa-miR-146b-3p_uri | 0.58577675 |
| | hsa-miR-192-5p_uri | 0.63060901 |
| | hsa-miR-223-5p_uri | 0.66472428 |
| **Cluster 3 vs 1 and 2** | | |
| **Multiomic data domain** | **Features** | **logFold Change** |
| **Metabolites plasma** | Glutamine_pla | -0.373509 |
| | Ornithine_pla | -0.1662461 |
| | Arginine_pla | 0.18595027 |
| | PC.ae.C32.1_pla | 0.13954326 |
| | Asparagine_pla | 0.14424786 |
| **miRNA plasma** | hsa-miR-196a-5p_pla | 0.89477769 |
| | hsa-miR-642a-3p_pla | 0.99400797 |
| | hsa-miR-6734-5p_pla | 0.72627786 |
| | hsa-miR-126-3p_pla | -0.6883903 |
| | hsa-miR-181d-5p_pla | -0.725938 |
| | hsa-miR-96-5p_pla | -0.6231056 |
| | hsa-miR-411-5p_pla | -0.7134185 |
| | hsa-miR-122-3p_pla | 0.58983703 |
| **miRNA synovial fluid** | hsa-miR-6500-3p_syn | -0.8657275 |

| | | |
|---|---|---|
| | hsa-miR-6510-5p_syn | -0.7671805 |
| | hsa-miR-1287-3p_syn | -0.673971 |
| | hsa-miR-6511b-3p_syn | -0.7141641 |
| | hsa-miR-579-3p_syn | -0.7009682 |
| | hsa-miR-335-3p_syn | -0.7827945 |
| | hsa-miR-3913-5p_syn | -0.9817368 |
| | hsa-miR-6780a-5p_syn | -0.6704367 |
| | hsa-miR-326_syn | -0.7741961 |
| | hsa-miR-106b-5p_syn | -0.7319185 |
| | hsa-miR-20b-5p_syn | -0.5969407 |
| | hsa-miR-181b-2-3p_syn | -0.6180446 |
| | hsa-miR-3684_syn | -0.6412287 |
| | hsa-miR-2114-3p_syn | -0.7501865 |
| | hsa-miR-4520-3p_syn | -0.6122823 |
| | hsa-miR-4685-3p_syn | -0.6674781 |
| | hsa-miR-885-5p_syn | 0.67285922 |
| | hsa-miR-500b-3p_syn | -0.6116633 |
| | hsa-miR-30b-3p_syn | -0.6035989 |
| | hsa-miR-891a-5p_syn | -0.5969773 |
| | hsa-miR-548d-3p_syn | -0.5899951 |
| | hsa-miR-412-5p_syn | 0.75091044 |
| **miRNA urine** | hsa-miR-501-5p_uri | -0.7387993 |
| | hsa-miR-142-3p_uri | 1.07527434 |
| | hsa-miR-4763-3p_uri | -0.6329543 |
| | hsa-miR-378c_uri | -0.778961 |
| | hsa-miR-223-3p_uri | 0.98452642 |
| | hsa-miR-1269a_uri | 0.7014579 |
| | hsa-miR-27a-5p_uri | 0.63128547 |
| | hsa-miR-205-5p_uri | 0.63785698 |
| | hsa-miR-607_uri | 0.64198177 |
| | hsa-miR-4662b_uri | 0.63001072 |
| | hsa-miR-148a-5p_uri | 0.68563968 |
| | hsa-miR-143-3p_uri | 0.63125843 |

**Supplementary Table 12.** Cluster-wise mean Gini impurity-based feature importance across various domains, including clinical data, miRNA plasma, miRNA synovial, miRNA urine, and metabolite plasma, within the integrated model classifying WOMAC pain response.

| **Cluster 1 vs 2 and 3** | | |
|---|---|---|
| **Data domain** | **Features** | **Importance Score** |
| **Clinical** | Age | 1.48987345 |
| | Sex | 1.99977745 |
| | BMI | 1.23073645 |
| | HADS anxiety category | 1.69382688 |
| | HADS depression category | 2.1850492 |
| | painDetect category | 0.90910171 |
| **Metabolites plasma** | alpha.Aminoadipicacid_pla | 1.16930778 |
| | trans.hydroxyproline_pla | 2.14407428 |
| | Citruline_pla | 1.36457426 |
| **miRNA plasma** | hsa-miR-4429_pla | 3.44339765 |
| | hsa-miR-208b-3p_pla | 1.25664547 |
| **miRNA synovial fluid** | hsa-miR-5698_syn | 1.41235052 |
| | hsa-miR-6758-3p_syn | 2.12527254 |
| | hsa-miR-1273h-5p_syn | 1.01292251 |
| | hsa-let-7f-1-3p_syn | 2.33744984 |
| | hsa-miR-2116-5p_syn | 2.10925454 |
| | hsa-miR-1251-5p_syn | 2.72226874 |
| | hsa-miR-6799-5p_syn | 1.78687897 |
| | hsa-miR-4688_syn | 2.02394642 |
| | hsa-miR-654-5p_syn | 1.11508053 |
| | hsa-miR-3150a-5p_syn | 1.36142274 |
| | hsa-miR-6837-3p_syn | 3.66528452 |
| | hsa-miR-548ax_syn | 0.92966102 |
| | hsa-miR-181d-3p_syn | 2.13177196 |
| | hsa-miR-873-5p_syn | 2.02361305 |
| | hsa-miR-619-5p_syn | 1.32502958 |
| | hsa-miR-3918_syn | 2.3442401 |
| | hsa-miR-1227-5p_syn | 0.97798606 |
| | hsa-miR-6729-5p_syn | 0.99102177 |
| | hsa-miR-4717-3p_syn | 1.68061809 |
| | hsa-miR-378d_syn | 1.67733614 |
| | hsa-miR-4670-5p_syn | 1.8394048 |
| | hsa-miR-4724-5p_syn | 2.1871171 |
| | hsa-miR-508-3p_syn | 2.1178174 |
| | hsa-miR-6820-3p_syn | 1.08663402 |
| | hsa-miR-12116_syn | 0.91915328 |
| | hsa-miR-4687-5p_syn | 0.92299535 |
| | hsa-miR-6871-5p_syn | 1.37076804 |
| | hsa-miR-3164_syn | 1.01791204 |
| | hsa-miR-4659b-5p_syn | 2.01873283 |
| | hsa-miR-129-5p_syn | 1.70789258 |
| | hsa-miR-4785_syn | 1.19519958 |
| | hsa-miR-4649-5p_syn | 1.36541298 |
| | hsa-miR-4689_syn | 2.0824401 |
| | hsa-miR-6730-3p_syn | 1.3177068 |
| | hsa-miR-548b-5p_syn | 1.05461162 |
| | hsa-miR-4776-3p_syn | 1.3982677 |
| | hsa-miR-4516_syn | 1.93774718 |
| | hsa-miR-6721-5p_syn | 1.61578532 |
| | hsa-miR-6762-3p_syn | 1.06628282 |
| | hsa-miR-93-5p_syn | 0.98980233 |
| | hsa-miR-145-3p_syn | 1.24333901 |
| | hsa-miR-3201_syn | 3.84534247 |
| | hsa-miR-711_syn | 1.46007961 |
| | hsa-miR-5571-3p_syn | 1.24986655 |
| | hsa-miR-378i_syn | 2.58532981 |
| | hsa-miR-6511a-3p_syn | 1.85727727 |
| | hsa-miR-1301-3p_syn | 2.01462466 |
| | hsa-miR-4429_syn | 3.44339765 |
| | hsa-miR-4664-3p_syn | 1.29028457 |
| | hsa-miR-122b-3p_syn | 1.54068165 |
| | hsa-miR-1306-5p_syn | 1.99704755 |
| | hsa-miR-6818-3p_syn | 1.04734318 |

| | | |
|---|---|---|
| | hsa-miR-196b-3p_syn | 1.94521523 |
| | hsa-miR-6843-3p_syn | 1.62915208 |
| | hsa-miR-5704_syn | 1.66955064 |
| | hsa-miR-3529-5p_syn | 1.95655597 |
| | hsa-miR-1236-5p_syn | 1.4017467 |
| | hsa-miR-6769b-3p_syn | 1.86727476 |
| | hsa-miR-106a-3p_syn | 1.89260265 |
| | hsa-miR-4715-3p_syn | 0.96793624 |
| | hsa-miR-3158-3p_syn | 3.95970722 |
| | hsa-miR-3174_syn | 0.93619069 |
| | hsa-miR-548u_syn | 1.4389754 |
| | hsa-miR-4703-3p_syn | 1.83080421 |
| | hsa-miR-488-3p_syn | 1.79213861 |
| | hsa-miR-6810-3p_syn | 2.18435551 |
| | hsa-miR-3085-5p_syn | 1.70286222 |
| | hsa-miR-557_syn | 1.09172883 |
| | hsa-miR-4714-3p_syn | 1.4549025 |
| | hsa-miR-10525-3p_syn | 1.32551937 |
| | hsa-miR-187-5p_syn | 1.4424236 |
| | hsa-miR-10395-3p_syn | 1.92130433 |
| | hsa-miR-3126-5p_syn | 1.06332036 |
| | hsa-miR-3667-3p_syn | 1.48690748 |
| | hsa-miR-6509-5p_syn | 1.72322208 |
| | hsa-miR-19b-1-5p_syn | 1.32334165 |
| | hsa-miR-6748-5p_syn | 2.17584523 |
| | hsa-miR-4716-3p_syn | 1.91846594 |
| | hsa-miR-4680-3p_syn | 1.91808059 |
| | hsa-miR-548w_syn | 1.90758569 |
| | hsa-miR-5191_syn | 1.1473301 |
| | hsa-miR-4286_syn | 2.02183812 |
| | hsa-miR-938_syn | 1.18401288 |
| | hsa-miR-30b-3p_syn | 2.12166915 |
| | hsa-miR-615-5p_syn | 1.44209326 |
| | hsa-miR-6785-5p_syn | 2.01331447 |
| | hsa-miR-4449_syn | 1.04745897 |
| | hsa-miR-4687-3p_syn | 1.28096165 |
| | hsa-miR-4677-5p_syn | 1.70887255 |
| | hsa-miR-4511_syn | 1.42796662 |
| | hsa-miR-6763-3p_syn | 1.75125953 |
| | hsa-miR-3150b-3p_syn | 1.74699897 |
| | hsa-miR-451a_syn | 1.85746446 |
| | hsa-miR-148a-3p_syn | 3.1108887 |
| | hsa-miR-6873-3p_syn | 1.66662804 |
| | hsa-miR-5091_syn | 1.59759952 |
| | hsa-miR-4709-3p_syn | 2.04634843 |
| | hsa-miR-511-3p_syn | 2.01358593 |
| | hsa-miR-5193_syn | 2.1470222 |
| | hsa-miR-6826-3p_syn | 1.82616906 |
| | hsa-miR-3939_syn | 1.86944147 |
| | hsa-miR-6740-5p_syn | 1.94880334 |
| | hsa-miR-2117_syn | 1.29922113 |
| | hsa-miR-6766-3p_syn | 1.13678383 |
| | hsa-miR-7113-3p_syn | 1.41265489 |
| | hsa-miR-2682-5p_syn | 1.55031592 |
| | hsa-miR-1199-5p_syn | 1.58384186 |
| | hsa-miR-877-3p_syn | 0.92116612 |
| | hsa-miR-6810-5p_syn | 1.90091232 |
| | hsa-miR-3613-5p_syn | 2.005014 |
| | hsa-miR-6826-5p_syn | 1.37592194 |
| | hsa-miR-3192-5p_syn | 1.49038677 |
| | hsa-miR-6802-5p_syn | 1.68206739 |
| | hsa-miR-3616-3p_syn | 2.06686007 |
| | hsa-miR-7702_syn | 1.42874526 |
| | hsa-miR-5699-3p_syn | 1.85282414 |
| | hsa-miR-6787-3p_syn | 1.36016803 |
| | hsa-miR-7847-3p_syn | 1.68642361 |
| | hsa-miR-539-5p_syn | 1.01281763 |
| | hsa-miR-6732-3p_syn | 1.30785322 |
| | hsa-miR-6800-3p_syn | 1.33610958 |
| | hsa-miR-3139_syn | 1.18385028 |
| | hsa-miR-100-5p_syn | 1.38966598 |

| | | |
|---|---|---|
| | hsa-miR-345-3p_syn | 1.18299146 |
| | hsa-miR-4715-5p_syn | 1.00925287 |
| | hsa-miR-4727-3p_syn | 0.90266253 |
| | hsa-miR-6796-5p_syn | 0.94293495 |
| | hsa-miR-4742-5p_syn | 1.09928618 |
| | hsa-miR-6767-5p_syn | 0.95011971 |
| | hsa-miR-6754-5p_syn | 2.04829748 |
| | hsa-miR-6755-3p_syn | 1.39092941 |
| | hsa-miR-2114-3p_syn | 1.1859099 |
| | hsa-miR-6812-3p_syn | 1.47066391 |
| | hsa-miR-887-5p_syn | 1.33981756 |
| | hsa-miR-2116-3p_syn | 1.16171375 |
| | hsa-miR-582-5p_syn | 2.58050045 |
| | hsa-miR-6842-5p_syn | 1.38565442 |
| | hsa-miR-6866-3p_syn | 2.10855811 |
| | hsa-miR-3667-5p_syn | 1.31756054 |
| | hsa-miR-4467_syn | 1.29354298 |
| | hsa-miR-4670-3p_syn | 0.96303717 |
| | hsa-miR-4645-3p_syn | 0.90166738 |
| | hsa-miR-4714-5p_syn | 2.03527638 |
| | hsa-miR-3911_syn | 2.11721322 |
| | hsa-miR-4632-5p_syn | 1.42088869 |
| | hsa-miR-2681-3p_syn | 1.01837804 |
| | hsa-miR-4489_syn | 1.44291559 |
| | hsa-miR-6882-3p_syn | 1.01243304 |
| | hsa-miR-19a-5p_syn | 1.42574609 |
| | hsa-miR-3181_syn | 1.58197614 |
| | hsa-miR-4526_syn | 0.97903976 |
| | hsa-miR-4757-5p_syn | 0.93108083 |
| | hsa-miR-6753-3p_syn | 1.563479 |
| | hsa-miR-6883-3p_syn | 1.22429542 |
| | hsa-miR-1305_syn | 2.11258906 |
| | hsa-miR-6876-5p_syn | 1.2241172 |
| | hsa-miR-4802-5p_syn | 2.06831175 |
| | hsa-miR-7156-3p_syn | 2.00698245 |
| | hsa-miR-6771-5p_syn | 2.00987244 |
| | hsa-miR-4669_syn | 1.65136152 |
| | hsa-miR-618_syn | 1.23126807 |
| | hsa-miR-4762-5p_syn | 2.16754899 |
| | hsa-miR-548t-5p_syn | 1.23764453 |
| | hsa-miR-3912-5p_syn | 0.99922288 |
| | hsa-miR-5684_syn | 1.82432254 |
| | hsa-miR-1281_syn | 1.60243784 |
| | hsa-miR-219a-1-3p_syn | 1.96402883 |
| | hsa-miR-3059-3p_syn | 2.10926382 |
| | hsa-miR-3616-5p_syn | 1.11632729 |
| | hsa-miR-1343-5p_syn | 2.04521212 |
| | hsa-miR-4726-5p_syn | 0.96742677 |
| | hsa-miR-4732-5p_syn | 1.57943968 |
| | hsa-miR-202-3p_syn | 0.93668904 |
| | hsa-miR-891a-5p_syn | 1.25336699 |
| | hsa-miR-548c-3p_syn | 1.96481572 |
| | hsa-miR-149-5p_syn | 1.60095585 |
| | hsa-miR-1250-3p_syn | 1.00984504 |
| | hsa-miR-1912-5p_syn | 1.91782748 |
| | hsa-miR-4716-5p_syn | 1.06401144 |
| | hsa-miR-708-5p_syn | 1.29013015 |
| | hsa-miR-144-5p_syn | 3.23968487 |
| | hsa-miR-10392-3p_syn | 1.06464886 |
| | hsa-miR-4748_syn | 1.49581489 |
| | hsa-miR-766-3p_syn | 1.08442001 |
| | hsa-miR-548aq-5p_syn | 1.22731322 |
| | hsa-miR-1255b-5p_syn | 1.38145384 |
| | hsa-miR-1226-5p_syn | 1.82146719 |
| | hsa-miR-6741-3p_syn | 2.08302389 |
| | hsa-miR-3617-5p_syn | 1.80098891 |
| | hsa-miR-6130_syn | 0.99813265 |
| | hsa-miR-765_syn | 1.57210617 |
| | hsa-miR-1825_syn | 1.26189365 |
| | hsa-miR-4732-3p_syn | 1.45472406 |
| | hsa-miR-486-3p_syn | 1.43649239 |

| | | |
|---|---|---|
| | hsa-miR-183-5p_syn | 1.71706751 |
| | hsa-miR-486-5p_syn | 1.96504803 |
| | hsa-miR-2053_syn | 1.59357838 |
| **miRNA urine** | hsa-miR-577_uri | 3.25483418 |
| | hsa-miR-301a-3p_uri | 2.53154265 |
| | hsa-miR-653-3p_uri | 3.52122823 |
| | hsa-miR-411-5p_uri | 1.85578074 |
| | hsa-miR-489-3p_uri | 1.40239094 |
| | hsa-miR-100-5p_uri | 1.87630534 |
| | hsa-miR-29a-3p_uri | 1.51844786 |
| | hsa-miR-30a-3p_uri | 1.88814108 |
| | hsa-miR-95-3p_uri | 3.88721558 |
| | hsa-miR-654-3p_uri | 1.90016097 |
| | hsa-miR-542-3p_uri | 1.51894934 |
| | hsa-miR-122-5p_uri | 3.05213571 |
| | hsa-miR-514a-3p_uri | 1.63434733 |
| | hsa-miR-127-3p_uri | 1.63366851 |
| | hsa-miR-136-5p_uri | 1.54415068 |
| | hsa-miR-451a_uri | 1.81431275 |
| | hsa-miR-96-5p_uri | 1.72385829 |
| | hsa-miR-454-3p_uri | 0.96267917 |
| | hsa-miR-135b-5p_uri | 2.04843153 |
| | hsa-miR-144-3p_uri | 2.34585433 |
| | hsa-miR-122b-3p_uri | 1.065645 |
| | hsa-miR-204-3p_uri | 1.21454304 |
| | hsa-miR-598-3p_uri | 2.03040595 |
| | hsa-miR-11181-3p_uri | 1.06349767 |
| | hsa-miR-340-5p_uri | 1.6000759 |
| | hsa-miR-494-3p_uri | 0.93526593 |
| | hsa-miR-379-5p_uri | 1.58828582 |
| | hsa-miR-2052_uri | 1.95442601 |
| | hsa-miR-320a-5p_uri | 1.8636647 |
| | hsa-miR-381-3p_uri | 2.04989743 |
| | hsa-miR-1269a_uri | 3.98528942 |
| | hsa-miR-150-3p_uri | 1.44608291 |
| | hsa-miR-20b-3p_uri | 1.67618457 |
| | hsa-miR-661_uri | 1.22903405 |
| | hsa-miR-183-5p_uri | 0.92431892 |
| | hsa-miR-193a-3p_uri | 0.97724587 |
| | hsa-miR-222-5p_uri | 2.80076992 |
| | hsa-miR-9851-3p_uri | 1.66239664 |
| | hsa-miR-7515_uri | 0.90260172 |
| | hsa-miR-1281_uri | 0.94357462 |
| | hsa-miR-148a-3p_uri | 3.1108887 |
| | hsa-miR-920_uri | 1.2145694 |
| | hsa-miR-152-5p_uri | 1.58193931 |
| | hsa-miR-196a-1-3p_uri | 1.70397802 |
| | hsa-miR-224-5p_uri | 1.9858381 |
| | hsa-miR-19a-5p_uri | 2.17318657 |
| | hsa-miR-3074-3p_uri | 2.14856887 |
| | hsa-let-7g-5p_uri | 0.9083253 |
| **Cluster 2 vs 1 and 3** | | |
| **Multiomic data domain** | **Features** | **Importance Score** |
| **Clinical** | Age | 1.02577823 |
| | Sex | 1.50628245 |
| | BMI | 1.60944184 |
| | HADS anxiety category | 0.90427594 |
| | HADS depression category | 1.03708807 |
| | painDetect category | 1.71576553 |
| **Metabolites plasma** | Alanine_pla | 1.74411473 |
| | Citruline_pla | 3.58627933 |
| | Ornithine_pla | 3.4173309 |
| | lysoPC.a.C20.4_pla | 2.51825916 |
| | Proline_pla | 1.46557754 |
| | Aspartic.acid_pla | 1.67295574 |
| | Acetylornithoine_pla | 1.92448388 |
| **miRNA plasma** | hsa-miR-379-5p_pla | 1.30916937 |
| | hsa-miR-3942-5p_pla | 2.4519029 |
| | hsa-miR-431-5p_pla | 1.53577017 |
| | hsa-miR-1299_pla | 1.05373933 |

| | | |
|---|---|---|
| | hsa-miR-33b-5p_pla | 2.72336601 |
| | hsa-miR-4669_pla | 4.18447476 |
| | hsa-miR-181c-3p_pla | 2.15308887 |
| | hsa-miR-370-3p_pla | 1.60237364 |
| **miRNA synovial fluid** | hsa-miR-185-5p_syn | 1.54035753 |
| | hsa-miR-19b-3p_syn | 1.79902634 |
| | hsa-miR-194-5p_syn | 1.64068921 |
| | hsa-miR-1-5p_syn | 1.59574293 |
| | hsa-miR-7106-3p_syn | 0.99353447 |
| | hsa-miR-363-3p_syn | 1.96381146 |
| | hsa-miR-20b-5p_syn | 1.00167127 |
| | hsa-miR-3622b-3p_syn | 1.87332111 |
| | hsa-miR-19a-3p_syn | 1.09505603 |
| | hsa-miR-362-5p_syn | 1.75544895 |
| | hsa-miR-145-3p_syn | 2.04139441 |
| | hsa-miR-484_syn | 2.17537787 |
| | hsa-miR-323a-3p_syn | 3.71216747 |
| | hsa-miR-374b-3p_syn | 3.64281684 |
| | hsa-miR-767-5p_syn | 1.708175 |
| | hsa-miR-34b-3p_syn | 3.14786695 |
| | hsa-miR-642a-5p_syn | 1.33669527 |
| | hsa-miR-451a_syn | 1.37983783 |
| | hsa-miR-26a-1-3p_syn | 1.86689418 |
| | hsa-miR-503-5p_syn | 2.02481725 |
| | hsa-miR-376a-3p_syn | 1.75309618 |
| | hsa-miR-192-5p_syn | 1.00908794 |
| | hsa-miR-4662b_syn | 1.11328419 |
| | hsa-miR-184_syn | 0.93449848 |
| | hsa-miR-505-3p_syn | 1.95665807 |
| | hsa-miR-133b_syn | 3.02888449 |
| | hsa-miR-1238-3p_syn | 1.4945204 |
| | hsa-miR-758-5p_syn | 1.13469096 |
| | hsa-miR-1226-5p_syn | 1.00949149 |
| | hsa-miR-642b-5p_syn | 1.8251262 |
| | hsa-miR-3144-3p_syn | 1.9820548 |
| | hsa-miR-6843-3p_syn | 1.39877892 |
| | hsa-miR-4773_syn | 2.02708035 |
| | hsa-miR-100-3p_syn | 1.71978194 |
| | hsa-miR-6883-3p_syn | 1.33561186 |
| | hsa-miR-3059-5p_syn | 1.49442681 |
| | hsa-miR-6855-3p_syn | 1.17041836 |
| | hsa-miR-1257_syn | 1.09695472 |
| | hsa-miR-766-5p_syn | 2.97835967 |
| | hsa-miR-329-3p_syn | 1.53623772 |
| | hsa-miR-642b-3p_syn | 1.82844253 |
| | hsa-miR-642a-3p_syn | 3.54202939 |
| | hsa-miR-96-5p_syn | 1.2489188 |
| | hsa-miR-1265_syn | 3.66526972 |
| | hsa-miR-16.2-3p_syn | 2.19603546 |
| | hsa-miR-493-3p_syn | 0.91036312 |
| | hsa-miR-144-5p_syn | 1.19473136 |
| | hsa-miR-4772-3p_syn | 1.07559466 |
| | hsa-miR-4732-3p_syn | 1.76353145 |
| | hsa-miR-486-3p_syn | 1.90499555 |
| | hsa-miR-206_syn | 2.14348349 |
| | hsa-miR-486-5p_syn | 1.5302017 |
| | hsa-miR-183-5p_syn | 1.43391972 |
| | hsa-miR-2053_syn | 1.61983195 |
| **miRNA urine** | hsa-miR-203b-5p_uri | 4.04954717 |
| | hsa-miR-203a-3p_uri | 2.02812789 |
| | hsa-miR-221-5p_uri | 3.45782513 |
| | hsa-miR-30d-3p_uri | 3.46326886 |
| | hsa-miR-944_uri | 3.11285195 |
| | hsa-miR-196a-3p_uri | 3.68592077 |
| | hsa-miR-891a-5p_uri | 3.41763245 |
| | hsa-miR-30a-3p_uri | 1.07068694 |
| | hsa-miR-150-5p_uri | 1.97246596 |
| | hsa-miR-143-3p_uri | 1.4246088 |
| | hsa-miR-615-3p_uri | 2.83154592 |
| | hsa-miR-142-5p_uri | 1.27835135 |
| | hsa-miR-223-5p_uri | 1.47792975 |

| | | |
|---|---|---|
| | hsa-miR-145-5p_uri | 1.86665762 |
| | hsa-miR-193a-3p_uri | 1.99107746 |
| | hsa-miR-11181-3p_uri | 1.69491211 |
| | hsa-miR-223-3p_uri | 1.77795667 |
| | hsa-miR-2052_uri | 1.39246693 |
| **Cluster 3 vs 1 and 2** | | |
| **Multiomic data domain** | **Features** | **Importance Score** |
| **Clinical** | Age | 1.46070165 |
| | Sex | 2.05646753 |
| | BMI | 2.06423538 |
| | HADS anxiety category | 2.00341768 |
| | HADS depression category | 1.55827887 |
| | painDetect category | 1.15486055 |
| **Metabolites plasma** | Acetylornithoine_pla | 3.70698409 |
| | Kynurenine_pla | 1.80683539 |
| | Serine_pla | 3.04826528 |
| | Citruline_pla | 1.4485979 |
| | Aspartic.acid_pla | 1.74233819 |
| **miRNA plasma** | hsa-miR-196a-5p_pla | 2.67142012 |
| | hsa-miR-5010-3p_pla | 2.08711711 |
| | hsa-miR-145-3p_pla | 2.09390532 |
| | hsa-miR-6842-3p_pla | 1.00764471 |
| | hsa-miR-889-3p_pla | 4.29733659 |
| | hsa-miR-3942-5p_pla | 3.47743674 |
| | hsa-miR-570-3p_pla | 1.84958113 |
| | hsa-miR-642a-3p_pla | 3.13625216 |
| **miRNA synovial fluid** | hsa-miR-6500-3p_syn | 2.55159667 |
| | hsa-miR-618_syn | 2.90966897 |
| | hsa-miR-1-3p_syn | 2.18639729 |
| | hsa-miR-642b-5p_syn | 2.76700152 |
| | hsa-miR-151a-3p_syn | 1.70080419 |
| | hsa-miR-4685-3p_syn | 3.68165265 |
| | hsa-miR-1301-5p_syn | 3.47963915 |
| | hsa-miR-642a-3p_syn | 3.13625216 |
| | hsa-miR-1265_syn | 3.94755864 |
| | hsa-miR-1287-3p_syn | 1.23839647 |
| | hsa-miR-3059-3p_syn | 1.58641289 |
| | hsa-miR-1249-3p_syn | 4.10783813 |
| | hsa-miR-6862-5p_syn | 2.10094053 |
| | hsa-miR-106b-5p_syn | 1.70092679 |
| | hsa-miR-335-3p_syn | 1.46868454 |
| | hsa-miR-3188_syn | 0.97095987 |
| | hsa-miR-3909_syn | 2.12213594 |
| | hsa-miR-539-3p_syn | 2.55177155 |
| | hsa-miR-376a-2-5p_syn | 1.43811878 |
| | hsa-miR-4742-3p_syn | 1.58664148 |
| | hsa-miR-500b-3p_syn | 1.78313832 |
| | hsa-miR-181c-5p_syn | 2.43133476 |
| | hsa-miR-499b-3p_syn | 1.77803832 |
| | hsa-miR-379-3p_syn | 2.23889088 |
| | hsa-miR-133b_syn | 2.04502627 |
| | hsa-miR-100-3p_syn | 1.45410092 |
| | hsa-miR-206_syn | 2.14180719 |
| **miRNA urine** | hsa-miR-142-3p_uri | 2.73808029 |
| | hsa-miR-376b-3p_uri | 4.21543355 |
| | hsa-miR-203a-3p_uri | 1.87411185 |
| | hsa-miR-6734-5p_uri | 3.67937025 |
| | hsa-miR-203b-5p_uri | 1.06313386 |
| | hsa-miR-223-3p_uri | 1.14906504 |
| | hsa-miR-1299_uri | 3.72244268 |
| | hsa-miR-122-5p_uri | 1.98081001 |
| | hsa-miR-142-5p_uri | 2.06490049 |

**Supplementary Table 13.** Cluster-wise mean Gini impurity-based feature importance across various domains, including clinical data, miRNA plasma, miRNA synovial, miRNA urine, and metabolite plasma, within the integrated model classifying WOMAC function response.

| Cluster 1 vs 2 and 3 | | |
|---|---|---|
| **Multiomic data domain** | **Features** | **Importance Score** |
| **Clinical** | Age | 0.94286047 |
| | Sex | 2.388873 |
| | BMI | 1.97431496 |
| | HADS anxiety category | 1.89995424 |
| | HADS depression category | 1.00883813 |
| | painDetect category | 2.08640528 |
| **Metabolites plasma** | PC.aa.C38.4_pla | 2.03586275 |
| | PC.aa.C38.0_pla | 3.62434475 |
| | lysoPC.a.C20.4_pla | 0.87961106 |
| | PC.aa.C40.6_pla | 1.94841974 |
| | PC.aa.C36.0_pla | 1.94552069 |
| | Tyrosine_pla | 0.84626035 |
| | PC.aa.C38.6_pla | 2.11868538 |
| | trans.hydroxyproline_pla | 1.07440138 |
| | PC.aa.C34.2_pla | 3.99771048 |
| | PC.aa.C28.1_pla | 3.93123781 |
| **miRNA plasma** | hsa-miR-1255a_pla | 2.65574024 |
| | hsa-miR-3173-5p_pla | 3.1299621 |
| | hsa-miR-497-5p_pla | 3.89838523 |
| | hsa-miR-320d_pla_pla | 2.05796376 |
| | hsa-miR-374a-3p_pla | 3.97898 |
| **miRNA synovial fluid** | hsa-let-7f-1-3p_syn | 2.59626386 |
| | hsa-miR-766-3p_syn | 1.80682419 |
| | hsa-miR-921_syn | 2.01680634 |
| | hsa-miR-153-5p_syn | 1.10479038 |
| | hsa-miR-500a-5p_syn | 0.91661593 |
| | hsa-miR-1227-5p_syn | 3.3905707 |
| | hsa-miR-26b-5p_syn | 2.8814131 |
| | hsa-miR-323b-5p_syn | 3.95137329 |
| | hsa-miR-3609_syn | 1.07979891 |
| | hsa-miR-429_syn | 2.33779638 |
| | hsa-miR-2053_syn | 1.95484463 |
| | hsa-miR-144-5p_syn | 2.28171868 |
| | hsa-miR-485-3p_syn | 2.75747886 |
| | hsa-miR-206_syn | 1.38755098 |
| **miRNA urine** | hsa-miR-155-5p_uri | 1.84404923 |
| | hsa-miR-577_uri | 1.09721168 |
| | hsa-miR-142-3p_uri | 1.55552868 |
| | hsa-miR-455-3p_uri | 2.52293438 |
| | hsa-miR-654-3p_uri | 2.07015368 |
| | hsa-miR-122-5p_uri | 1.94307562 |
| | hsa-miR-379-5p_uri | 2.14781474 |
| | hsa-miR-10a-3p_uri | 3.60552423 |
| | hsa-miR-222-3p_uri | 3.70634973 |
| | hsa-miR-3184-3p_uri | 3.72100055 |
| | hsa-miR-136-5p_uri | 2.02380787 |
| | hsa-miR-125b-1-3p_uri | 0.99995869 |
| | hsa-miR-449c-5p_uri | 0.84903817 |
| | hsa-miR-375-3p_uri | 0.8288031 |
| | hsa-miR-100-3p_uri | 1.47077959 |
| | hsa-miR-377-3p_uri | 1.38795262 |
| | hsa-miR-219a-5p_uri | 1.28531554 |
| | hsa-miR-4755-3p_uri | 1.01120271 |
| | hsa-miR-625-3p_uri | 3.75370186 |
| | hsa-miR-487b-3p_uri | 1.92969695 |
| | hsa-miR-127-3p_uri | 1.0448223 |
| | hsa-miR-494-3p_uri | 1.86235454 |
| | hsa-miR-411-5p_uri | 2.38014099 |
| | hsa-miR-1271-5p_uri | 1.95246158 |
| | hsa-miR-653-3p_uri | 1.9337109 |
| | hsa-miR-664a-3p_uri | 2.00033386 |
| | hsa-miR-142-5p_uri | 2.38137853 |
| | hsa-miR-432-5p_uri | 1.15290636 |

| | | |
|---|---|---|
| | hsa-miR-204-3p_uri | 1.15906057 |
| | hsa-miR-489-3p_uri | 2.142128 |
| | hsa-miR-514a-3p_uri | 1.43110079 |
| | hsa-miR-409-3p_uri | 2.15966028 |
| | hsa-miR-223-3p_uri | 0.90435103 |
| | hsa-miR-191-5p_uri | 0.82271918 |
| | hsa-miR-30a-3p_uri | 0.98451173 |
| **Cluster 2 vs 1 and 3** | | |
| **Multiomic data domain** | **Features** | **Importance Score** |
| **Clinical** | Age | 1.82093034 |
| | Sex | 1.03221949 |
| | BMI | 1.01615208 |
| | HADS anxiety category | 1.79010765 |
| | HADS depression category | 1.17661817 |
| | painDetect category | 1.78147877 |
| **Metabolites plasma** | SM.OH.C22.2_pla | 2.89845701 |
| | Glutamine_pla | 2.71599654 |
| | PC.ae.C36.3_pla | 3.34602758 |
| | Aspartic.acid_pla | 2.91132569 |
| | Serine_pla | 3.02488427 |
| **miRNA plasma** | hsa-miR-3942-5p_pla | 2.60203155 |
| | hsa-miR-1976_pla | 2.37536772 |
| | hsa-miR-379-5p_pla | 2.56524003 |
| | hsa-miR-30b-5p_pla | 3.30730435 |
| | hsa-miR-4685-3p_pla | 3.54581869 |
| | hsa-miR-1260b_pla | 3.83125095 |
| | hsa-miR-676-3p_pla | 1.95637453 |
| **miRNA synovial fluid** | hsa-miR-941_syn | 2.18006411 |
| | hsa-miR-378c_syn | 0.94546787 |
| | hsa-miR-1265_syn | 1.64693258 |
| | hsa-miR-6516-5p_syn | 1.89850973 |
| | hsa-miR-642a-5p_syn | 1.44377568 |
| | hsa-miR-33a-3p_syn | 2.3846684 |
| | hsa-miR-3928-3p_syn | 3.15686612 |
| | hsa-miR-3605-3p_syn | 3.27233063 |
| | hsa-miR-101-3p_syn | 3.50658455 |
| | hsa-miR-1307-3p_syn | 3.74865541 |
| | hsa-miR-642b-3p_syn | 1.70180026 |
| | hsa-miR-660-3p_syn | 0.94615276 |
| | hsa-miR-548aw_syn | 1.45921915 |
| | hsa-miR-1251-5p_syn | 2.01346689 |
| | hsa-miR-1537-5p_syn | 3.7933631 |
| | hsa-miR-548aq-3p_syn | 1.59354954 |
| | hsa-miR-758-5p_syn | 1.24531772 |
| | hsa-miR-495-3p_syn | 1.82237675 |
| | hsa-miR-20b-5p_syn | 1.08097323 |
| | hsa-miR-496_syn | 1.46381225 |
| | hsa-miR-4454_syn | 1.31223773 |
| | hsa-miR-362-5p_syn | 1.53443136 |
| | hsa-miR-4732-3p_syn | 2.19276904 |
| | hsa-miR-34b-3p_syn | 1.31319623 |
| | hsa-miR-100-3p_syn | 1.82770771 |
| | hsa-miR-184_syn | 1.27702509 |
| | hsa-miR-206_syn | 2.29519134 |
| | hsa-miR-145-3p_syn | 1.46857137 |
| | hsa-miR-144-5p_syn | 2.32380973 |
| **miRNA urine** | hsa-miR-584-5p_uri | 2.7760144 |
| | hsa-miR-26a-1-3p_uri | 2.77835001 |
| | hsa-miR-22-3p_uri | 3.53737291 |
| | hsa-miR-146b-3p_uri | 3.77287329 |
| | hsa-miR-192-5p_uri | 3.98693034 |
| | hsa-miR-223-5p_uri | 2.04794709 |
| **Cluster 3 vs 1 and 2** | | |
| **Multiomic data domain** | **Features** | **Importance Score** |
| **Clinical** | Age | 1.42366873 |
| | Sex | 1.91838196 |
| | BMI | 1.66273134 |
| | HADS anxiety category | 2.12157328 |
| | HADS depression category | 2.22516654 |

| | | |
|---|---|---|
| | painDetect category | 2.27871996 |
| **Metabolites plasma** | Glutamine_pla | 2.86884398 |
| | Ornithine_pla | 3.47148768 |
| | Arginine_pla | 3.68846514 |
| | PC.ae.C32.1_pla | 3.9211561 |
| | Asparagine_pla | 3.91641089 |
| **miRNA plasma** | hsa-miR-196a-5p_pla | 1.38210858 |
| | hsa-miR-642a-3p_pla | 1.39883847 |
| | hsa-miR-6734-5p_pla | 1.88710433 |
| | hsa-miR-126-3p_pla | 2.32247383 |
| | hsa-miR-181d-5p_pla | 3.44314266 |
| | hsa-miR-96-5p_pla | 3.52207447 |
| | hsa-miR-411-5p_pla | 3.56178815 |
| | hsa-miR-122-3p_pla | 1.35478208 |
| **miRNA synovial fluid** | hsa-miR-6500-3p_syn | 1.6187221 |
| | hsa-miR-6510-5p_syn | 1.26845978 |
| | hsa-miR-1287-3p_syn | 2.13139234 |
| | hsa-miR-6511b-3p_syn | 1.32854143 |
| | hsa-miR-579-3p_syn | 0.80311064 |
| | hsa-miR-335-3p_syn | 0.83107646 |
| | hsa-miR-3913-5p_syn | 1.38521487 |
| | hsa-miR-6780a-5p_syn | 1.05405626 |
| | hsa-miR-326_syn | 1.83395158 |
| | hsa-miR-106b-5p_syn | 0.81211786 |
| | hsa-miR-20b-5p_syn | 0.89287769 |
| | hsa-miR-181b-2-3p_syn | 1.20715484 |
| | hsa-miR-3684_syn | 1.50354307 |
| | hsa-miR-2114-3p_syn | 1.5611783 |
| | hsa-miR-4520-3p_syn | 2.04128862 |
| | hsa-miR-4685-3p_syn | 1.06143314 |
| | hsa-miR-885-5p_syn | 2.08055786 |
| | hsa-miR-500b-3p_syn | 0.99389318 |
| | hsa-miR-30b-3p_syn | 1.42505859 |
| | hsa-miR-891a-5p_syn | 2.17692876 |
| | hsa-miR-548d-3p_syn | 2.1601428 |
| | hsa-miR-412-5p_syn | 2.29523193 |
| **miRNA urine** | hsa-miR-501-5p_uri | 1.75996513 |
| | hsa-miR-142-3p_uri | 0.91867489 |
| | hsa-miR-4763-3p_uri | 2.09210684 |
| | hsa-miR-378c_uri | 1.11401454 |
| | hsa-miR-223-3p_uri | 1.25481765 |
| | hsa-miR-1269a_uri | 1.68342099 |
| | hsa-miR-27a-5p_uri | 0.9138626 |
| | hsa-miR-205-5p_uri | 2.02228188 |
| | hsa-miR-607_uri | 2.29897897 |
| | hsa-miR-4662b_uri | 2.15476001 |
| | hsa-miR-148a-5p_uri | 0.92775078 |
| | hsa-miR-143-3p_uri | 1.74453418 |
